# Global Prevalence of Long COVID, its Subtypes and Risk factors: An Updated Systematic Review and Meta-Analysis

**DOI:** 10.1101/2025.01.01.24319384

**Authors:** Yiren Hou, Tian Gu, Zhouchi Ni, Xu Shi, Megan L. Ranney, Bhramar Mukherjee

## Abstract

**Importance:** Updated knowledge regarding the global prevalence of long COVID (or post-COVID-19 condition), its subtypes, risk factors, and variations across different follow-up durations and geographical regions is necessary for informed public health recommendations and healthcare delivery.

**Objective:** The primary objective of this systematic review is to evaluate the global prevalence of long COVID and its subtypes and symptoms in individuals with confirmed COVID-19 diagnosis, while the secondary objective is to assess risk factors for long COVID in the same population.

**Data Sources:** Studies on long COVID published from July 5, 2021, to May 29, 2024, searched from PubMed, Embase, and Web of Science were used for this systematic review. Supplemental updates to the original search period were made.

**Study Selection:** There were four inclusion criteria: (1) human study population with confirmed COVID-19 diagnosis; (2) appropriate index diagnosis date; (3) outcome must include either prevalence, risk factors, duration, or symptoms of long COVID; and (4) follow-up time of at least two months after the index date. The exclusion criteria were: (1) non-human study population; (1) case studies or reviews; (2) studies with imaging, molecular, and/or cellular testing as primary results; (3) studies with specific populations such as healthcare workers, residents of nursing homes, and/or those living in long-term care facilities; and (4) studies that did not meet the sample size threshold needed to estimate overall prevalence with margin of error of 0.05.

**Data Extraction and Synthesis:** Two screeners independently performed screenings and data extraction, and decision conflicts were collectively resolved. The data were pooled using a random-effects meta-analysis framework with a DerSimonian-Laird inverse variance weighted estimator.

**Main Outcomes and Measures:** The primary estimand (target population parameter of interest) was the prevalence of long COVID and its subtypes among individuals with confirmed COVID-19 diagnoses, and the secondary estimand was effect sizes corresponding to ten common risk factors of long COVID in the same population.

**Results:** A total of 442 studies were included in this mega-systematic review, and 429 were meta-analyzed for various endpoints, avoiding duplicate estimates from the same study. Of the 442 studies, 17.9% of the studies have a high risk of bias. Heterogeneity is evident among meta-analyzed studies, where the *I*^2^ statistic is nearly 100% in studies that estimate overall prevalence. Global estimated pooled prevalence of long COVID was 36% among COVID-19 positive individuals (95% confidence interval [CI] 33%-40%) estimated from 144 studies. Geographical variation was observed in the estimated pooled prevalence of long COVID: Asia at 35% (95% CI 25%-46%), Europe at 39% (95% CI 31%-48%), North America at 30% (95% CI 24%-38%), and South America at 51% (95% CI 35%-66%). Stratifying by follow-up duration, the estimated pooled prevalence for individuals with longer follow-up periods of 1 to 2 years (47% [95% CI 37%-57%]) compared to those with follow-up times of less than 1 year (35% [95% CI 31%-39%]) had overlapping CI and were therefore not statistically distinguishable. Top five most prevalent long COVID subtypes among COVID-19 positive cases were respiratory at 20% (95% CI 14%-28%) estimated from 31 studies, general fatigue at 20% (95% CI 18%-23%) estimated from 121 studies, psychological at 18% (95% CI 11%-28%) estimated from 10 studies, neurological at 16% (95% CI 8%-30%) estimated from 23 studies, and dermatological at 12% (95% CI 8%-17%) estimated from 10 studies. The most common symptom based on estimated prevalence was memory problems estimated at 11% (95% CI 7%-19%) meta-analyzed from 12 studies. The three strongest risk factors for long COVID were being unvaccinated for COVID-19, pre-existing comorbidity, and female sex. Individuals with any of these risk factors had higher odds of having long COVID with pooled estimated odds ratios of 2.34 (95% CI 1.49-3.67) meta-analyzed from 6 studies, 1.59 (95% CI 1.28-1.97) from 13 studies, and 1.55 (95% CI 1.25-1.92) from 22 studies, respectively.

**Conclusions and Relevance:** This study shows long COVID is globally prevalent in the COVID-19 positive population with highly varying estimates. The prevalence of long COVID persists over extended follow-up, with a high burden of symptoms 1 to 2 years post-infection. Our findings highlight long COVID and its subtypes as a continuing health challenge worldwide. The heterogeneity of the estimates across populations and geographical regions argues for the need for carefully designed follow-up with representative studies across the world.

**Key points:** *Question:* What are the prevalence and patterns of long COVID and its subtypes, and what are the risk factors of long COVID?

*Results:* Meta-analysis of 429 studies published from 2021-2024 estimated a pooled global long COVID prevalence of 36% in COVID-19 positive individuals. Variations in geographical regions showed that South America had the highest pooled prevalence of 51% (95% CI: 35%-66%), and the prevalence does not seem to diminish with extended follow-up (less than 1 year: 35%, 95% CI: 31%-39% vs. 1 to 2 years: 47%, 95% CI: 37%-57%). The estimated pooled prevalence of eight major long COVID subtypes in COVID-19 positive individuals were 20% (respiratory), 20% (general fatigue), 18% (psychological), 16% (neurological), 12% (dermatological), 10% (cardiovascular), 9% (musculoskeletal) and 5% (gastrointestinal).

*Meaning:* Quantitative evidence shows a persistent prevalence of long COVID globally, with a significant burden of symptoms 1 to 2 years post-infection, underscoring the need for having accurate and standardized diagnostic tests and biomarkers for long COVID, a better understanding of the physiology of the condition, its treatment, and its potential effect on healthcare needs and workforce participation. The heterogeneity and wide range of the prevalence estimates call for representative samples in well-designed follow-up studies of long COVID across the world.

## Introduction

The Coronavirus Disease 2019 (COVID-19) pandemic, caused by the severe acute respiratory syndrome coronavirus 2 (SARS-CoV-2), has presented unprecedented challenges to public health and healthcare systems worldwide in the last five years. As of December 8, 2024, more than 777 million COVID-19 cases and 7 million deaths have been documented worldwide [1]. Those who survived COVID-19 are known to be at-risk for long COVID, a complex multisystemic disease with sequelae across almost every organ system [2]. Such conditions have been clinically known as post-acute sequelae of COVID-19 (PASC) [3] and referred to as long COVID in the United States [4]. In literature and media, long COVID has been given a broad nomenclature: Post-Acute COVID-19 Syndrome (PACS) [5], Chronic COVID-19 Syndrome [6], Long Haul COVID-19 [7], and COVID Long Haulers [8]. Due to the many names that characterize the long-term health effects of COVID-19, the World Health Organization (WHO) proposed a clinical definition and a name “post-COVID-19 condition” to unify existing definitions [9]. The *International Classification of Diseases*, *Eleventh Revision* (ICD-11) also established RA02 as the code corresponding to post-COVID-19 condition [10]. In this paper, we will use long COVID to describe our phenotype.

By clinical definition, “post COVID-19 condition occurs in individuals with a history of probable or confirmed SARS CoV-19 infection, usually 3 months from the onset of COVID-19, with symptoms that last for at least 2 months and cannot be explained by an alternative diagnosis” [11]. The Institute for Health Metrics and Evaluation (IHME) estimated that by the end of 2021, 3.7% or 144.7 million people developed long COVID as defined by the WHO clinical case definition, with 15.1% or 22 million having persistent symptoms at 12 months after infection onset [12]. These persistent symptoms and sequelae of COVID-19 are of growing interest to public health professionals not just for their impact on those with the diagnosis, but also for their ability to illuminate other infection-related chronic illnesses. To address the healthcare and societal challenges of a significantly large population that is battling with lingering aftermath of COVID, it is important to synthesize existing evidence to enable data-driven decision and policy making.

Existing reviews and analyses typically focus on specific long COVID symptoms or the broad long COVID phenotype. In their *Nature* review article, Davis et al. [13] explored current literature on long COVID subtypes, such as neurological, cardiovascular, pulmonary, and immune symptoms, and their risk factors. A systematic review and meta-analysis of long-term sequelae of COVID-19 estimated that 41.7% of COVID-19 survivors experienced at least one unresolved symptom and 14.1% were unable to return to work at a 2-year follow-up after SARS-CoV-2 infection [14]. Recent meta-analyses also found that while some symptom prevalence estimates decreased, prevalence estimates for fatigue and anosmia at 12-month follow-up remained consistent [15], and female sex, older age, and high BMI were associated with an increased risk of developing long COVID [16]. However, the underlying prevalence of each subtype in a well-curated list of long COVID subtypes with consideration of a large set of symptoms and risk factors for long COVID has not been examined in a unified manner.

Approximately 4 years into the COVID-19 pandemic, there are now adequate numbers of large, high-quality studies on long COVID and symptoms with a longer follow-up time than that in our previous study by Chen et al [17]. An updated meta-analysis is necessary to assess the global prevalence of long COVID. Motivated by our prior study on the global prevalence of long COVID [17], the objective of this updated systematic review and meta-analysis is to investigate the global prevalence and risk factors of long COVID, and the prevalence of eight major subtypes and 40 specific symptoms, nested within the subtypes.

## Methods

### Search strategy

We used the Population, Intervention, Comparison, and Outcome (PICO) framework and Preferred Reporting Items for Systematic Reviews and Meta-Analyses (PRISMA) [18] framework to guide our entire search process (Supplementary eTable 1). Three literature databases, PubMed, Embase, and Web of Science Core Collection, were searched on May 29, 2024, with a supplementary search conducted on July 23, 2024. The supplementary search included a second search to find papers published from May 29, 2024, to July 23, 2024, in the same three databases and a grey literature search on major journals, Google Scholar, and Latin American and Caribbean Health Sciences Literature (Supplementary eMethods 1). The search aimed to capture papers published between 2021 and 2024 related to long COVID and to examine prevalence, risk factors, and/or duration, subtypes and symptoms. To prevent language and index bias, we avoided using an English-only filter to exclude journals originally published in non-English languages, and we excluded publications without English translation during the screening process. The search strategy included search blocks on long COVID, the outcome of interest, and subtypes and symptoms. More than 600 keywords that characterize long COVID subtypes and symptoms were used in constructing the search blocks. The full search strategy, including filters for each database, is presented in Supplementary eMethods 1.

### Screening Procedure

We conducted a two-step screening approach: initially screening based on title/abstract and then a full-text screening. Two human screeners (screeners 1 and 2) independently performed both screening phases. After each screening, any conflicting decisions were resolved through discussion and re-examination. Our inclusion criteria are as follows: (1) human study population with confirmed COVID-19 diagnosis through polymerase chain reaction (PCR) test, antibody test, or a clinical diagnosis; (2) index date of first test/diagnosis, date of hospitalization, discharge date, date of clinical recovery/negative test, or date of symptom occurrence; (3) outcome must include prevalence, risk factors, duration, or symptoms of long COVID; and (4) the follow-up time was at least two months after the index date. The definition of long COVID from the World Health Organization (WHO) was applied in criteria (4).

Our exclusion criteria include: (1) non-human study population; (2) case studies or reviews; (3) studies with imaging, molecular, and/or cellular testing as primary results; (4) studies with only healthcare workers, residents of nursing homes, and/or those in long-term care facilities (as these high-risk populations may skew the estimates); and (5) studies with only overall prevalence outcome that did not meet the sample size threshold of 323 to ensure a margin of error of 0.05 with an expected a priori’ prevalence of 30% (more details in Statistical Analysis and Supplementary eMethods 2).

### Data Extraction

After studies were selected from full-text screening, the relevant data were manually extracted by both screeners: article title, authors, date of publication, study purpose, study design, population, setting, country, total sample size, long COVID samples and non-long COVID samples, method of COVID-19 confirmation, index date, follow-up time, demographic variables (i.e., age and sex), classified long COVID subtypes, and outcomes examined.

### Outcome and Measures

The primary estimand of interest was the prevalence of long COVID, subtypes, and symptoms at least two months after the index date, across various follow-up times and geographic regions. For this systematic review, long COVID is defined as at least one new or persisting symptom during the follow-up time. Eight long COVID subtypes (guided by the reports in extant papers) and 40 related symptoms were tabulated: Neurological (11 related symptoms), Psychological (5 symptoms), Cardiovascular (4 symptoms), Respiratory (6 symptoms), Gastrointestinal (4 symptoms), Musculoskeletal (3 symptoms), Dermatological (2 symptoms), General (5 symptoms). The initial tabulation of subtypes and symptoms is presented in Supplementary eTable 2. To ensure accuracy in the meta-analysis, we included only the subtypes and symptoms reported in more than three studies, resulting in 8 subtypes and 40 distinct symptoms. Additional descriptions of subtypes and symptoms are in Supplementary eTable 3. Follow-up time is stratified by either less than 1 year (< 1 year), between 1 and 2 years (1 – 2 years), and 2 or more years (2+ years). Based on the study cohorts’ nationalities, studies were categorized into six continental regions (Africa, Asia, Europe, North America, South America, and Oceania). We also conducted a stratified analysis based on the clinical study population: hospitalized, non-hospitalized, and a mix of both.

The secondary estimands were odds ratios/risk ratios corresponding to ten potential risk factors for long COVID, including age per year, at least one comorbidity, cardiovascular disease, diabetes, female sex, history of chronic obstructive pulmonary disease, pre-existing hypertension, intensive care unit (ICU admission), obesity, and being unvaccinated for COVID-19. The list of relevant comorbidities varies across studies and commonly includes diabetes, hypertension, chronic heart disease, asthma, dyslipidemia, HIV, chronic kidney disease, chronic liver disease, pulmonary disease, and stroke [19-21]. Additional comorbidities such as allergic rhinitis and eczema sporadically are considered by Li et al. [22], and gastrointestinal disease and major depressive disorder are considered by Jangnin et al. [23].

### Statistical Analysis

For the primary aim, we used a random effects model with inverse variance weighting to meta-analyze the prevalence of long COVID, subtypes, and symptoms. The between-study variance is estimated by the DerSimonian-Laird estimator (Supplementary eMethods 3). Sample size thresholds were applied to include the qualified papers. For studies that reported the overall prevalence of long COVID, we used a sample size threshold of 323 to ensure a margin of error of 0.05 with an expected a priori’ prevalence of 30% reported in our previous meta-analysis. For studies that reported subtype and/or symptom prevalence, the sample size threshold was computed using the estimated pooled prevalence from all relevant studies as the expected prevalence for sample size calculations. Details of sample size calculation and margin of error for each subtype and/or symptom sample size are specified in Supplementary eMethods 2. Heterogeneity between studies was assessed by the *I*^2^ statistic, with *I*^2^ between 75% and 100% indicating considerable heterogeneity. The same procedure is applied to analyze different follow-up times and geography-specific analyses. For our secondary aim, log odds ratios corresponding to the ten risk factors are meta-analyzed using the same procedure.

The risk of bias analysis was conducted following a checklist-based tool for prevalence studies from Joanna Briggs Institute (JBI) [24], and publication bias was evaluated by funnel plots for asymmetry and Egger’s and Begg’s test of association (Supplementary eFigure 1). All analysis was conducted in R (4.3.2) using packages meta [25,26] and metafor [27].

## Results

### Search Results

From our initial literature search in May 2024, we collected 8,515 unique publications through abstract and title screening and conducted full-text screening for 1,414 eligible studies. From the first search in May 2024 and a supplementary search in July 2024, a total of 429 studies were included for meta-analysis or quantitative synthesis. The PRISMA flow diagram is presented in Figure 1. A detailed PRISMA flow diagram for the supplementary search is presented in Supplementary eFigure 3. There were 13 studies excluded from the meta-analysis due to overlapping cohorts reporting on the same estimand at different time points. The rationale for these exclusions is provided in Supplementary eMethods 3, and the details of the 13 studies are described in Supplementary eResults 1.

**Figure 1.**
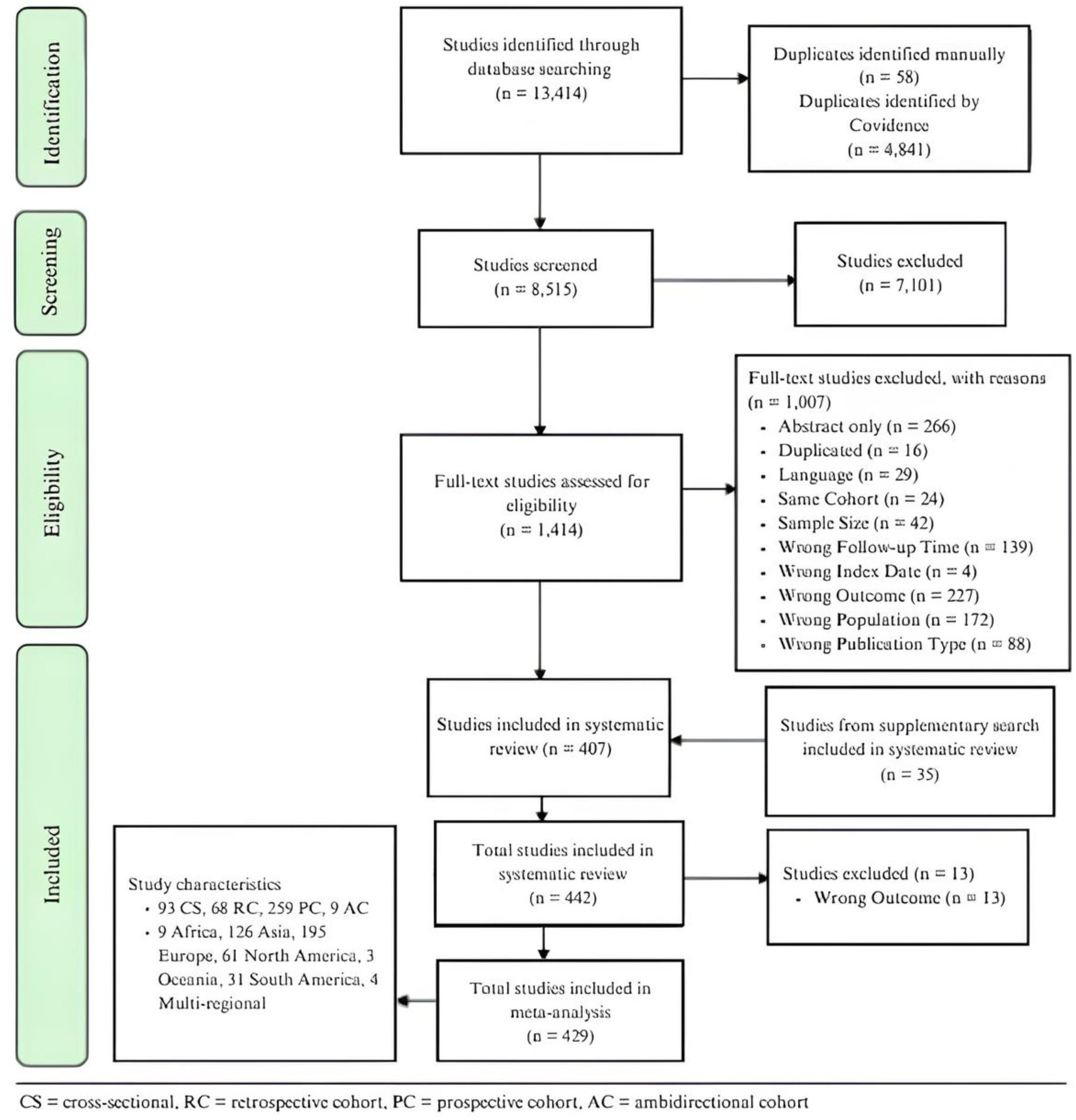
PRISMA Flow Diagram. Additional study characteristics such as cohort type, region, and status of all included studies are listed in the box in the bottom left.

### Study Characteristics

Due to the large number of studies in our systematic review, we summarized the included studies in Supplementary eTable 5. The studies were from six continental regions: Africa (9 studies), Asia (126 studies), Europe (195 studies), North America (61 studies), Oceania (3 studies), South America (31 studies). The other four studies consisted of populations from multiple geographical regions. As shown in Figure 2B, the top five countries with the most studies were the United States (44 studies), China (36 studies), Italy (37 studies), Spain (35 studies), and Brazil (23 studies). In Figure 2, we also plotted the global distribution of studies from our last meta-analysis in 2021 (Chen et al. [17]). The contrast between panels A and B shows that over time, more global literature has emerged, except in Africa and Oceania. Over 2 million individuals who tested positive for or were diagnosed with COVID-19 were included in our current analysis. Among the 144 studies that reported the overall prevalence, we identified specific prevalence rates based on five main factors: hospitalization status (27 studies focused on hospitalized individuals, 13 studies on non-hospitalized individuals, and 104 studies included mixed populations), follow-up duration (122 studies had less than 1 year follow-up, 18 studies had follow-ups of 1 to 2 years, and four studies examined outcomes beyond two years), biological sex (7 studies), and age range (49 studies included all age groups, while 87 studies focused on adults over 18 years old and eight studies with non-adults).

**Figure 2.**
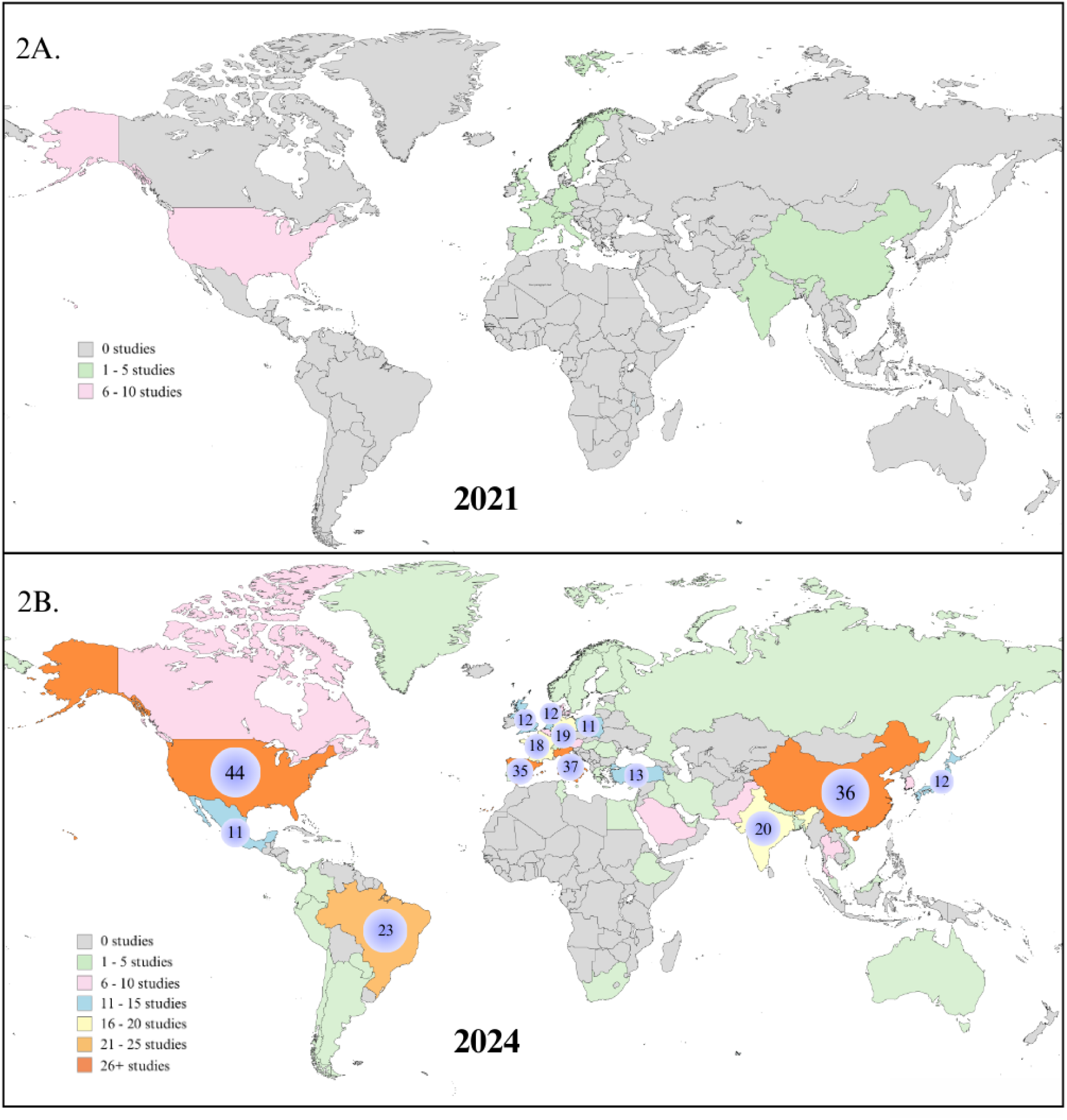
Panel 2A shows global coverage amongst the 33 studies included in meta-analysis by Chen et al. [17]. Panel 2B shows global coverage amongst the 429 studies included in this current meta-analysis. Countries with greater than ten studies are marked with circles indicating the exact number of studies. The lack of studies in the grey areas shows the lack of representation in global datasets and the information gap. The change from 2A to 2B shows that except Africa and Oceania, considerable literature has emerged in the past three years in other parts of the world.

### Pooled Prevalence of Long COVID

As illustrated in Figure 3, we conducted a meta-analysis of 144 studies that reported an overall prevalence of long COVID with a minimum sample size of 323. The pooled global prevalence of long COVID was estimated to be 36% (95% CI 33%-40%) in COVID-19 positive individuals. There was substantial variation among the studies (*I*^2^ = 100%, P < .001), possibly due to the heterogeneity of the studied populations, the changing course of the disease, testing and prevention/treatment strategies over a wide time span from 2021 to 2024 and differences in design/analytic and measurement or coding choices made.

**Figure 3.**
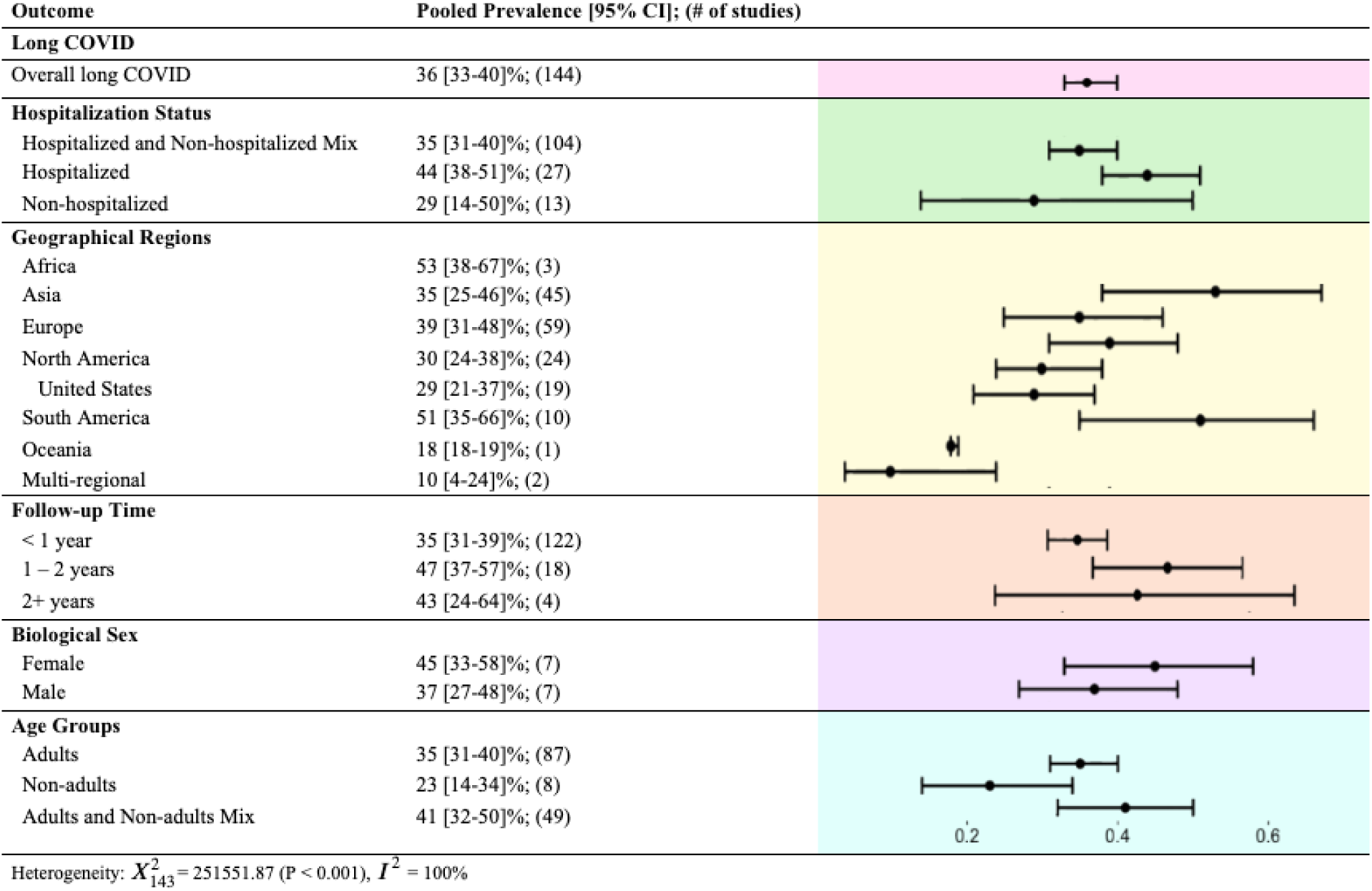
Forest plot for pooled long COVID prevalence, corresponding 95% confidence intervals and number of contributing studies stratified by hospitalization status, geographical regions, follow-up time, biological sex, and age groups.

When looking at the stratified meta-analyses on publication year (Supplementary eFigure 5), the estimated pooled global prevalence of long COVID among COVID-19 cases was 38% (95% CI 28%-50%) ranging from 10% to 62% for studies published in 2021, 37% (95% CI 26%-49%) ranging from 1% to 92% for studies published in 2022, and 37% (95% CI 30%-45%) ranging from 6% to 87% for publications in 2023. The prevalence reduced marginally to 34% (95% CI 29%-41%), ranging from 3% to 80% for publications in 2024. *I*^2^ statistic for studies published in 2021 is 99%, and the *I*^2^ statistic was 100% for studies published in 2022, 2023, and 2024. Although the estimated prevalence remained remarkably consistent over time, the variation between studies remained consistent in every publication year.

### Prevalence of long COVID by Hospitalization Status, Geographic Regions, Follow-up Time, Biological Sex, and Age

When stratified by hospitalization status, the estimated pooled prevalence of long COVID for studies including a mix of hospitalized and non-hospitalized patients was found to be 35% (95% CI 31%-40%). Studies that only involved hospitalized patients showed a higher pooled prevalence of 44% (95% CI 38%-51%), while studies focusing only on non-hospitalized patients showed a lower pooled prevalence of 29% (95% CI 14%-50%). The estimated prevalence varied widely in all three groups: 1% to 89% in the mixed group, 17% to 92% in the hospitalized group, and 3% to 56% in the non-hospitalized group (Supplementary eFigure 7).

When stratifying studies by continents, South America had a higher estimated pooled prevalence of 51% (95% CI 35%-66%), while Asia and Europe had estimates of 35% (95% CI 25%-46%) and 39% (95% CI 31%-48%), respectively. Note that there were less than five studies from Africa included in the analysis, so its estimate should be interpreted with caution. Among studies from North America, the estimated pooled prevalence was 30% (95% CI 24%-38%). Furthermore, a meta-analysis of 19 studies conducted only in the United States showed an estimated pooled prevalence of 29% (95% CI 21%-37%).

When stratifying the studies by follow-up time, we categorized the time from index date as follows: less than 1 year (< 1 year), 1 to 2 years (1 – 2 years), or more than 2 years (2+ years). Among studies with less than 1 year follow-up, the estimated pooled prevalence was 35% (95% CI 31%-39%). A slightly higher pooled prevalence of 47% (95% CI 37%-57%) was estimated by studies with 1 to 2 years of follow-up, and a pooled prevalence of 43% (95% CI 24%-64%) was estimated by studies with more than 2 years of follow-up after the index date. When the follow-up duration was further stratified by hospitalization status, there were no statistically significant differences noted (Supplementary eFigure 6).

Based on seven studies that reported prevalence stratified by biological sex, the estimated pooled prevalence was higher in the female group at 45% (95% CI 33%-58%) than in the male group at 37% (95% CI 27%-48%). Categorizing the study population by age, the estimated pooled prevalence was 35% (95% CI 31%-40%) in adults over 18 years old, and 23% (95% CI 14%-34%) in non-adults. A wider range of prevalence estimates was observed in adults (1%-92%) compared to non-adults (6%-53%, in Supplementary eFigure 7). In the all-age population, the estimated pooled prevalence was 41% (95% CI 32%-50%), with the lowest estimate being 3% and the highest being 89%.

### Prevalence of Specific long COVID Subtypes and Symptoms

We assessed eight subtypes of long COVID based on the reported measures from 429 studies. We stratified each study into less than 1 year (< 1 year) or at least 1 year follow-up time (1 – 2+ years). As shown in Table 1, the five most prevalent subtypes by estimated pooled subtype prevalence were respiratory at 20% (95% CI 14%-28%) estimated from 31 studies, general fatigue at 20% (95% CI 18%-23%) estimated from 121 studies, psychological at 18% (95% CI 11%-28%) estimated from 10 studies, neurological at 16% (95% CI 8%-30%) estimated from 23 studies, and dermatological at 12% (95% CI 8%-17%) estimated from 10 studies. After stratifying by follow-up time, the estimated pooled prevalence of the neurological and general fatigue subtypes increased over time, with the estimated pooled prevalence of the neurological subtype increasing from 13% (95% CI 5%-27%) in < 1 year to 27% (95% CI 15%-44%) at a follow-up time 1 – 2+ years and that of general fatigue increased from 19% (95% CI 16%-22%) in < 1 year after the index date to 26% (95% CI 20%-33%) at 1 – 2+ years after the index date. The respiratory subtype was observed with a gradual decrease from 20% (95% CI 14%-29%) to 19% (95% CI 14%-26%).

**Table 1.** Pooled inverse-variance weighted estimate of the prevalence of long COVID subtypes and symptoms, with corresponding 95% CI obtained by random effects meta-analysis.

| Subtype/symptom | Pooled estimate<br>[95% CI]; (# of<br>studies) | Subtype/symptom | Pooled estimate<br>[95% CI]; (# of studies) |
| --- | --- | --- | --- |
| Neurological | 16 [8-30]%; (23) | Psychological | 18 [11-28]%; (10) |
| < 1 year | 13 [5-27]%; (16) | < 1 year | 10 [5-20]%; (5) |
| 1 – 2+ years | 27 [15-44]%; (7) | 1 – 2+ years | 29 [14-51]%; (5) |
| Concentration/confusion/brainfog | 4 [3-6]%; (28) | Anxiety | 6 [4-9]%; (26) |
| Headache | 6 [5-8]%; (57) | Depression | 8 [5-13]%; (17) |
| Malaise | 8 [3-20]%; (5) | Insomnia | 6 [4-9]%; (29) |
| Memory problems | 11 [7-19]%; (12) | Mood swings | 7 [3-16]%; (6) |
| Sleep problems | 7 [5-10]%; (16) | PTSD symptoms | 10 [1-56]%; (4) |
| Smell | 5 [4-7] %; (43) |  |  |
| Smell or Taste | 4 [2-8]%; (8) |  |  |
| Taste | 4 [3-7]%; (30) |  |  |
| Tinnitus | 1 [1-2]%; (17) |  |  |
| Tremors/Chills | 2 [1-5]%; (12) |  |  |
| Vision problems | 2 [1-3]%; (12) |  |  |
| Cardiovascular | 10 [4-25]%; (16) | Respiratory | 20 [14-28]%; (31) |
| < 1 year | 8 [2-22]%; (11) | < 1 year | 20 [14-29]%; (25) |
| 1 – 2+ years | 20 [11-33]%; (5) | 1 – 2+ years | 19 [14-26]%; (6) |
| Arrhythmia | 2 [0-17]%; (3) | Breathlessness | 8 [3-22]%; (4) |
| Hypertension | 2 [1-5]%; (5) | Chest pain | 4 [3-5]%; (47) |
| Palpitations | 3 [2-5]%; (31) | Chest tightness | 2 [1-3]%; (7) |
| Tachycardia | 4 [2-7]%; (9) | Cough | 6 [5-7]%; (45) |
|  |  | Dyspnea | 7 [6-10]%; (31) |
|  |  | Nasal congestion | 2 [1-4]%; (9) |
| Musculoskeletal | 9 [5-16]%; (13) | Dermatological | 12 [8-17]%; (10) |
| < 1 year | 8 [5-14]%; (12) | < 1 year | 11 [7-19]%; (7) |
| 1 – 2+ years | 30 [29-32]%; (1) | 1 – 2+ years | 6 [1-37]%; (3) |
| Joint pain | 7 [4-11]%; (14) | Hair loss | 5 [3-8]%; (26) |
| Muscle weakness | 11 [5-23]%; (5) | Skin rash | 2 [1-3]%; (19) |
| Myalgia | 5 [4-7]%; (24) |  |  |
| Gastrointestinal | 5 [5-7]%; (34) | General Fatigue | 20 [18-23]%; (121) |
| < 1 year | 6 [5-7]%; (28) | < 1 year | 19 [16-22]%; (95) |
| 1 – 2+ years | 4 [1-10]%; (6) | 1 – 2+ years | 26 [20-33]%; (26) |
| Abdominal pain | 1 [1-2]%; (15) | Loss of appetite | 2 [2-4]%; (30) |
| Constipation | 1 [0-3]%; (5) | Dizziness | 3 [2-4]%; (38) |
| Diarrhea | 2 [1-3]%; (12) | Fever | 2 [1-3]%; (40) |
| Stomach pain | 2 [1-4]%; (3) | Sore throat | 4 [2-6]%; (25) |
|  |  | Sweats | 2 [1-5]%; (10) |

Among various symptoms measured in our included studies, we summarized 40 distinct key symptoms of long COVID as listed within each subtype in Table 1. The most common symptoms, based on estimated prevalence, were memory problems with an estimated prevalence of 11% (95% CI 7%-19%) meta-analyzed by 12 studies, followed by muscle weakness of 11% (95% CI 5%-23%) by 5 studies, breathlessness of 8% (95% CI 3%-22%) by 4 studies, dyspnea of 7% (95% CI 6%-10%) by 31 studies, joint pain of 7% (95% CI 4%-11%) by 14 studies, and cough of 6% (95% CI 5%-7%) by 45 studies.

The variation between studies in terms of the subtype and symptom-specific prevalences can be observed in the detailed forest plots (Supplementary eFigure 8). Many plots exhibit right skewness, with a few studies have a higher subtype or symptom prevalence. For example, we note that a study by Dryden et al. [28] from South Africa has reported a very high malaise symptom prevalence of 50% (95% CI 48%-53%) compared to the other four studies with estimated prevalence ranging from 2% to 7%.

### Risk of bias and sensitivity analysis for 442 included papers

The risk of bias among 442 studies was assessed using the JBI appraisal checklist [24] for prevalence. A lower score out of 9 represents a higher risk of bias in the study. Within the 442 studies, 4.1% (18 studies) scored a 4/9, and 13.8% (61 studies) scored a 5/9. We conducted a sensitivity analysis by removing these 79 studies (17.9%) with a higher risk of bias (scores of 4/9 and 5/9). The pooled global prevalence of long COVID was estimated to be 35% (95% CI 31%-39%) using 129 studies, and the estimated pooled subtype and symptom prevalence remained consistent. Detailed results from this sensitivity analysis are provided in Supplementary eFigure 4 and eTable 6. Reports on the risk of bias are provided in Supplementary eMethods 4.

### Meta-analysis of Association Parameters Corresponding to Risk Factors for long COVID

Among the ten potential risk factors we assessed, we found that those who were unvaccinated for COVID-19 have significantly higher odds of having long COVID compared to those with any vaccination with pooled estimated odds ratios (ORs) of 2.34 (95% CI 1.49-3.67) from 6 studies. We also found that female sex, those with existing hypertension, and those with obesity had higher odds of having long COVID with pooled estimated ORs of 1.55 (95% CI 1.25-1.92) from 22 studies, 1.41 (95% CI 1.10-1.81) from 9 studies, and 1.28 (95% CI 0.99-1.66) from 6 studies, respectively (Supplementary eFigure 9). In addition, those with pre-existing cardiovascular disease, at least one comorbidity, and ICU admission had higher odds of having long COVID, with the pooled estimated ORs of 1.50 (95% CI 1.24-1.81) from 5 studies, 1.59 (95% CI 1.28-1.97) from 13 studies, and 1.43 (95% CI 1.02-2.02) from 8 studies, respectively (Supplementary eFigure 9). A forest plot of the ten risk factors for long COVID is shown in Figure 4.

**Figure 4.**
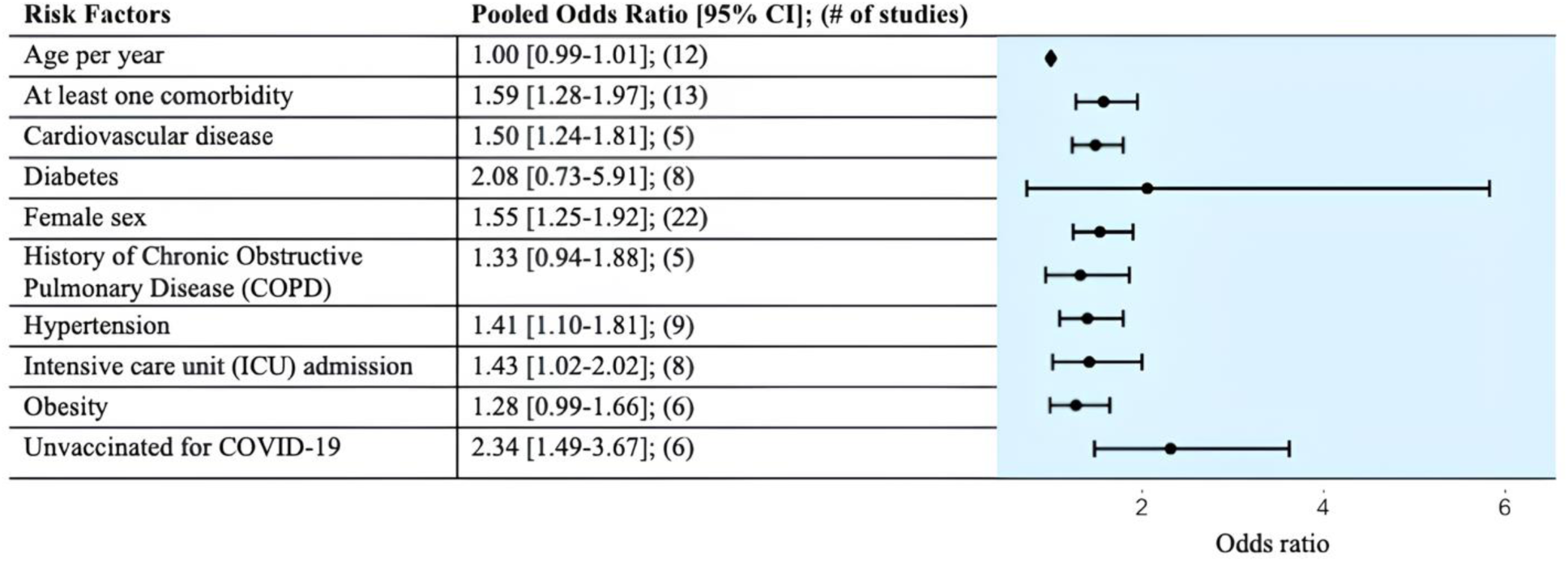
Forest plot for pooled odds ratio estimates for long COVID associated with nine risk factors with corresponding 95% confidence intervals and the number of contributing studies.

We assessed age as a continuous variable and found that older age did not suggest higher odds of having long COVID. Categorizing age may reveal more information, as indicated in studies by Petrakis et al. [29] and Fatima et al. [30], where age was categorized as either less than or equal to 60 years old and greater than 60 years old. In addition, several studies have reported subtype-specific risk factors.

For subtype-specific risk factors, three studies [31-33] described risk factors for neurological subtypes, while Wong-Chew et al. [34], Estrada-Codecido et al. [35], and va Zon et al. [36] provided risk factors for respiratory or cardiovascular subtypes. Since there were fewer than five studies to assess each risk factor for long COVID subtypes, further exploration of meta-analyses of risk factors for subtypes and symptoms was not conducted.

## Discussion

We synthesized information from 442 studies, with 429 contributing to meta-analysis. Our findings suggest a global prevalence of long COVID of approximately 36% among individuals with confirmed COVID-19 diagnoses, although there is a wide range of estimates and large heterogeneity across studies. Thus, these results should be compared with national benchmark estimates when available. We estimated the prevalence of long COVID at 29% (95% CI 21%-37%) among COVID-19 positive individuals in the United States. Our result is strikingly similar to the report by the Household Pulse Survey conducted by the U.S. Census Bureau, which the most recent survey in Phase 4.2 (August 20 to September 16, 2024) estimated 29.8% (95% CI 28.7%-30.8%) among adults who ever had COVID-19 experienced long COVID [37].

In our meta-analysis, the prevalence of long COVID remained consistent irrespective of time since diagnosis, with 35% (95% CI 31%-39%) at less than 1 year follow-up and 47% (95% CI 37%-57%) at one to two years, suggesting a sustained burden of symptoms without a decrease over longer follow-up durations. A similar pattern was observed in the prevalence of neurological subtype and general fatigue prevalence, which increased to 27% and 26%, respectively, at follow-up times of one year or more after the index date, compared to 13% and 19% at less than 1 year follow-up. However, it is important to note that a potential bias in these estimates: follow-up visits may not have been readily available due to healthcare systems’ backlogs during and post-pandemic. Thus, longer follow-up time (>= 1 year) may artificially yield a higher prevalence of long COVID compared to shorter follow-up time (< 1 year of follow-up) due to delayed screening in cohorts that used hospital/clinical records.

A unique strength of the current study is its evaluation of the prevalence of eight distinct subtypes and 40 symptoms associated with long COVID. Among these, neurological symptoms emerge as a detrimental long-term problem for COVID-19 positive individuals, which is rarely seen in common respiratory virus infections [38]. The neurological subtype has an estimated pooled prevalence of 16% of confirmed COVID-19 cases – closely following respiratory (20%), general fatigue (20%), and psychological (18%) subtypes. The prevalence of specific symptoms within the neurological subtypes, such as memory problems (11%) and brain fog (4%) highlight the cognitive impact of long COVID in COVID-19 positive population on a global scale. This finding highlights the need for heightened attention to neurological complications within long COVID care and research.

Among the ten potential risk factors we assessed, we found that those who were unvaccinated for COVID-19 have significantly higher odds of having long COVID compared to those with any vaccination with pooled estimated odds ratios (ORs) of 2.34 (95% CI 1.49-3.67) from 6 studies. There was less variability in the odds ratio estimates than in the prevalence estimates.

The majority of the studies for our systematic review are from Asia, Europe, and North America. Under-represented geographical regions include Africa and Oceania. Of 126 studies from Asia, 28.6% of studies are from the People’s Republic of China, and 15.9% are from India. Countries from the Middle East and Southeast Asia make up 16.7% and 12.7% of studies from Asia. Out of the 195 studies from Europe, most of the publications are distributed across the western and southern regions of Europe. Among study cohorts in South America, 74.2% of the cohorts are from Brazil; and among study cohorts in North America, 72.1% of the cohorts are from the United States. In addition to data disparities between continents, we observe inequities in the availability of data within continents. We also observe considerable heterogeneity (*I*^2^ statistics between 99% to 100%) across geographical regions. A great emphasis should be placed on increasing the number of well-designed studies within and across continents to reduce uncertainties in our prevalence estimates.

Our study builds on the growing body of research on the prevalence of and risk factors associated with long COVID, aligning with recent findings by Al-Aly et al. [39], emphasizing the substantial and lasting impact of long COVID on healthcare systems and public health policy. By evaluating various subtypes and symptoms of long COVID, we provide quantitative insights into the diverse manifestations of long COVID in the global population. Our results highlight the need for longitudinal studies to investigate the progression of long COVID subtypes and offer guidance for personalized care addressing the heterogeneity of the gamut of symptoms that have been reported. Furthermore, these findings support the call for enhanced health education on long COVID, improvement in data inequities and well-developed studies within the global medical community.

### Limitations

We acknowledge the presence of language bias as we only included studies written in English. The supplementary search for grey literature, i.e., information outside traditional databases, was not comprehensive. Due to the abundant grey literature on long COVID from July 5, 2021, we only searched for publications from January 1, 2024, to July 23, 2024, on Google Scholar and Latin American and Caribbean Health Sciences Literature (LILACS). We scanned major journals for studies published in 2024 as well. The included papers from grey literature supplementary search were a part of the supplementary search conducted on July 23, 2024, and were added after data extraction conducted in our first search from July 5, 2021, to May 29, 2024. It is possible that more publications exist from studies completed between 2021 and 2024.

Another major limitation is that we did not meta-analyze the downstream long COVID related outcomes such as quality of life, functional status, mortality and survival have been documented elsewhere [40-44] in the literature. Among the studies included in our paper, 62 studies assessed the quality of life (Supplementary eTable 4). These studies reported a decrease in average functional and physical health scoring patterns among patients with long COVID and reported continuing mental health issues such as anxiety and depression among COVID-19 survivors. Eight studies drew attention to a decline in survival among patients with long COVID, and one study by Wimmer et al. [45] assessed disability in long COVID survivors. These outcomes are critical in understanding the full spectrum of the impact of long COVID. Future systematic reviews should further explore the impact of long COVID on quality of life, mortality and survival, and functional status. This is an exceptionally challenging question as many other factors are at play during a contemporaneous timeframe and it is hard to tease apart the specific contribution of long COVID on downstream outcomes. Nonetheless, we believe that the finest causal discovery and design tools should be applied to answer this question.

There were several measured and unmeasured confounders that could have affected the difference in observed prevalence between the groups. The condition of patients and any pre-existing comorbidities may be factors that differentiate between hospitalized and non-hospitalized individuals. Among hospitalized and non-hospitalized COVID-19 positive individuals, the access to quality testing sites may differ. Those hospitalized were more likely to visit follow-up clinics where healthcare professionals would examine their long COVID, subtypes, and/or symptoms according to appropriate guidelines. In contrast, non-hospitalized individuals, including those in the mixed group, were more likely to receive follow-up surveys, where the respondents would need to assess their own health status. There were obvious risk factors we did not examine, for example, we did not consider different strains of SARS-Cov-2 as a risk factor since there were not enough studies (less than five studies) to meta-analyze. Additionally, variables such as healthcare access, income inequality, and educational awareness are worth examining in relation to long COVID. Collecting and analyzing detailed measures of social determinants of health in properly designed COVID survivorship studies, such as the RECOVER cohort in the United States [46], the COVIMPACT prospective observational cohort in Belgium [47], and the Lifelines biobank in the Netherlands [48] are in order.

To visualize the considerable amount of heterogeneity across the prevalence estimates extracted for meta-analysis, we created boxplots for overall and subtype prevalence estimates in Figure 5. This gives a sense of the range of variability in the estimates. There are several potential sources of this heterogeneity. Some of these were induced by study design and varying definitions of outcomes. Vast variation in how people were labeled as those with or without long COVID, given the absence of any diagnostic biomarkers, contributes to heterogeneity between studies and uncertainties in our prevalence estimates. Additional contributors such as long COVID definition, subtype and symptom definition, and study cohort identification can induce variations among studies. Within the hospitalized COVID-19 positive population, the definition of hospitalization was not consistent. For example, hospitalization was defined as having to be admitted to inpatient care [49], intensive care [50-52], and/or ward units [53,54]. Hospitalization due to other health conditions, such as respiratory diseases [55] or kidney failure [56], in individuals diagnosed with COVID-19 was also present in some studies. Since long COVID subtypes are not well-established, our self-organization of these designations may contribute to heterogeneity among studies used in estimating subtype prevalence. True population heterogeneity due to differences in age structure, healthcare infrastructure, access to vaccines, and biological and genetic differences may also exist but currently we do not have the tools to distinguish between the two sources.

**Figure 5.**
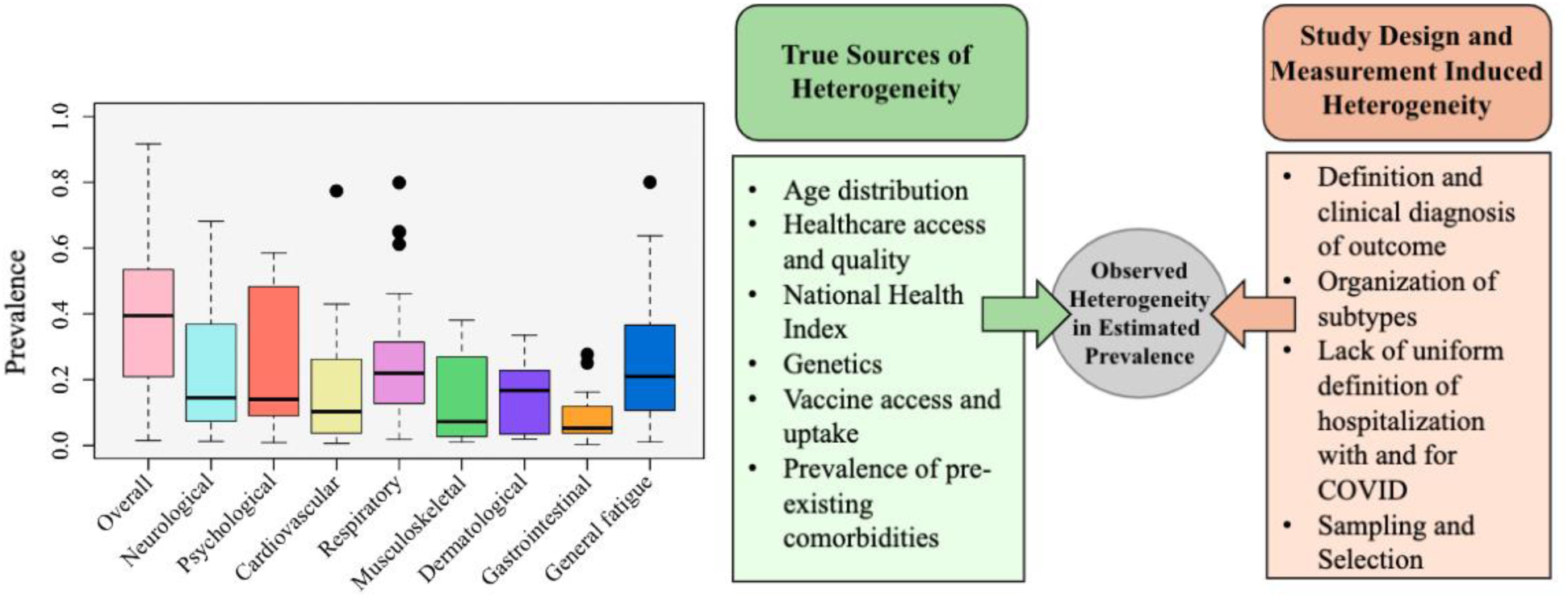
Examination of heterogeneity by boxplot of overall long COVID prevalence, neurological, psychological, cardiovascular, respiratory, musculoskeletal, dermatological, gastrointestinal, and general fatigue subtype prevalence. Potential sources of true etiological heterogeneity versus study-induced heterogeneity are listed.

Finally, with such heterogeneity in estimates across and within geography, one could argue that pooling such a large number of studies across the world is meaningless. While we recognize the challenges of a common effect meta-analysis model in the face of such wide variability [57], the inverse-variance weighted pooled estimate across the globe provides a precision weighted average of available estimates and displays the variation through the forest plots (Supplementary eFigure 7 and 8). Despite all these limitations, the agreement of the pooled US estimate with nationally representative household pulse survey, the consistency of the prevalence estimates over the years, and the robustness of the estimate when studies with a high risk of bias were eliminated gives us some confidence in the legitimacy of the pooled estimates and the analysis, despite the challenges with available data.

## Conclusions

This systematic review and meta-analysis provide a comprehensive overview of empirical estimates regarding the prevalence of long COVID, its subtypes, symptoms, and associated risk factors among individuals with confirmed COVID-19 diagnoses. Our findings reveal significant regional variation and heterogeneity in prevalence estimates, with higher estimates observed in certain geographical areas. However, there was considerable variation in estimates within the same geographical region. Representative samples and well-designed data collection are needed to reduce heterogeneity between studies across the same geography and produce precise estimates of long COVID prevalence. The study also emphasizes global data inequity, with limited representation from populations in Africa and Oceania. Finally, the persistence of long COVID symptoms across varying follow-up durations highlights the long-term burden of these conditions and calls for a better understanding of long COVID physiology, discovery of diagnostic biomarkers for long COVID, its treatment, and its effect on healthcare needs and workforce participation.

## Supporting information

eMethods 1a. Unregistered systematic review protocol

eMethods 1b. Final search strategy

eMethods 1c. Long COVID subtypes justification

eMethods 1d. Supplementary search strategy

eMethods 4. Risk of bias assessment across included studies

eTable 3. Characterization of subtypes and synonyms of symptoms

eTable 5. Summary of included studies

## Data Availability

Data used in this meta-analysis were derived from previously published studies, which are publicly available through databases such as PubMed, Embase, and Web of Science. No new or proprietary data were generated or collected in this study. A summary of the papers used can be found in the Supplement.

## Acknowledgments

The authors thank Harlan Krumholz, MD, SM, affiliated with the Department of Health Policy and Management at Yale School of Public Health, Center for Outcomes Research and Evaluation at Yale-New Haven Hospital, and Section of Cardiovascular Medicine, Department of Internal Medicine at Yale School of Medicine, for guidance on exploration of heterogeneity and study variations in this meta-analysis.

## Author Contributions

Conceptualization: BM

Methodology: BM

Investigation: YH (Screener 1), ZN (Screener 2), BM

Supervision: BM, TG

Writing – original draft: YH, TG, ZN, BM

Writing – review & editing: YH, TG, XS, MLR, BM

## Data Access and Responsibility Statement

The authors take full responsibility for the integrity of the data and the accuracy of the analysis. All authors had access to the data used in this study and have reviewed and approved the final manuscript.

