## Supplementary Materials for "Global Prevalence of Long COVID, its Subtypes and Risk factors: An Updated Systematic Review and Meta-Analysis"

**eMethods 1.** Systematic review procedure

**eMethods 2.** Sample size calculation

**eMethods 3.** Meta-analysis framework

**eMethods 4.** Risk of bias assessment across included articles

**eResults 1.** Summary of outcomes from non-meta-analyzed studies

**eTable 1**. PRISMA checklist

**eTable 2.** Initially tabulated long COVID subtypes and symptoms

**eTable 3.** Characterization and synonyms of symptoms

**eTable 4.** Quality of life, mortality, and other potential outcome summarized table

**eTable 5.** Summary of included studies

**eTable 6.** Pooled inverse-variance weighted estimate of the prevalence of long COVID subtypes and symptoms in COVID-19 positive individuals, with corresponding 95% CI obtained by random effects meta-analysis, studies with high risk of bias (both 4/9 and 5/9 scores) removed

**eFigure 1.** Funnel plot for publication bias assessment among studies used to estimate the pooled long COVID prevalence in COVID-19 positive individuals

**eFigure 2.** PRISMA flow diagram

**eFigure 3.** PRISMA flow diagram for supplementary search

**eFigure 4.** Forest plot for pooled long COVID prevalence in COVID-19 positive individuals, corresponding 95%confidence intervals and number of contributing studies stratified by hospitalization status, geographic region, follow-up time, biological sex, and age-group. Studies with high risk of bias (both 4/9 and 5/9 scores) removed.

**eFigure 5.** Forest plots with the prevalence estimates of long COVID in COVID-19 positive individuals stratified by publication year

**eFigure 6.** Forest plots with the prevalence estimates of long COVID in COVID-19 positive individuals stratified by follow-up time and hospitalization status.

**eFigure 7.** Forest plots with the prevalence estimates of long COVID in COVID-19 positive individuals stratified by A) Biological sex, B) Geographical regions, C) Hospitalization status, D) Follow-up time, E) Age groups

**eFigure 8.** Supplementary meta-analysis of long COVID subtype-specific and symptom-specific prevalence in COVID-19 positive individuals

**eFigure 9.** Forest plots of risk factors for long COVID in COVID-19 positive individuals

**eReferences**

**eMethods 1.** Systematic review procedure

For this updated systematic review on long COVID, we systematically collected publications that concerned prevalence, risk factors, and/or duration of long COVID and its subtypes in any country. Initially, we classified ten subtypes (neurological, psychological, cardiovascular, respiratory, gastrointestinal, renal, musculoskeletal, dermatological, endocrine or metabolic, general), and the subtype justifications are available in ***eMethods 1c***.

To identify relevant publications, we searched in the following databases: PubMed, Embase, and Web of Science Core Collection. The search was conducted on May 29, 2024. Following the complete search conducted by Chen et al. [1] on July 5, 2021, we aimed to capture studies reflecting those available from July 5, 2021 to May 29, 2024. Note the second search on August 12, 2021 and the third search on March 13, 2022 in Chen et al.’s paper was restricted to well-established medical journals, thus we continued from the first search on July 5, 2021. A comprehensive supplementary search is carried out on July 23, 2024 after full-text screening, and the strategy can be found in ***eMethods 1d***.

Search blocks and filters for PubMed, Embase, and Web of Science Core Collection are detailed in ***eMethods 1b***. Search block 1 is from PubMed Clinical queries long COVID filter, which was announced in Fall 2021 and was not available at the time of the original search by Chen et al [1]. Search block 2 is modified from search block 3 of search strategy by Chen et al [1]. Search blocks 3 to 12 are of long COVID subtypes by using symptom keywords and synonyms.

After securing citations from the search engines to Covidence [2], a tool to streamline systematic review, the resulting body of citations were deduplicated and loaded to the screening process. The manual search flow is presented in the PRISMA flowchart in **eFigure 2**. The extended or supplementary search flow is shown in **eFigure 3**. We did prepare a formal systematic review protocol, presented in ***eMethods 1a***, however it was not registered.

***eMethods 1a.*** Unregistered_SystematicReview_Protocol.docx

***eMethods 1b.*** Final_Search_Strategy.docx

***eMethods 1c.*** LongCOVID_Subtypes_Justification.docx

***eMethods 1d.*** Supplementary_Search_Strategy.docx

**eMethods 2.** Sample size calculation

The inclusion/exclusion of sample size in the main text screening was based on margin of error in confidence interval for Binomial proportions. The formula is as follows:

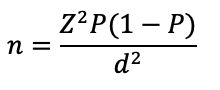
,

where n is sample size, and *Z* denotes the *Z*-statistics for 95% confidence interval. Note the 97.5 percentile point of the standard Normal distribution is approximately 1.96.

For estimating pooled overall prevalence of long COVID, the expected proportion *P* is 0.3, and the margin of error *d* is 0.05. The sample size threshold is at least 323 samples.

For estimating pooled subtype prevalence of long COVID, the expected proportion *P* is the estimate from meta-analyzing all studies that reported the subtype prevalence. The margin of error *d* is 0.05 for neurological, psychological, respiratory, musculoskeletal, dermatological, general fatigue, cardiovascular subtype. The margin of error *d* is 0.03 for gastrointestinal subtype.

For estimating pooled symptom prevalence of long COVID, the expected proportion *P* is the estimate from meta-analyzing all studies that reported the symptom prevalence. The margin of error *d* is 0.02 for symptoms under neurological, respiratory, musculoskeletal, dermatological, general fatigue, cardiovascular subtype. The margin of error *d* is 0.03 for symptoms under psychological subtype. The margin of error *d* is 0.01 for symptoms under gastrointestinal subtype.

The following table summarizes specified margin of errors, estimated expected proportion from meta-analyzing all studies that reported the subtype or symptom prevalence, and the calculated sample size threshold.

| **Long COVID Subtypes** | **Margin of Error** | **Estimated expected Proportion** | **Sample Size Threshold (n)** |
| --- | --- | --- | --- |
| **Neurological Subtype**  **Symptoms**  Concentration/confusion/brain fog  Headache  Malaise  Memory problems  Sleep problems  Smell  Smell or Taste  Taste  Tinnitus  Tremors/Chills  Vision problems | 0.05  0.02 | 0.21  0.08  0.08  0.10  0.13  0.11  0.06  0.09  0.05  0.03  0.03  0.04 | 255  707  707  864  1086  940  542  787  456  279  279  369 |
| **Psychological Subtype**  **Symptoms**  Anxiety  Depression  Insomnia  Mood swings  PTSD symptoms | 0.05  0.03 | 0.25  0.13  0.11  0.07  0.09  0.13 | 288  483  418  278  350  483 |
| **Cardiovascular Subtype**  **Symptoms**  Arrhythmia  Hypertension  Palpitations  Tachycardia | 0.05  0.02 | 0.11  0.02  0.02  0.03  0.06 | 150  188  188  279  542 |
| **Respiratory Subtype**  **Symptoms**  Breathlessness  Chest pain  Chest tightness  Cough  Dyspnea  Nasal congestion | 0.05  0.02 | 0.20  0.20  0.06  0.03  0.08  0.15  0.04 | 246  1537  542  279  707  1225  369 |
| **Musculoskeletal Subtype**  **Symptoms**  Joint pain  Muscle weakness  Myalgia | 0.05  0.02 | 0.11  0.10  0.16  0.10 | 150  864  1291  864 |
| **Dermatological Subtype**  **Symptoms**  Hair loss  Skin rash | 0.05  0.02 | 0.13  0.08  0.03 | 174  707  279 |
| **Gastrointestinal Subtype**  **Symptoms**  Abdominal pain  Constipation  Diarrhea  Stomach pain | 0.03  0.01 | 0.06  0.02  0.02  0.03  0.03 | 241  753  753  1118  1118 |
| **General Fatigue Subtype**  **Symptoms**  Loss of appetite  Dizziness  Fever  Sore throat  Sweats | 0.05  0.02 | 0.24  0.03  0.05  0.02  0.05  0.03 | 280  279  456  188  456  279 |

**eMethods 3.** Meta-analysis framework

We included the reported measure from studies with no overlap between patient cohorts. If there were studies with overlapping cohorts, these studies can be included in the meta-analysis as long as they are reporting different outcomes. In the instance of overlapping cohorts on the same outcome measure, we chose one study, and following Chen et al. we preferred studies with the following characteristics: 1) being published more recently, 2) multicenter, 3) longer follow-up time, and 4) lower risk of bias.

Random effects model was used with logit transformation and the DerSimonian-Laird (DL) [3,4] estimator for between-study variance

$$\tau^{2}$$

. The pooled estimated prevalence of long COVID (
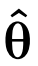
) is calculated as:

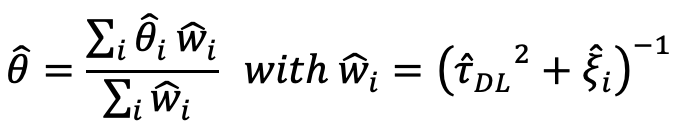
,

where represents the *i*-th study out of total *n* studies, *i* = 1,...,*n*. The logit transformed prevalence
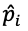
 in study *i* is denoted as
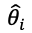
, where it is calculated as:

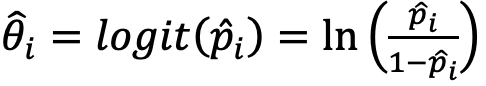
.

The estimated weight for study *i* is
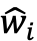
, and the inverse-variance method is used to combine logit transformed prevalence of total *n* studies into a weighted average. The total variance of study *i* is the sum of within-study variability denoted by
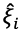
, and between-study variability calculated by the DL estimator
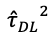
. Then
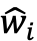
 is the inverse of the total variance.

The DL estimator is calculated as the following:

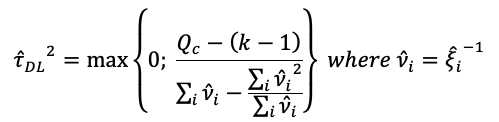
,

and

$$Q_{c}$$

 is the Cochran’s Q statistic that is the weighted sum of the squared deviations of the estimated effects
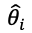
from their weighted mean

$$\overline{\theta}$$

. . The Cochran’s Q statistic is as follows:

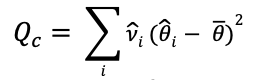
.

**eMethods 4.** Risk of bias assessment across included articles

Risk of bias among 442 studies were assessed using the Joanna Briggs Institute (JBI) appraisal checklist [5] for prevalence studies. Studies were evaluated based on nine common sources of bias in observational studies, and a score out of 9 reflected how well each study addressed bias in its design, conduct, and analysis. A lower score represents a higher risk of bias in the study. Out of the 442 studies in systematic review, 4.1% (18 studies) scored 4/9; 13.8% (61 studies) scored 5/9; 35.3% (156 studies) scored 6/9; 32.8% (145 studies) scored 7/9; 12.7% (56 studies) scored 8/9; and 1.4% (6 studies) scored 9/9. Out of the 429 studies in meta-analysis, 4.2% (18 studies) scored 4/9; 14.0% (60 studies) scored 5/9; 34.7% (149 studies) scored 6/9; 32.9% (141 studies) scored 7/9; 12.8% (55 studies) scored 8/9; and 1.4% (6 studies) scored 9/9.

Supplementary file *Supplementary_Risk_of_Bias_Assessment.xlsx* contains full risk of bias assessment across the included studies.

**eResults 1.** Summary of outcomes from non-meta-analyzed studies

Thirteen studies were not included in the meta-analysis overlapping cohorts on same outcome measures. Malheiro et al. [6] reported separately hospitalized and non-hospitalized COVID-19 positive adults in Hospital Israelita Albert Einstein located in Brazil, and overall long COVID symptom prevalence and risk factors were stratified by hospitalized and non-hospitalized group. Huang et al. [7] reported overall and symptom prevalence of COVID-19 positive adults in Jin Yin-tan Hospital, Wuhan, China. Ye et al. [8] estimated symptom prevalence in anxiety (44.7%), chest tightness (21.2%), and dyspnea (19.1%) and found risk factors for symptom fatigue from hospitalized females in multicenter setting (Huoshenshan Hospital and Taikang Tongji Hospital in Wuhan, China). Pazukhina et al. [9] conducted a multicenter cohort study in Russia, and estimated long COVID symptom prevalence and risk factors were stratified by adults and non-adults. Using data from six Dutch hospitals, Klinkhammer et al. [10] estimated cognitive dysfunction in intensive care unit (37%) and non-intensive care unit group (44%). Plywaczewska-Jakubowska et al. [11] estimated overall prevalence and risk factors for long COVID in non-hospitalized COVID-19 positive adults in Poland. In the United States, Richard et al. [12] Estimated the overall prevalence of long COVID in an Epidemiology, Immunology, and Clinical Characteristics of Emerging Infectious Diseases with Pandemic Potential (EPICC) study implemented in the U.S. military health system. Among COVID-19 positive adults in Mass General Brigham, an academic health system in the Northeast United States, Wang et al. [13] estimated symptom prevalence and risk factors for long COVID.

Several studies evaluated prevalence of long COVID in non-adult cohorts. In an Italian cohort study, Trapani et al. [14] found overall and symptom prevalence and risk factors for long COVID in COVID-19 positive children, stratified by primary care and hospitalized group. Yildirim Arslan et al. [15] reported symptom prevalence, stratified by Delta or Omicron variant COVID-19 cases in children aged four to eighteen from a tertiary-level university in Turkey. Another study in Turkey by Metbulut et al. [16] examined symptom prevalence in pediatric patients with asthma. There were multiple studies analyzing the CLoCk cohort in the United Kingdom, where non-hospitalized non-adults with previous COVID-19 diagnoses were recruited. Stephenson et al. [17,18] evaluated the overall and symptom prevalence in the CLoCk cohort.

**eTable 1**. PRISMA checklist

| **Section and Topic** | **Item #** | **Checklist item** | **Location where item is reported ^a^** |
| --- | --- | --- | --- |
| **TITLE** | | | |
| Title | 1 | Identify the report as a systematic review. | Report page 1 |
| **ABSTRACT** | | | |
| Abstract | 2 | Background: Provide an explicit statement of the main objective(s) or question(s) the review addresses. Methods: Specify the inclusion and exclusion criteria for the review. Specify the information sources (e.g. databases, registers) used to identify studies and the date when each was last searched. Specify the methods used to assess risk of bias in the included studies. Specify the methods used to present and synthesize results. Results: Give the total number of included studies and participants and summarize relevant characteristics of studies. Present results for main outcomes, preferably indicating the number of included studies and participants for each. If meta-analysis was done, report the summary estimate and confidence/credible interval. If comparing groups, indicate the direction of the effect (i.e., which group is favoured). Discussion: Provide a brief summary of the limitations of the evidence included in the review (e.g., study risk of bias, inconsistency and imprecision). Provide a general interpretation of the results and important implications. Other: Specify the primary source of funding for the review. Provide the register name and registration number. | Report page 2 - 4 |
| **INTRODUCTION** | | | |
| Rationale | 3 | Describe the rationale for the review in the context of existing knowledge. | Report page 6 -7 |
| Objectives | 4 | Provide an explicit statement of the objective(s) or question(s) the review addresses. | Report page 7 |
| **METHODS** | | | |
| Eligibility criteria | 5 | Specify the inclusion and exclusion criteria for the review and how studies were grouped for the syntheses. | eMethods 1, Report page 8 - 9 |
| Information sources | 6 | Specify all databases, registers, websites, organizations, reference lists and other sources searched or consulted to identify studies. Specify the date when each source was last searched or consulted. | Report page 8 |
| Search strategy | 7 | Present the full search strategies for all databases, registers and websites, including any filters and limits used. | eMethods 1 |
| Selection process | 8 | Specify the methods used to decide whether a study met the inclusion criteria of the review, including how many reviewers screened each record and each report retrieved, whether they worked independently, and if applicable, details of automation tools used in the process. | eMethods 1, report page 8 - 10 |
| Data collection process | 9 | Specify the methods used to collect data from reports, including how many reviewers collected data from each report, whether they worked independently, any processes for obtaining or confirming data from study investigators, and if applicable, details of automation tools used in the process. | eMethods 1 |
| Data items | 10a | List and define all outcomes for which data were sought. Specify whether all results that were compatible with each outcome domain in each study were sought (e.g., for all measures, time points, analyses), and if not, the methods used to decide which results to collect. | eTable 2, eTable 3 |
|  | 10b | List and define all other variables for which data were sought (e.g., participant and intervention characteristics, funding sources). Describe any assumptions made about any missing or unclear information. | Report page 9 - 10 |
| Study risk of bias assessment | 11 | Specify the methods used to assess risk of bias in the included studies, including details of the tool(s) used, how many reviewers assessed each study and whether they worked independently, and if applicable, details of automation tools used in the process. | eMethods 4 |
| Effect measures | 12 | Specify for each outcome the effect measure(s) (e.g., risk ratio, mean difference) used in the synthesis or presentation of results. | Report page 9 - 10 |
| Synthesis methods | 13a | Describe the processes used to decide which studies were eligible for each synthesis (e.g. tabulating the study intervention characteristics and comparing against the planned groups for each synthesis (item #5)). | Figure 1, Report page 12 - 14 |
|  | 13b | Describe any methods required to prepare the data for presentation or synthesis, such as handling of missing summary statistics, or data conversions. | Report page 12 - 14 |
|  | 13c | Describe any methods used to tabulate or visually display results of individual studies and syntheses. | eTable 5 report page 12 - 14 |
|  | 13d | Describe any methods used to synthesize results and provide a rationale for the choice(s). If meta-analysis was performed, describe the model(s), method(s) to identify the presence and extent of statistical heterogeneity, and software package(s) used. | eMethods 2 and 3, report page 10 11 |
|  | 13e | Describe any methods used to explore possible causes of heterogeneity among study results (e.g., subgroup analysis, meta-regression). | Figure 2, 5, eFigure 4 – 6, eTable 6 |
|  | 13f | Describe any sensitivity analyses conducted to assess robustness of the synthesized results. | eFigure 4 - 6 |
| Reporting bias assessment | 14 | Describe any methods used to assess risk of bias due to missing results in a synthesis (arising from reporting biases). | eMethods 4, report page 20 |
| Certainty assessment | 15 | Describe any methods used to assess certainty (or confidence) in the body of evidence for an outcome. | eMethods 4 and eFigure 1 |
| **RESULTS** | | | |
| Study selection | 16a | Describe the results of the search and selection process, from the number of records identified in the search to the number of studies included in the review, ideally using a flow diagram. | Report page 10, Figure 1 |
|  | 16b | Cite studies that might appear to meet the inclusion criteria, but which were excluded, and explain why they were excluded. | Report page 13, Figure 1 |
| Study characteristics | 17 | Cite each included study and present its characteristics. | eTable 1 |
| Risk of bias in studies | 18 | Present assessments of risk of bias for each included study. | eMethods 4 |
| Results of individual studies | 19 | For all outcomes, present, for each study: (a) summary statistics for each group (where appropriate) and (b) an effect estimate and its precision (e.g. confidence/credible interval), ideally using structured tables or plots. | Figure 3, 4 Table 1, report page 15 - 21 |
| Results of syntheses | 20a | For each synthesis, briefly summarise the characteristics and risk of bias among contributing studies. | eMethods 4 |
|  | 20b | Present results of all statistical syntheses conducted. If meta-analysis was done, present for each the summary estimate and its precision (e.g. confidence/credible interval) and measures of statistical heterogeneity. If comparing groups, describe the direction of the effect. | eFigure 4 - 9, Table 1, report page 15 - 21 |
|  | 20c | Present results of all investigations of possible causes of heterogeneity among study results. | Figure 5, Report page 23 - 27 |
|  | 20d | Present results of all sensitivity analyses conducted to assess the robustness of the synthesized results. | eFigure 1, eFigure 4 - 6 |
| Reporting biases | 21 | Present assessments of risk of bias due to missing results (arising from reporting biases) for each synthesis assessed. | eMethods 4 |
| Certainty of evidence | 22 | Present assessments of certainty (or confidence) in the body of evidence for each outcome assessed. | eFigure 1 |
| **DISCUSSION** | | | |
| Discussion | 23a | Provide a general interpretation of the results in the context of other evidence. | Report page 22 - 24 |
|  | 23b | Discuss any limitations of the evidence included in the review. | Report page 24 - 27 |
|  | 23c | Discuss any limitations of the review processes used. | Report page 24 |
|  | 23d | Discuss implications of the results for practice, policy, and future research. | Report page 28 |
| **OTHER INFORMATION** | | |  |
| Registration and protocol | 24a | Provide registration information for the review, including register name and registration number, or state that the review was not registered. | eMethods 1 |
|  | 24b | Indicate where the review protocol can be accessed, or state that a protocol was not prepared. | eMethods 1 |
|  | 24c | Describe and explain any amendments to information provided at registration or in the protocol. | eMethods 1 |
| Support | 25 | Describe sources of financial or non-financial support for the review, and the role of the funders or sponsors in the review. | Report page 36 |
| Competing interests | 26 | Declare any competing interests of review authors. | Report page 36 |
| Availability of data, code and other materials | 27 | Report which of the following are publicly available and where they can be found: template data collection forms; data extracted from included studies; data used for all analyses; analytic code; any other materials used in the review. | Report page 36 |

**eTable 2.** Initially tabulated long COVID subtypes and symptoms

| Subtype (# of symptoms) |  |
| --- | --- |
| Neurological (27) | Renal (5) |
| Psychological (12) | Musculoskeletal (8) |
| Cardiovascular (17) | Dermatological (4) |
| Respiratory (11) | Metabolic (9) |
| Gastrointestinal (14) | General (11) |

| **Neurological symptoms** | **Psychological symptoms** | **Cardiovascular symptoms** | **Respiratory symptoms** |
| --- | --- | --- | --- |
| Abnormal cerebrospinal fluid | Anxiety | Anemia | Abnormal gas exchange |
| Abnormal movement | Anxiety- and fear-related disorders | Arrhythmias | Asthma |
| Autonomic dysfunction (dysautonomia) | Attention deficit | Bleeding events | Chest pain or discomfort |
| Blurred vision | Delirium | Blood cell alteration | Chronic obstructive pulmonary disease |
| Brain or brain stem hypometabolism | Depression | Bradycardia | Cough/chronic cough |
| Concentration/confusion/brain fog | Insomnia | Cardiac impairment/abnormalities/heart failure | Dyspnea (shortness of breath) |
| Headache | Irritability | Chest pain | Hypoxaemia |
| Hearing impairment | Post-traumatic stress disorder (PTSD) | Coagulation - Thromboembolism | Lower respiratory disease |
| Memory problems/loss | Psychomotor impairment | Coronary atherosclerosis | Nasal congestion |
| Multi-lineage cellular dysregulation | Sleep disturbance | Endothelial inflammation/dysfunction | Pulmonary embolism |
| Myelin loss | Substance abuse | Heart attack | Sleep apnea |
| Nerve inflammation/pain | Trauma- and stress- related disorders | Hypertension |  |
| Neuroimaging changes |  | Microangiopathy |  |
| Numbness/tingling (Paresthesia) |  | Myocardial inflammation |  |
| Oxidative stress |  | Palpitations |  |
| Paralysis |  | Postural orthostatic tachycardia syndrome (POTS) |  |
| Post-exertional malaise |  | Tachycardia |  |
| Reduction in grey matter thickness |  |  |  |
| Seizures |  |  |  |
| Sensorimotor symptoms |  |  |  |
| Serotonin reduction |  |  |  |
| Sleep problems (sleep-wake disorders) |  |  |  |
| Smell (anosmia/hyposmia) |  |  |  |
| Smell or taste (hypogeusia) |  |  |  |
| Stroke |  |  |  |
| Tinnitus |  |  |  |
| Tremor |  |  |  |

| **Gastrointestinal symptoms** | **Renal symptoms** | **Musculoskeletal symptoms** | **Dermatological symptoms** |
| --- | --- | --- | --- |
| Abdominal pain | Bladder problems (cystitis) | Connective tissue disease | Hair loss |
| Constipation | Chronic kidney disease | Joint pain | Skin color changes |
| Diarrhea | Dehydration | Muscle weakness | Skin pain |
| Dry mouth | Disorders of fluid, electrolyte, and acid-base balance | Musculoskeletal pain | Skin rash |
| Dysgeusia | Kidney injury | Myalgia |  |
| Dyspepsia |  | Osteoarthritis |  |
| Esophageal disorders |  | Spondylopathies |  |
| Gastritis and duodenitis |  | Tendon and synovial disorders |  |
| Gastroesophageal reflux disease |  |  |  |
| Gastrointestinal disorders |  |  |  |
| Gut dysbiosis/microbiome dysbiosis |  |  |  |
| Mouth pain |  |  |  |
| Nausea and vomiting |  |  |  |
| Teeth issues |  |  |  |

| **Metabolic symptoms** | **General symptoms** |
| --- | --- |
| Diabetes mellitus | Appetite |
| Dyslipidemia | Autoimmunity |
| Hormonal disorders | Dizziness |
| Hyperlipidaemia | Fatigue |
| Metabolic disorders | Fever |
| Obesity | Fever/sweats |
| Pancreas injury | Immunosuppression |
| Reproductive hormone deficits | Sore throat |
| Thirst | Swelling |
|  | Unspecified pain |
|  | Viral persistence |

**eTable 3.** Characterization and synonyms of symptoms

Initially, there were ten long COVID subtypes classified based on human body systems and ICD-11 codes. Within each subtype, there are various symptoms. Because there would be discrepancies in symptoms names across studies, we grouped symptoms and their associated synonyms in Supplementary file *Supplementary_Subtypes_and_Symptoms.xlsx.*

**eTable 4.** Quality of life, mortality, and other potential outcome summarized table

| Potential outcome\Number of studies (stratified studies) | Studies from first search | Studies from supplementary search |
| --- | --- | --- |
| Mortality | 8 (0) | 0 (0) |
| Quality of Life | 62 (46) | 3 (1) |
| Disability | 1 (0) | 0 (0) |

**eTable 5.** Summary of included studies

There are 442 studies included in the systematic review, and summary of studies are in supplementary file *Supplementary_Summary.xlsx.*

**eTable 6.** Pooled inverse-variance weighted estimate of the prevalence of long COVID subtypes and symptoms in COVID-19 positive individuals, with corresponding 95% CI obtained by random effects meta-analysis, studies with high risk of bias (both 4/9 and 5/9 scores) removed

| **Subtype/symptom** | **Pooled estimate**  **[95% CI]; (# of studies)** | **Subtype/symptom** | **Pooled estimate**  **[95% CI]; (# of studies)** |
| --- | --- | --- | --- |
| Neurological | 17 [8-32]% (20) | Psychological | 18 [10-31]%; (9) |
| < 1 year | 13 [5-30]%; (13) | < 1 year | 10 [2-31]%; (4) |
| 1 – 2+ years | 27 [15-44]%; (7) | 1 – 2+ years | 29 [14-51]%; (5) |
| Concentration/confusion/brainfog | 4 [3-5]%; (24) | Anxiety | 5 [3-7]%; (22) |
| Headache | 5 [4-7]%; (53) | Depression | 6 [4-10]%; (13) |
| Malaise | 8 [3-20]%; (5) | Insomnia | 6 [4-10]%; (25) |
| Memory problems | 8 [5-11]%; (9) | Mood swings | 6 [2-15]%; (5) |
| Sleep problems | 5 [3-9]%; (11) | PTSD symptoms | 14 [2-52]%; (6) |
| Smell | 5 [3-6] %; (36) |  |  |
| Smell or Taste | 5 [3-10]%; (10) |  |  |
| Taste | 3 [1-5]%; (25) |  |  |
| Tinnitus | 1 [1-2]%; (16) |  |  |
| Tremors/Chills | 1 [0-5]%; (9) |  |  |
| Vision problems | 2 [1-4]%; (9) |  |  |
| Cardiovascular | 11 [4-31]%; (12) | Respiratory | 21 [14-31]%; (24) |
| < 1 year | 8 [2-29]%; (8) | < 1 year | 22 [14-32]%; (19) |
| 1 – 2+ years | 23 [13-37]%; (4) | 1 – 2+ years | 21 [14-28]%; (5) |
| Arrhythmia | 2 [0-17]%; (3) | Breathlessness | 6 [1-29]%; (3) |
| Hypertension | 3 [1-7]%; (4) | Chest pain | 3 [3-4]%; (45) |
| Palpitations | 3 [2-4]%; (29) | Chest tightness | 2 [1-3]%; (7) |
| Tachycardia | 2 [1-6]%; (8) | Cough | 5 [4-6]%; (42) |
|  |  | Dyspnea | 7 [5-10]%; (25) |
|  |  | Nasal congestion | 2 [1-4]%; (10) |
| Musculoskeletal | 9 [5-16]%; (11) | Dermatological | 13 [8-20]%; (7) |
| < 1 year | 8 [4-13]%; (10) | < 1 year | 14 [7-25]%; (4) |
| 1 – 2+ years | 30 [29-32]%; (1) | 1 – 2+ years | 12 [5-24]%; (3) |
| Joint pain | 6 [3-10]%; (13) | Hair loss | 4 [3-5]%; (21) |
| Muscle weakness | 8 [3-22]%; (2) | Skin rash | 2 [1-3]%; (18) |
| Myalgia | 5 [3-7]%; (19) |  |  |
| Gastrointestinal | 5 [4-6]%; (29) | General Fatigue | 19 [16-22]%; (102) |
| < 1 year | 6 [5-7]%; (23) | < 1 year | 17 [15-21]%; (80) |
| 1 – 2+ years | 4 [1-11]%; (6) | 1 – 2+ years | 25 [19-33]%; (22) |
| Abdominal pain | 1 [1-2]%; (13) | Loss of appetite | 2 [1-3]%; (25) |
| Constipation | 1 [0-3]%; (5) | Dizziness | 3 [2-4]%; (32) |
| Diarrhea | 1 [1-3]%; (10) | Fever | 2 [1-3]%; (35) |
| Stomach pain | 1 [0-7]%; (2) | Sore throat | 3 [2-5]%; (22) |
|  |  | Sweats | 2 [1-4]%; (9) |

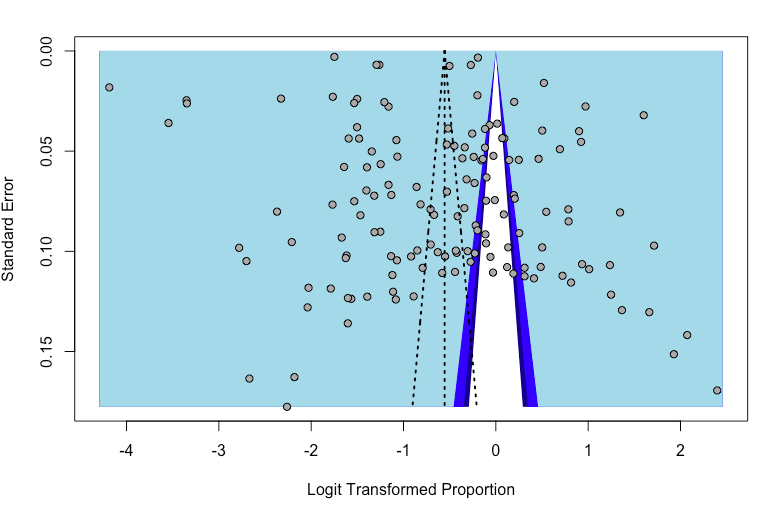

**eFigure 1.** Funnel plot for publication bias assessment among studies used to estimate the pooled long COVID prevalence in COVID-19 positive individuals

A funnel plot based on 144 studies was created to assess publication bias. Upon inspecting the plot, asymmetry may be present. Using Egger’s linear regression test and Begg’s rank correlation test for asymmetry, the conclusion that publication bias does not appear to be of concern was reached. Egger’s test-statistic 1.72 is not significant (p-value = 0.0873), and Begg’s test result -1.11 is also not significant (p-value 0.2660).

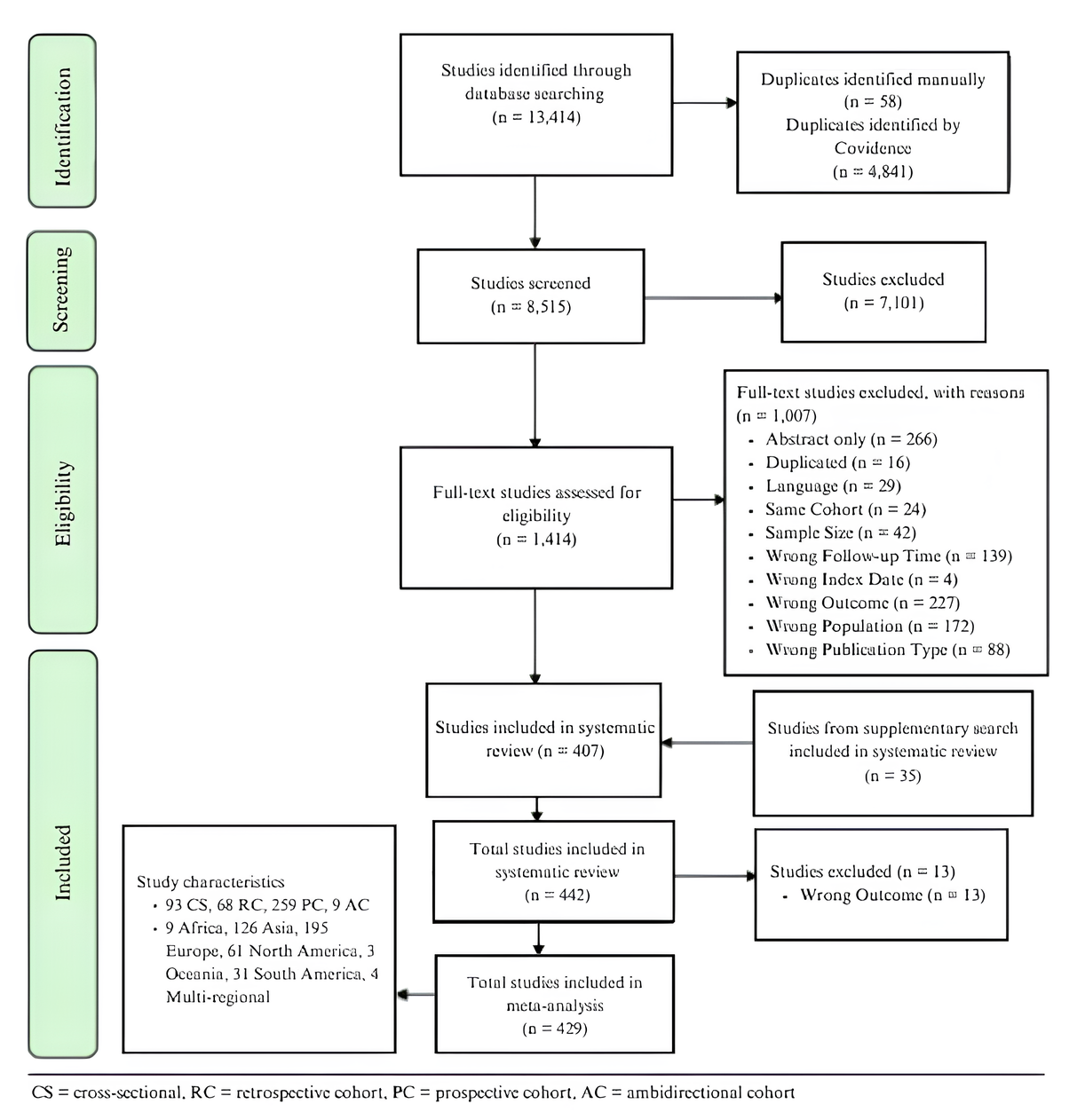

**eFigure 2.** PRISMA flow diagram

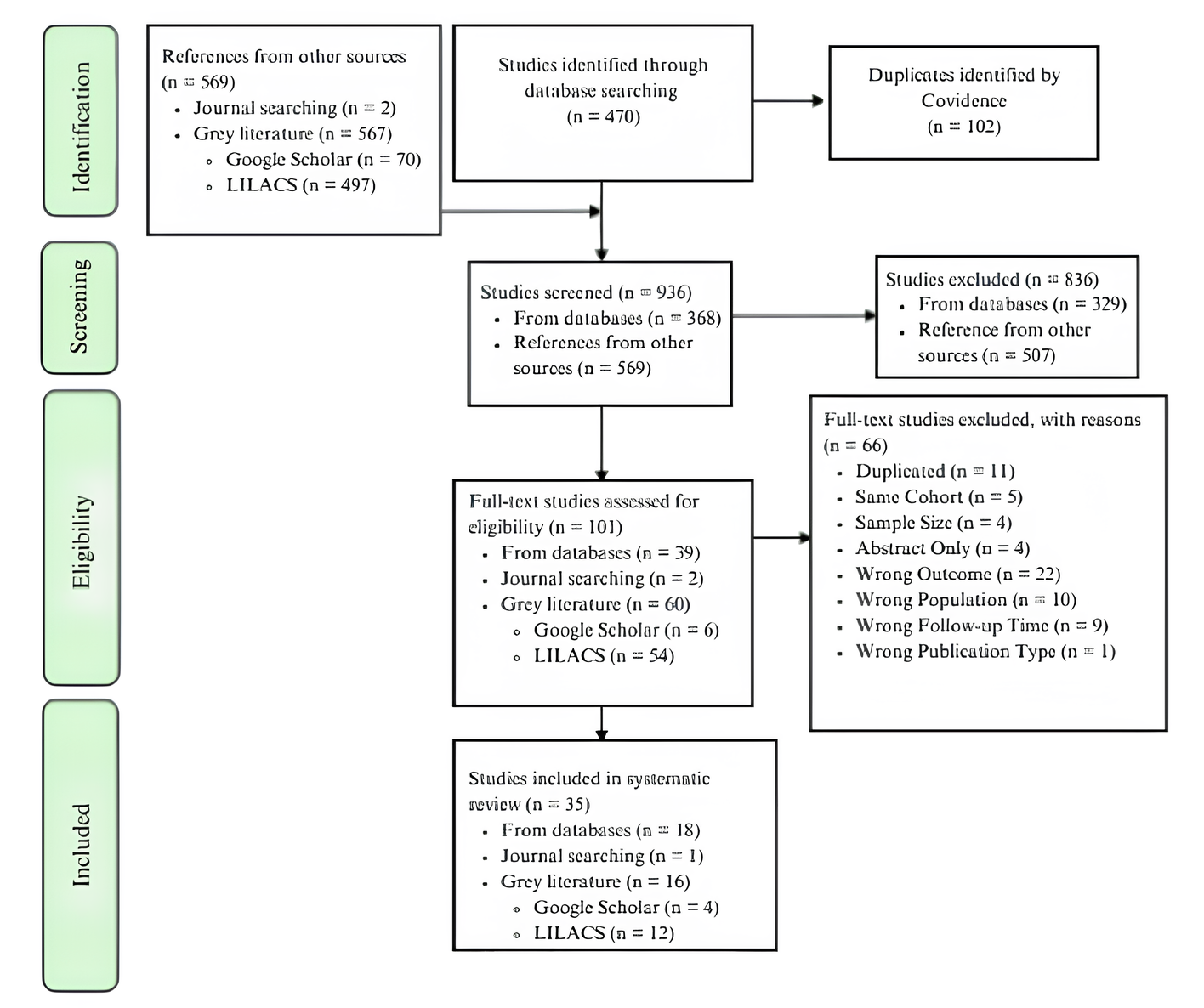

**eFigure 3.** PRISMA flow diagram for supplementary search

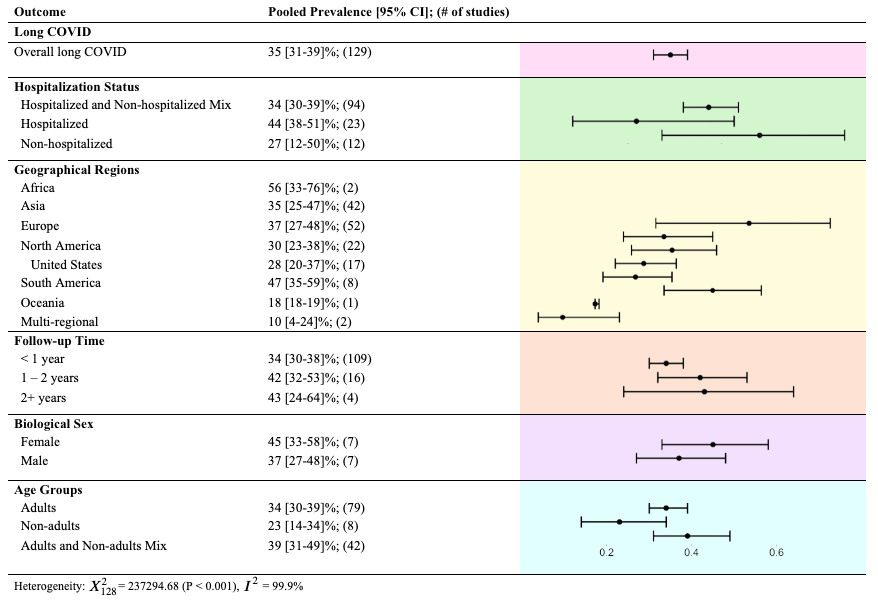

**eFigure 4.** Forest plot for pooled long COVID prevalence in COVID-19 positive individuals, corresponding 95% confidence intervals and number of contributing studies stratified by hospitalization status, geographic region, follow-up time, biological sex, and age-group. Studies with high risk of bias (both 4/9 and 5/9 scores) removed.

**eFigure 5.** Forest plots with the prevalence estimates of long COVID in COVID-19 positive individuals stratified by publication year

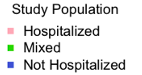

Publication year 2021 - 2024

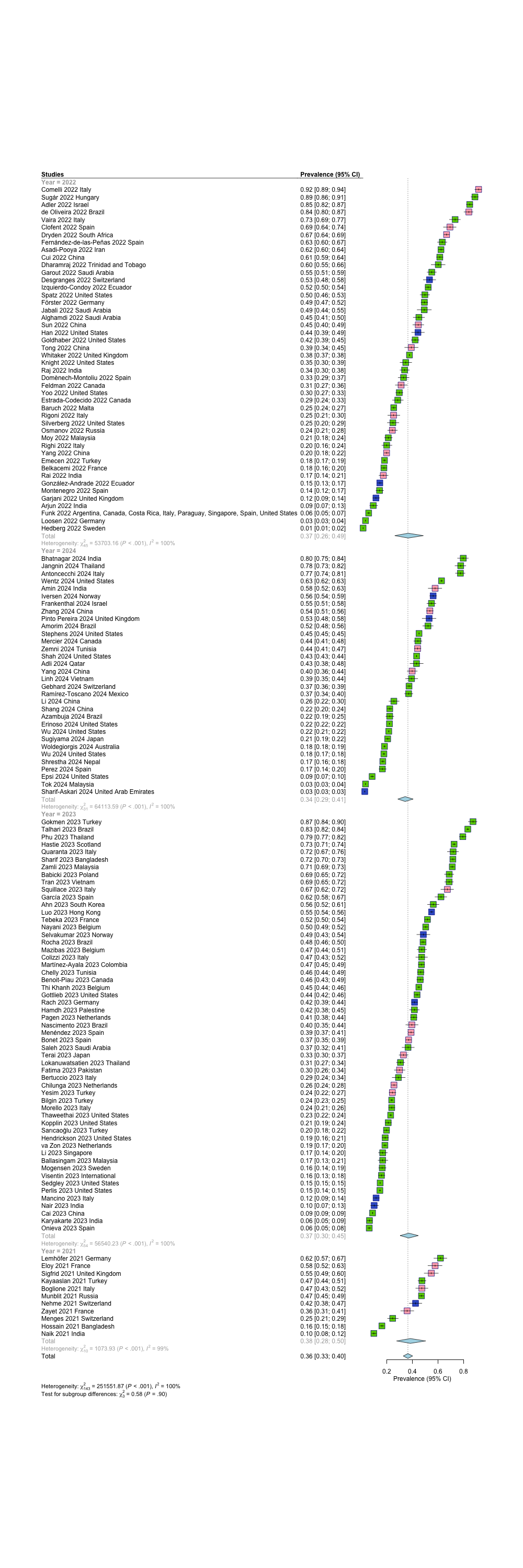

**eFigure 6.** Forest plots with the prevalence estimates of long COVID in COVID-19 positive individuals stratified by follow-up time and hospitalization status.

Follow-up time < 1 year:

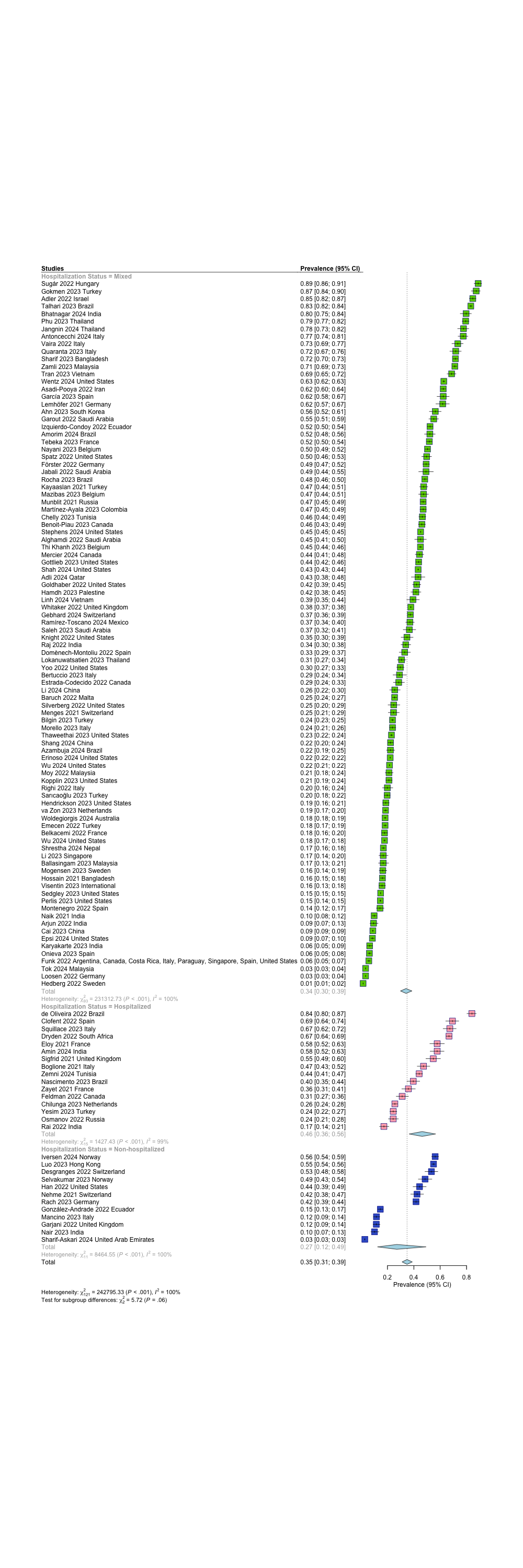

Follow-up time 1 – 2 years:

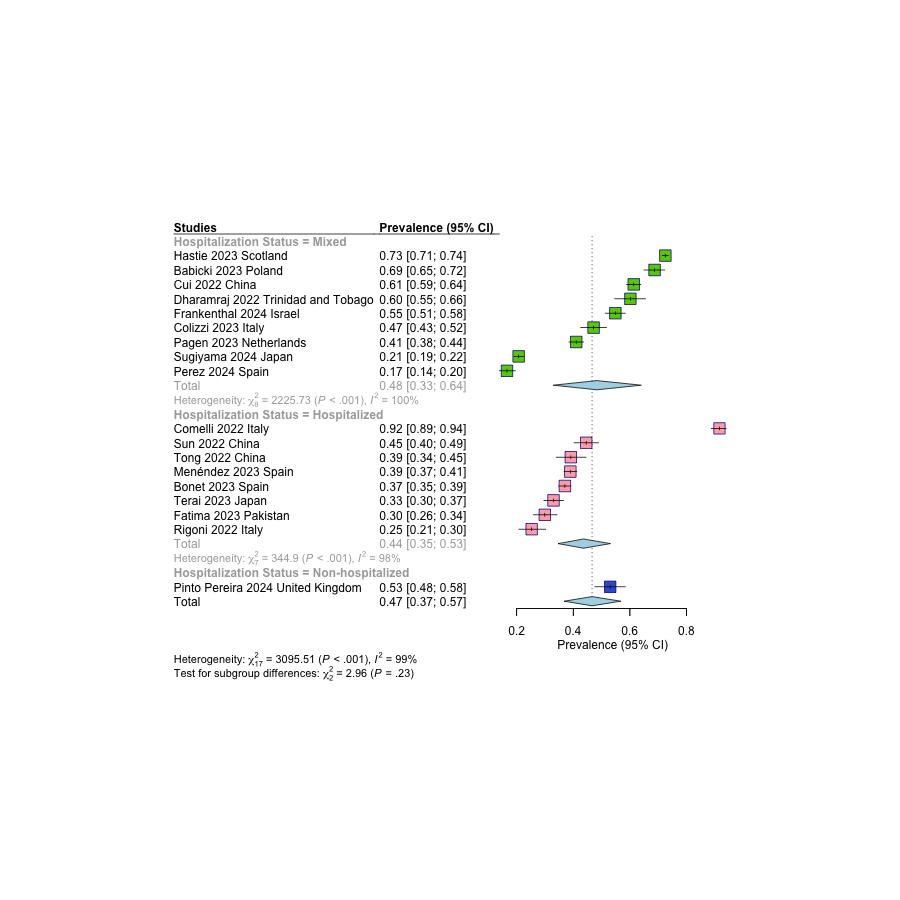

Follow-up time 2+ years:

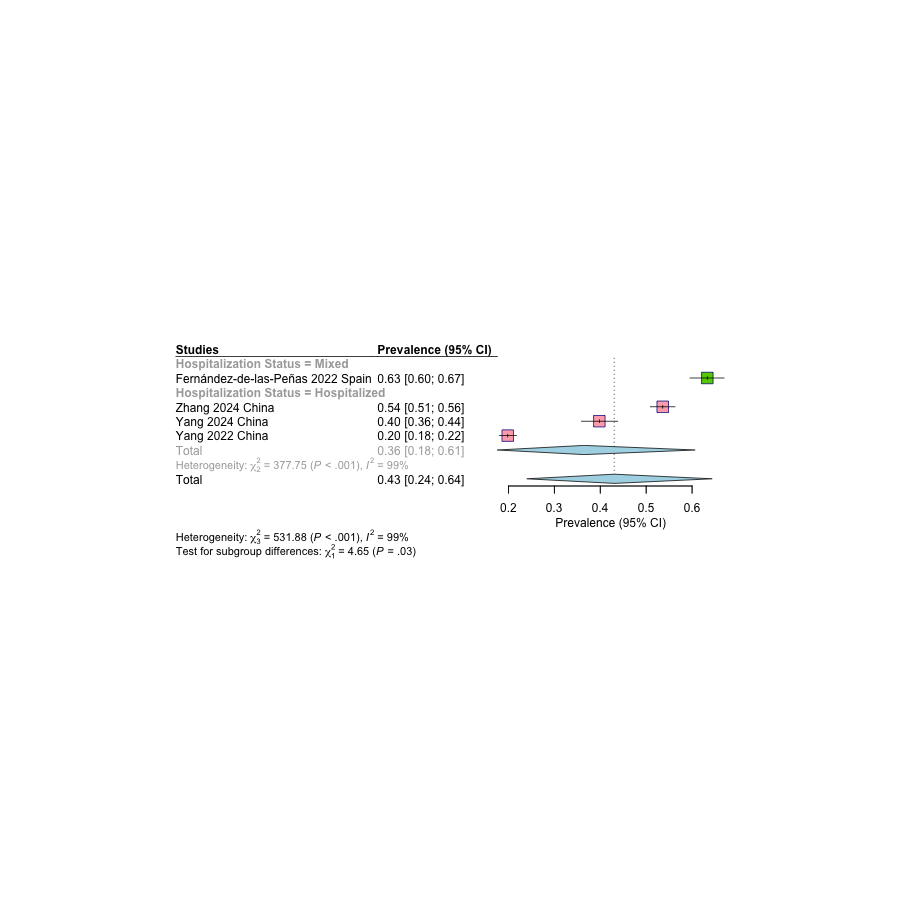

**eFigure 7.** Forest plots with the prevalence estimates of long COVID in COVID-19 positive individuals stratified by A) Biological sex, B) Geographical regions, C) Hospitalization status, D) Follow-up time, E) Age groups

A) Biological sex

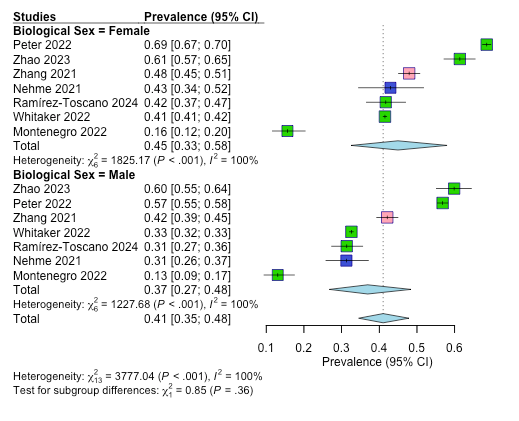

B) Geographical regions

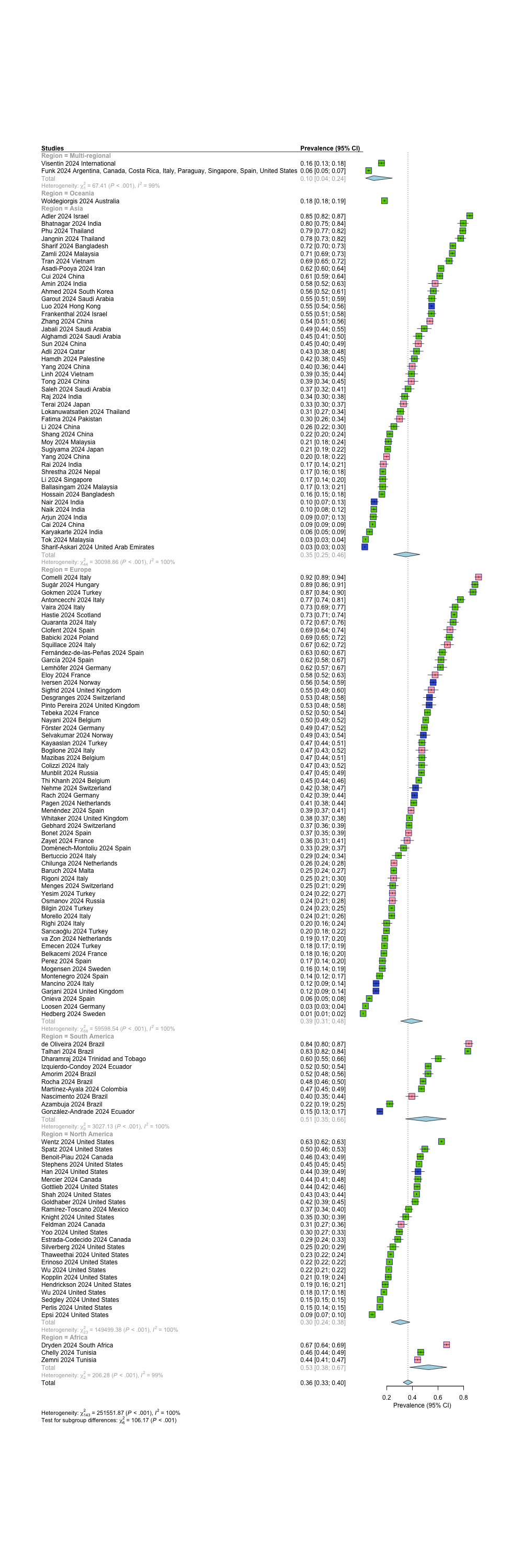

United States

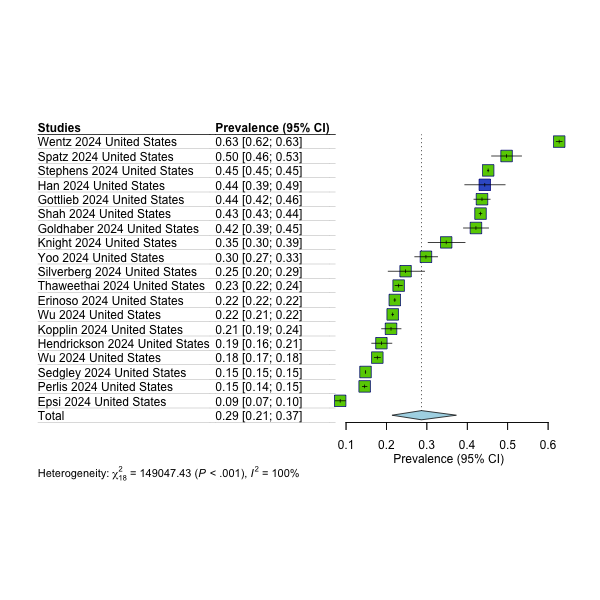

C) Hospitalization status

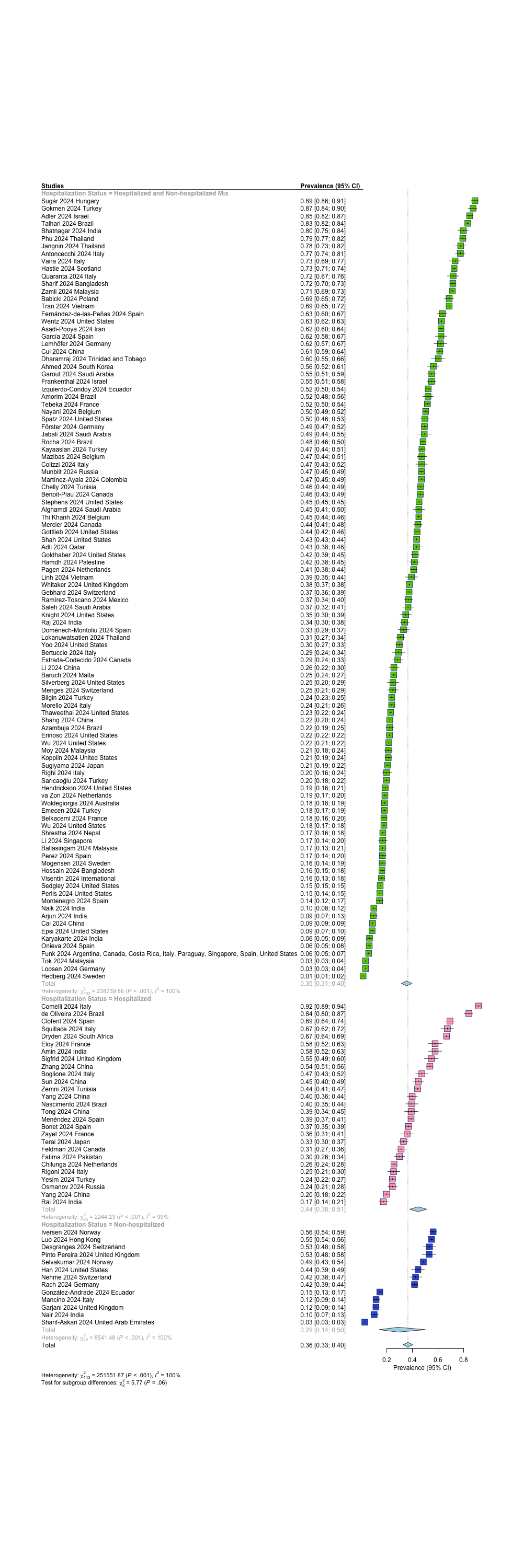

D) Follow-up time

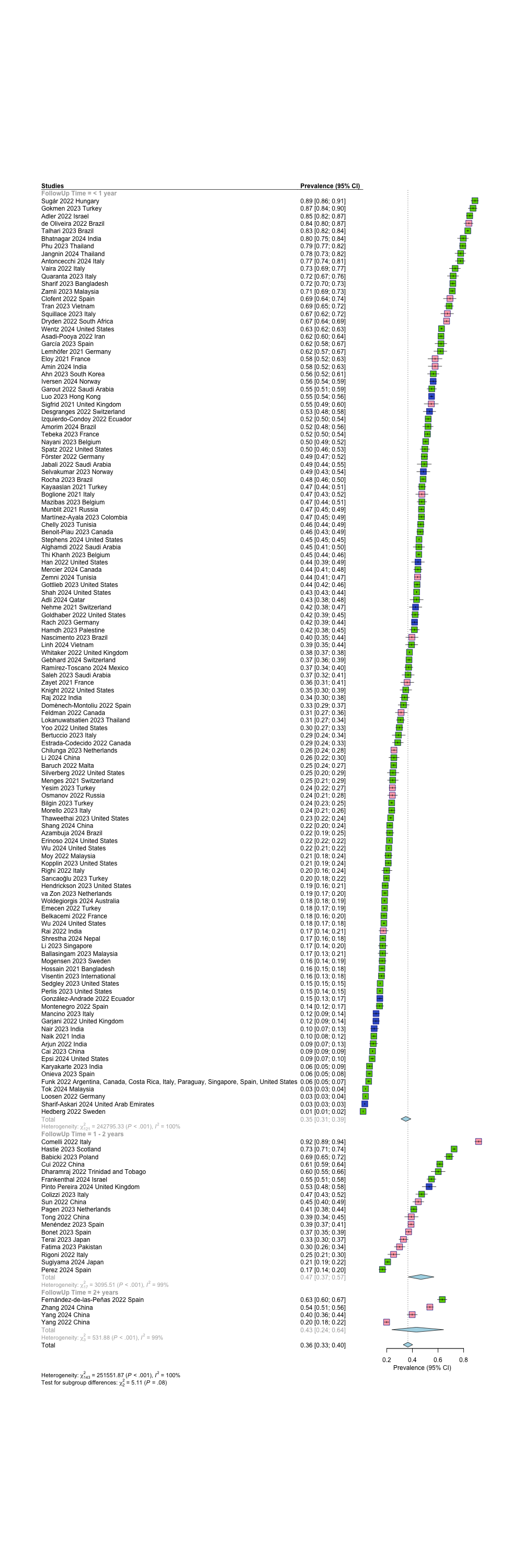

E) Age, categorized by adults, non-adults, or all-age

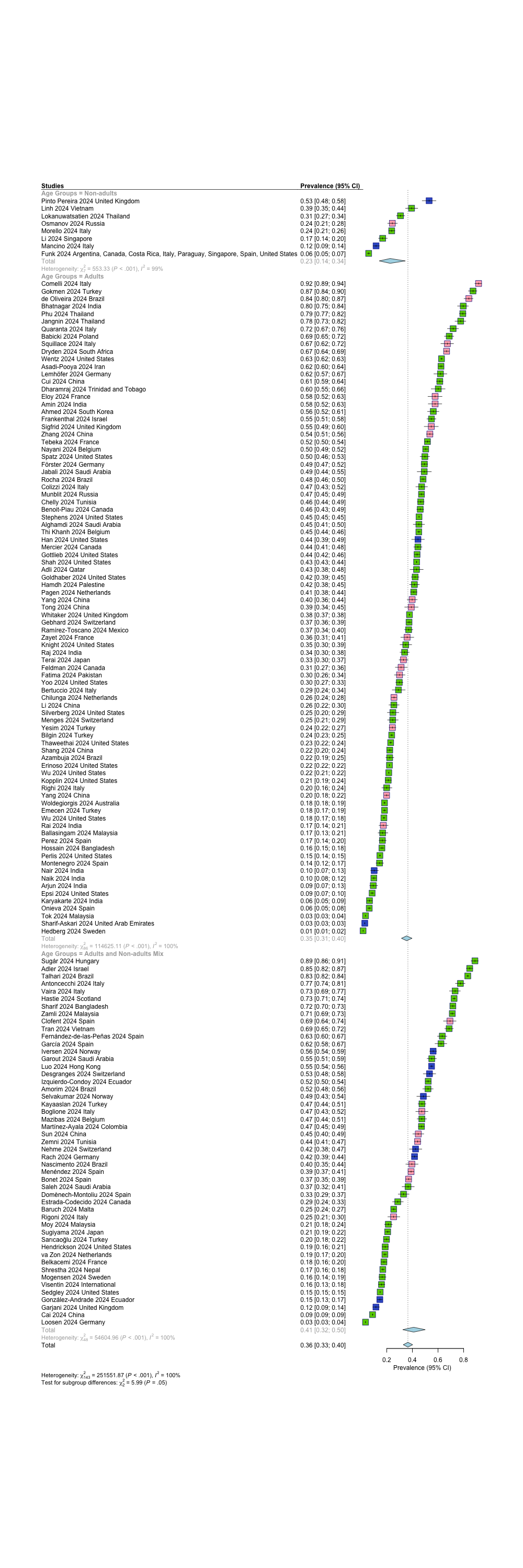

**eFigure 8.** Supplementary meta-analysis of long COVID condition subtype-specific symptom-specific prevalence in COVID-19 positive individuals

Neurological Subtype

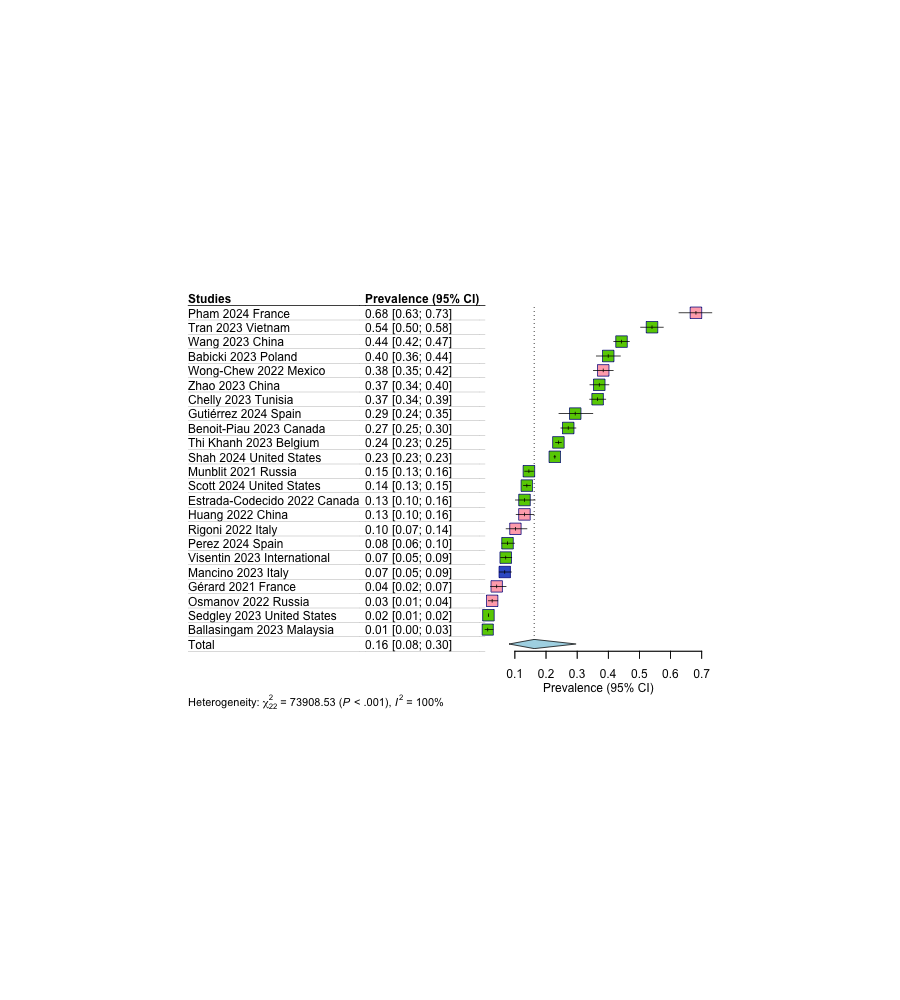

Neurological Follow-up

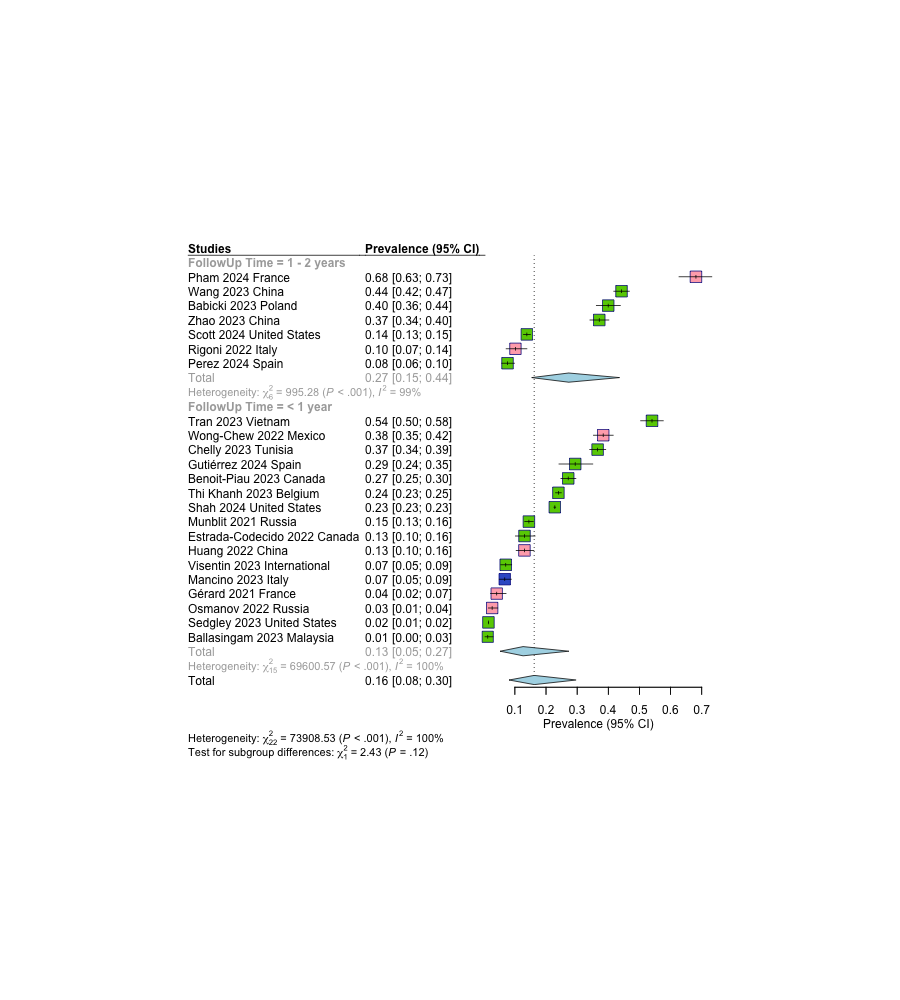

Concentration/confusion/brain

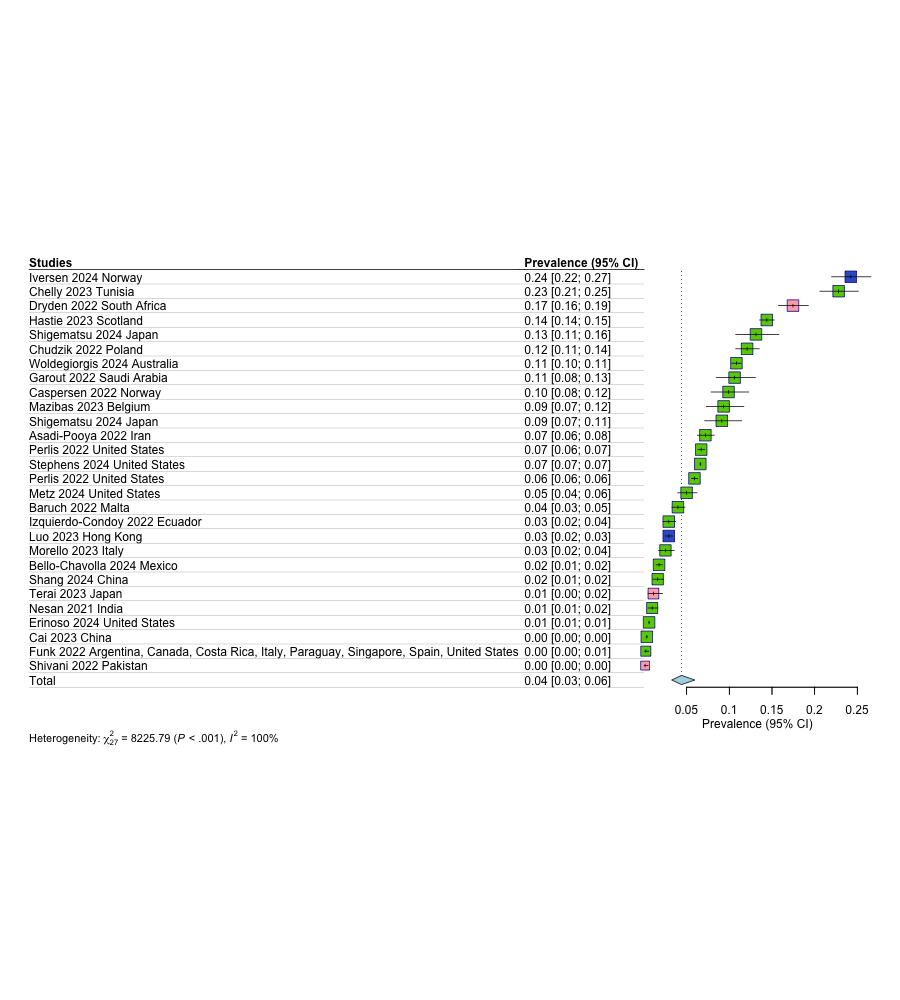

Headache

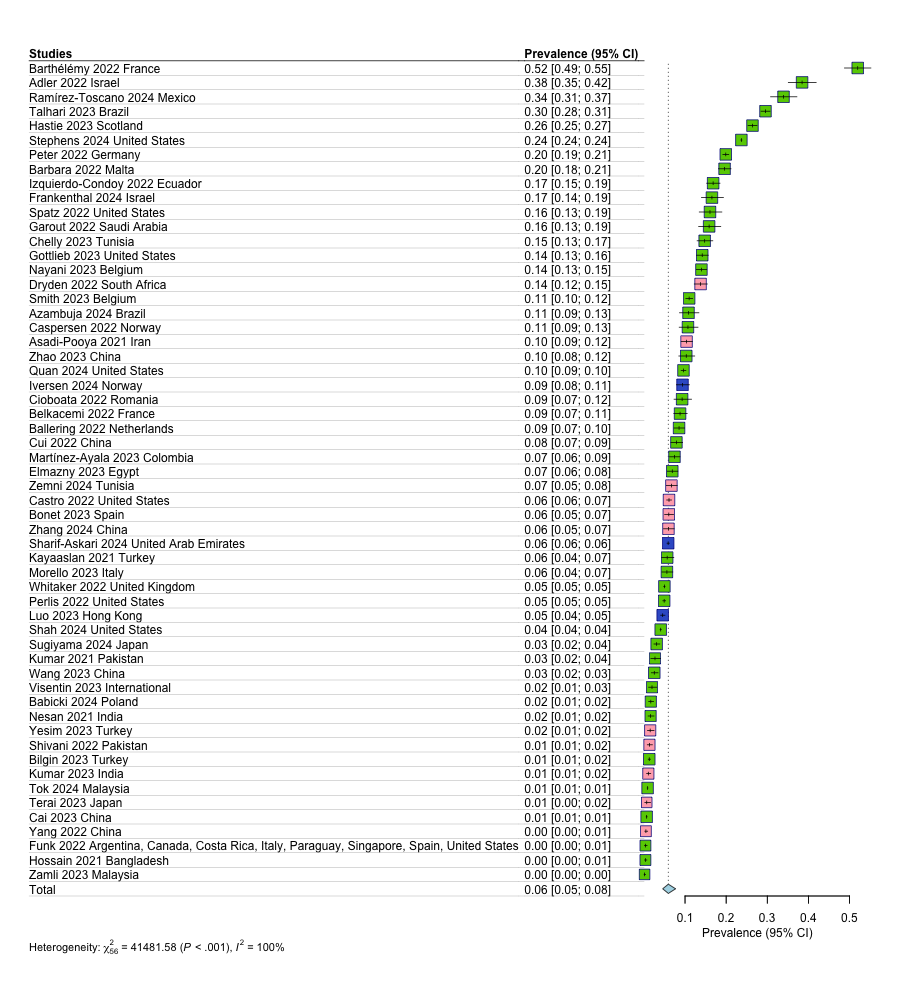

Malaise

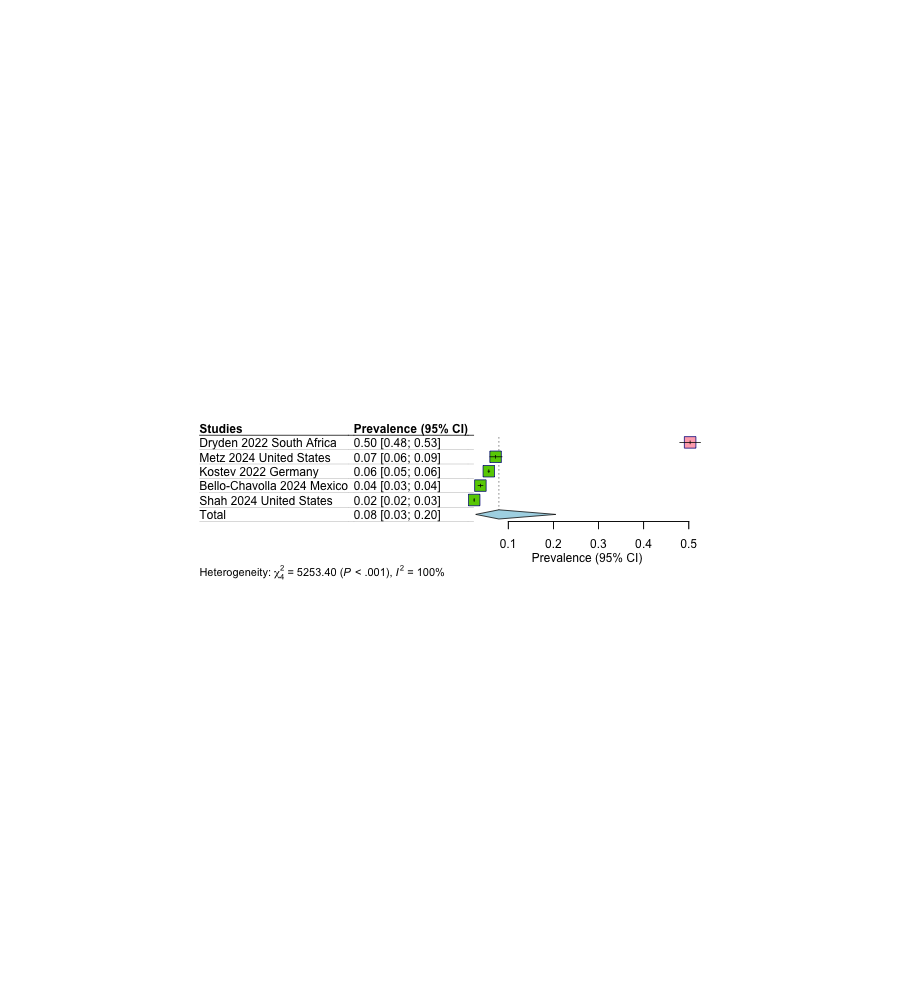

Memory problems

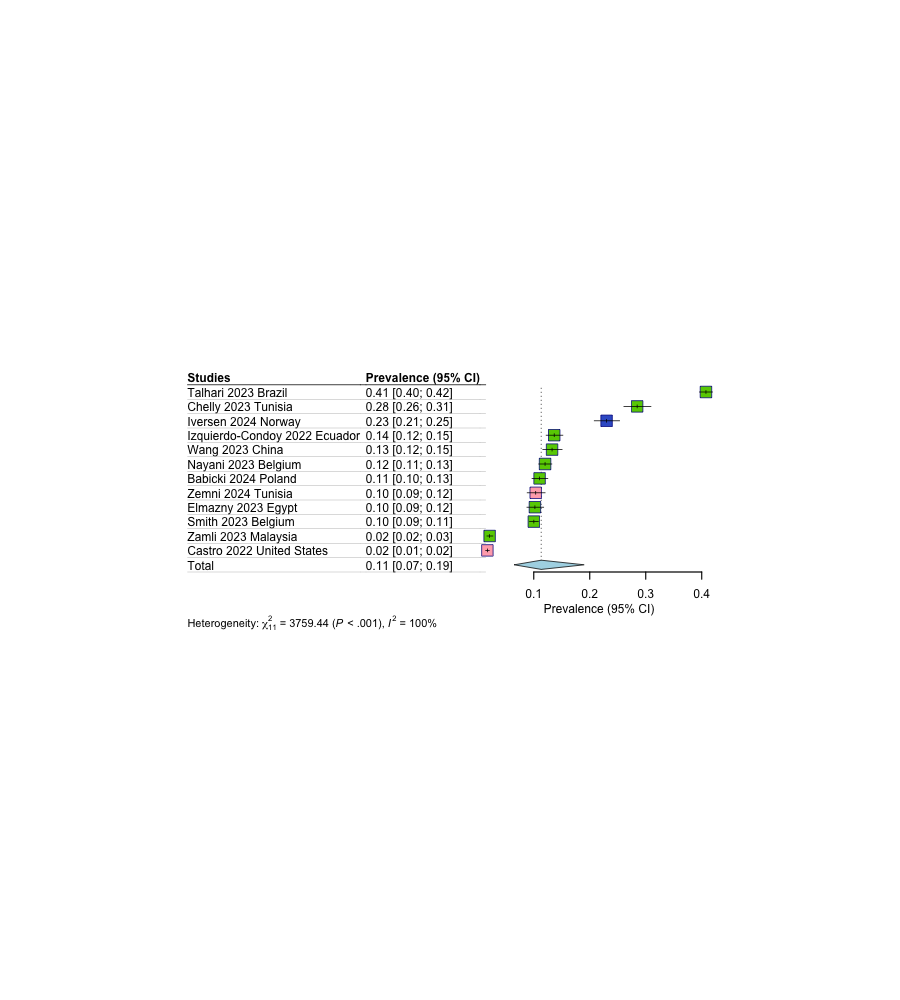

Sleep problems

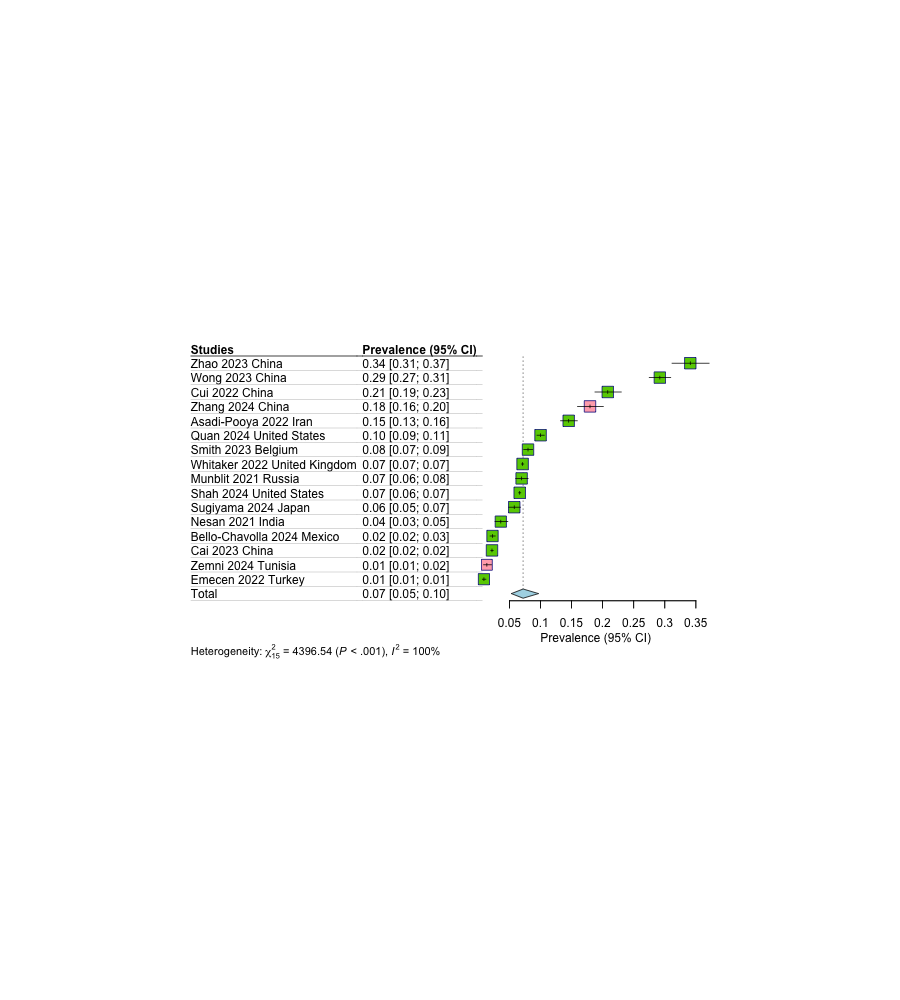

Smell

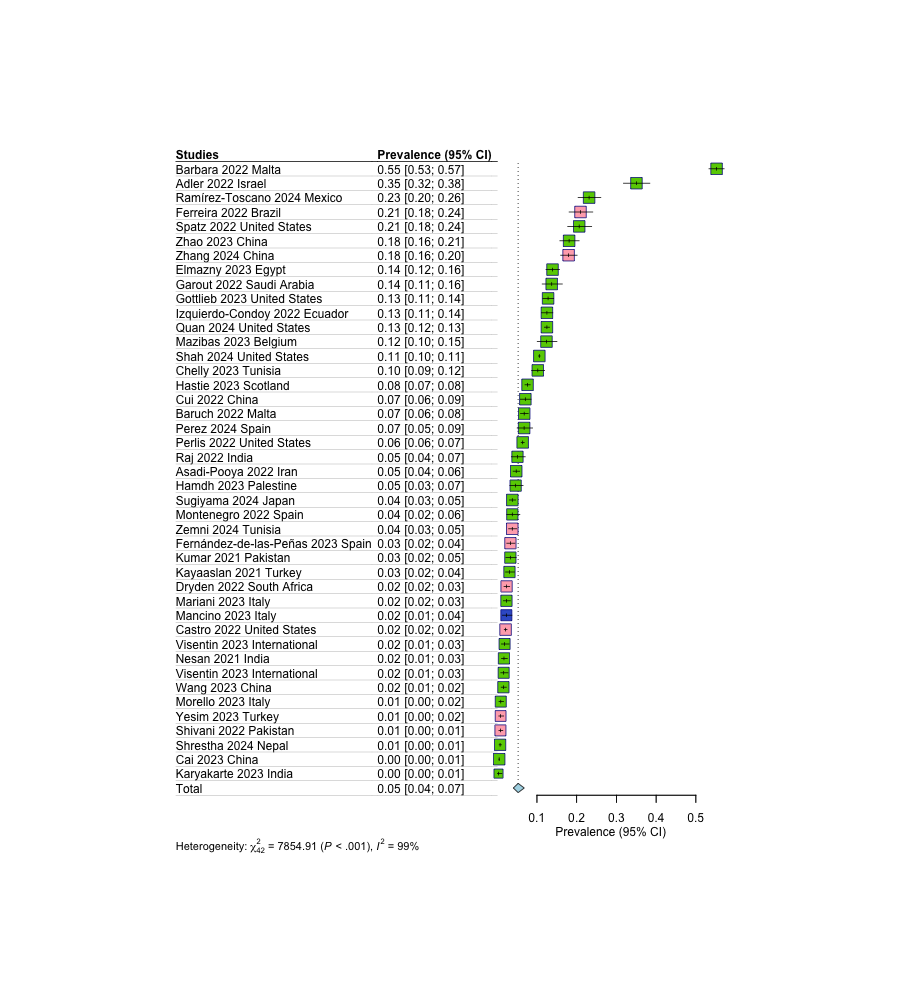

Smell or Taste

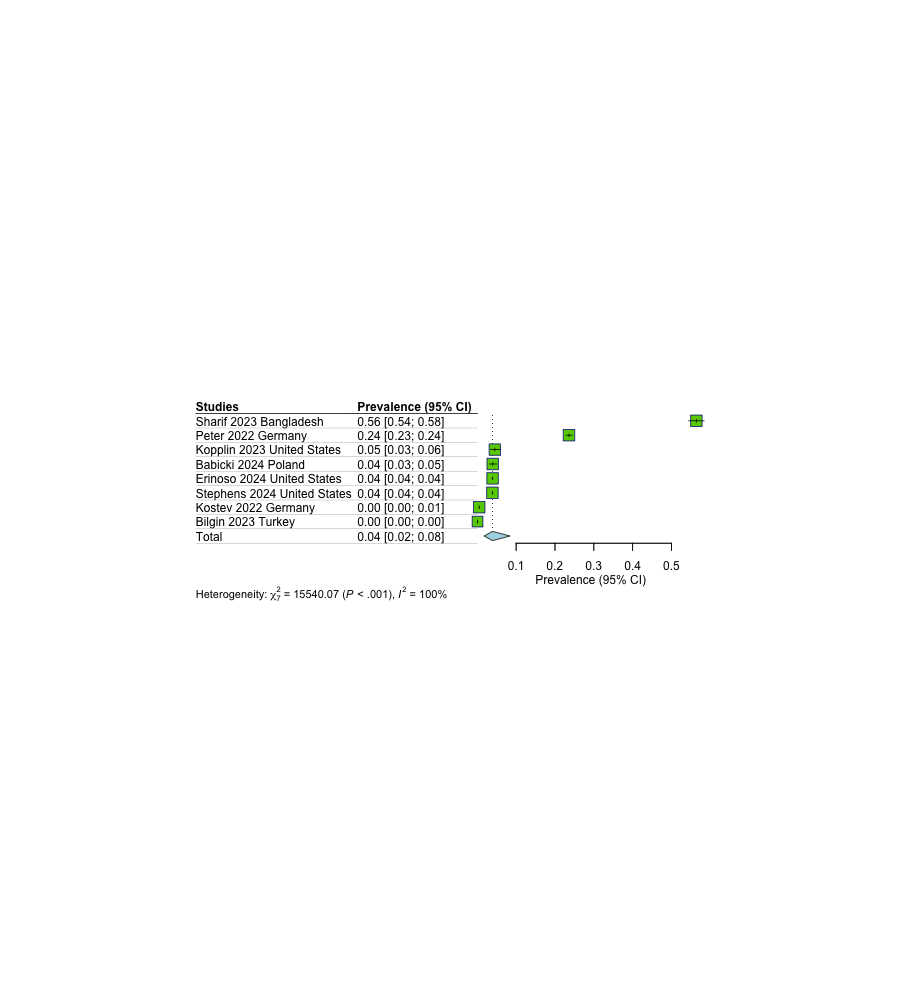

Taste

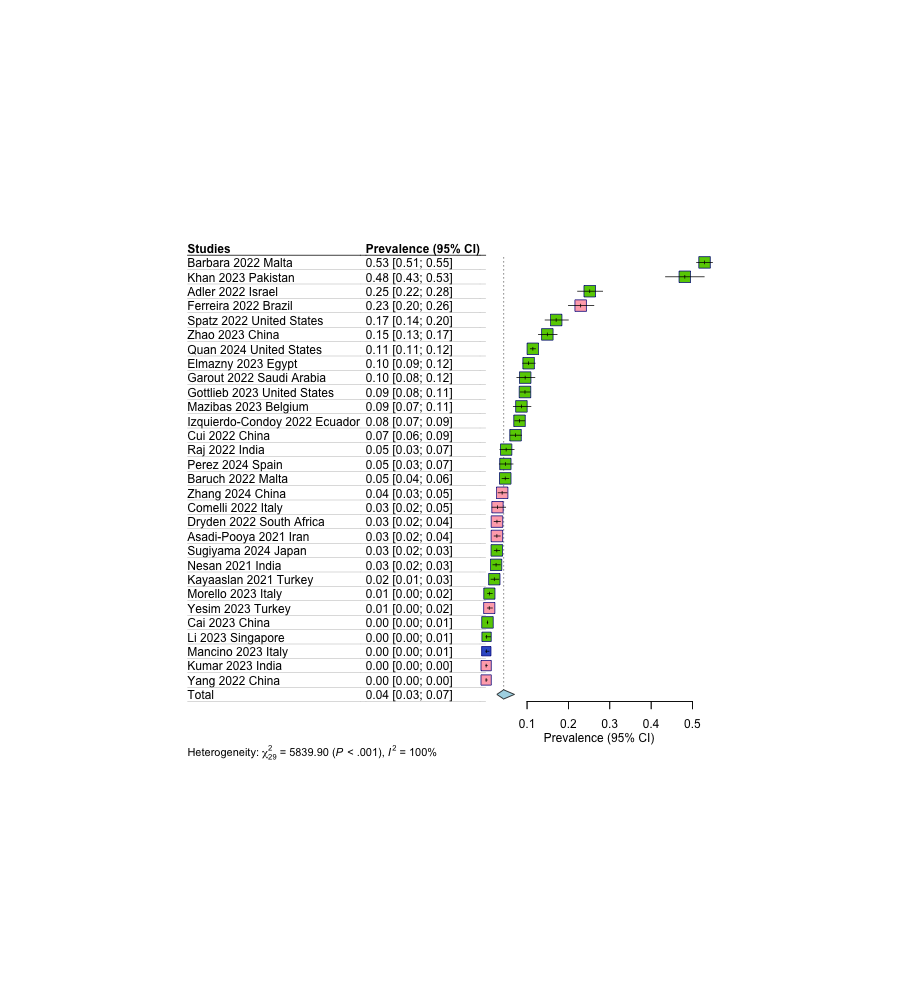

Tinnitus

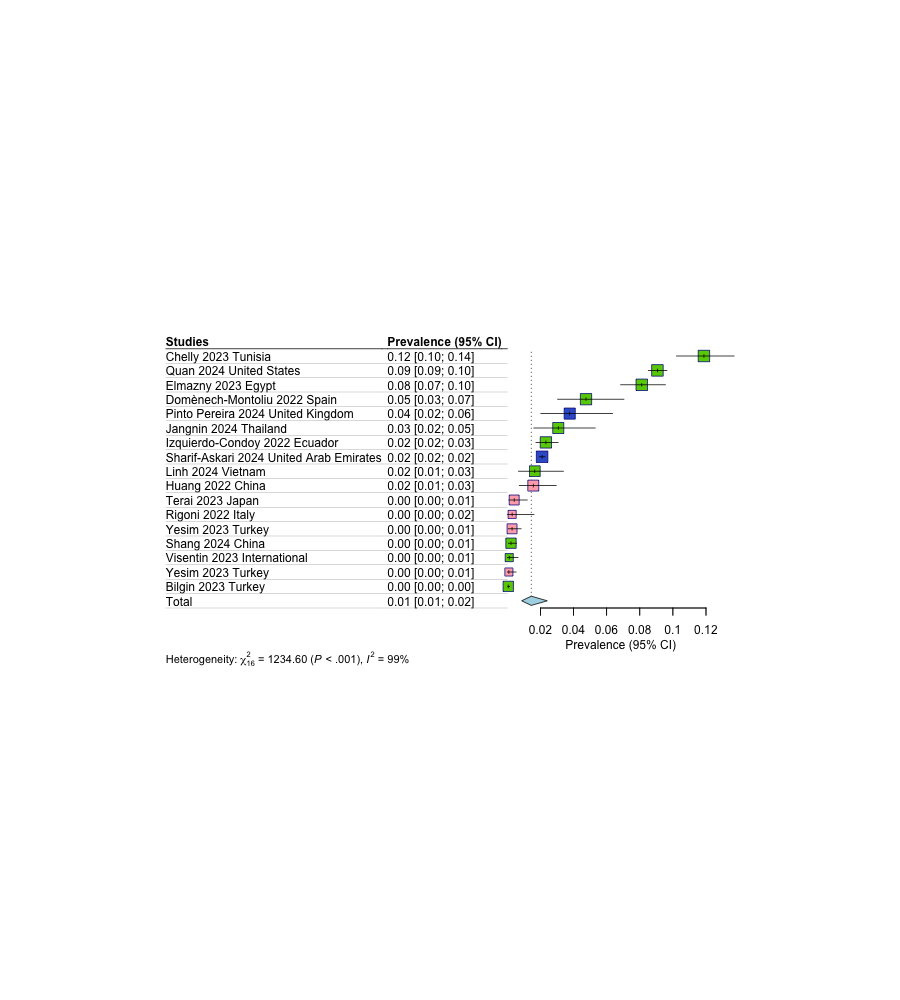

Tremors/Chills

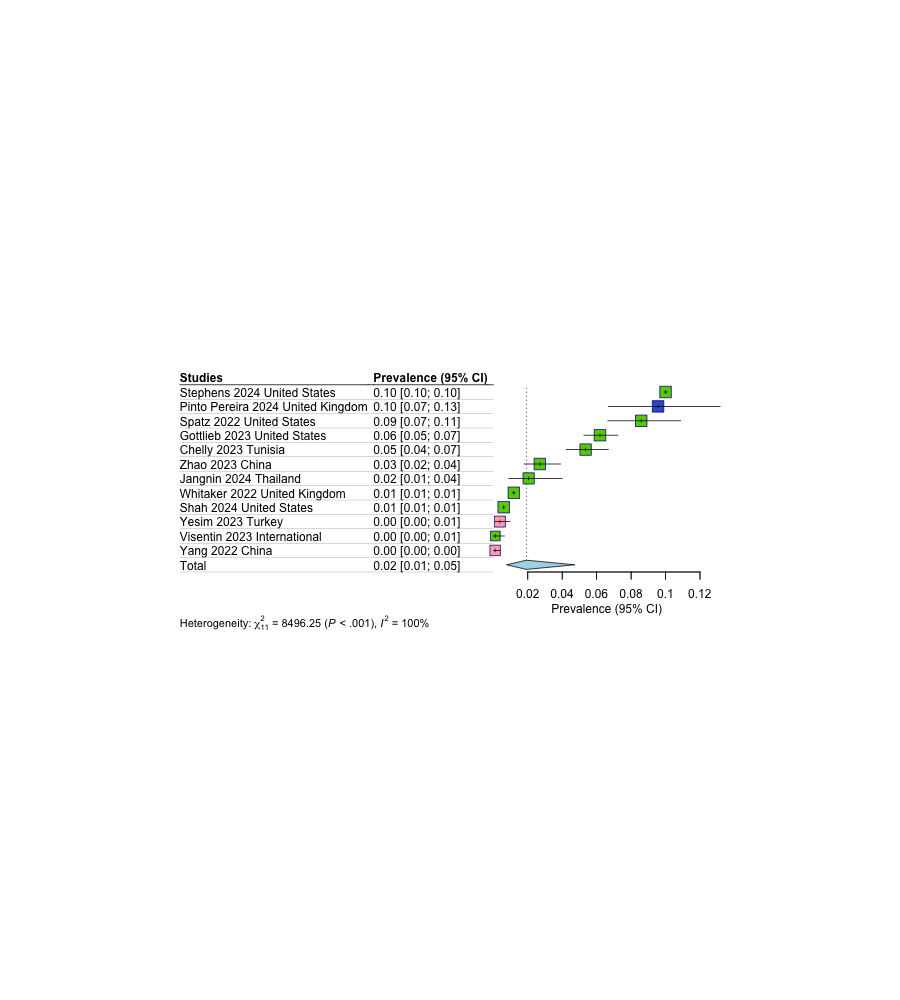

Vision problems

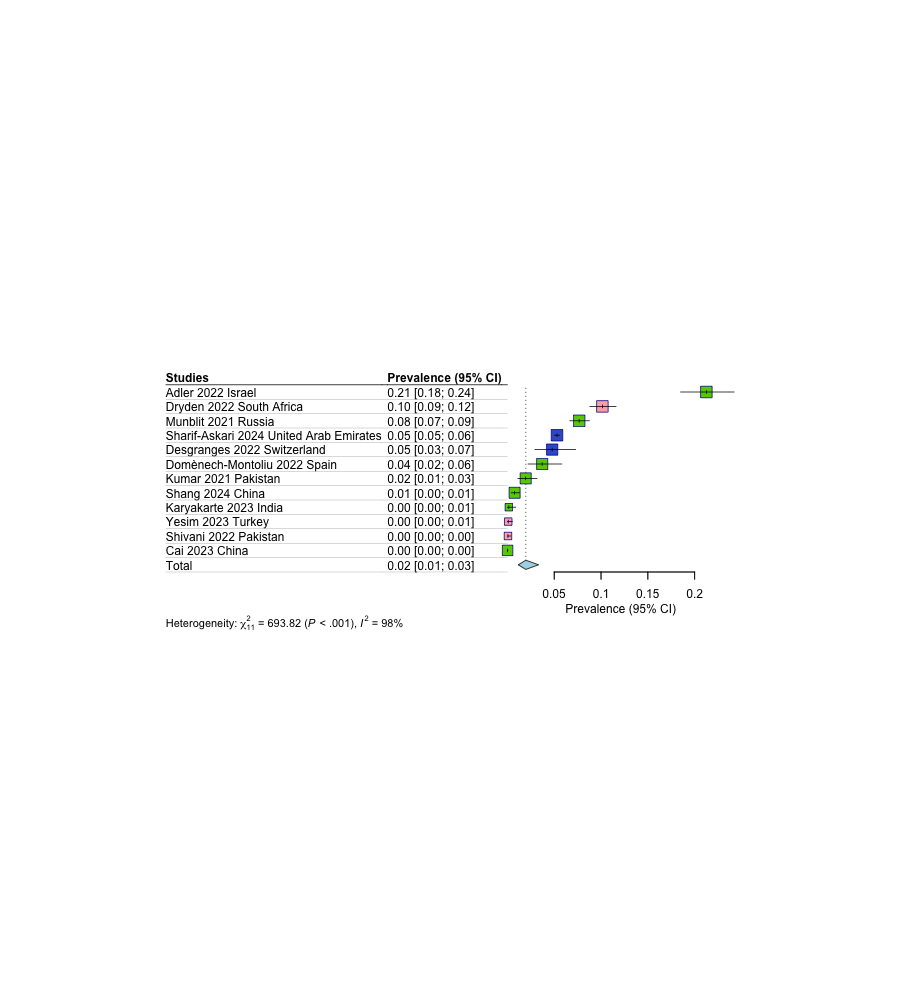

Psychological Subtype

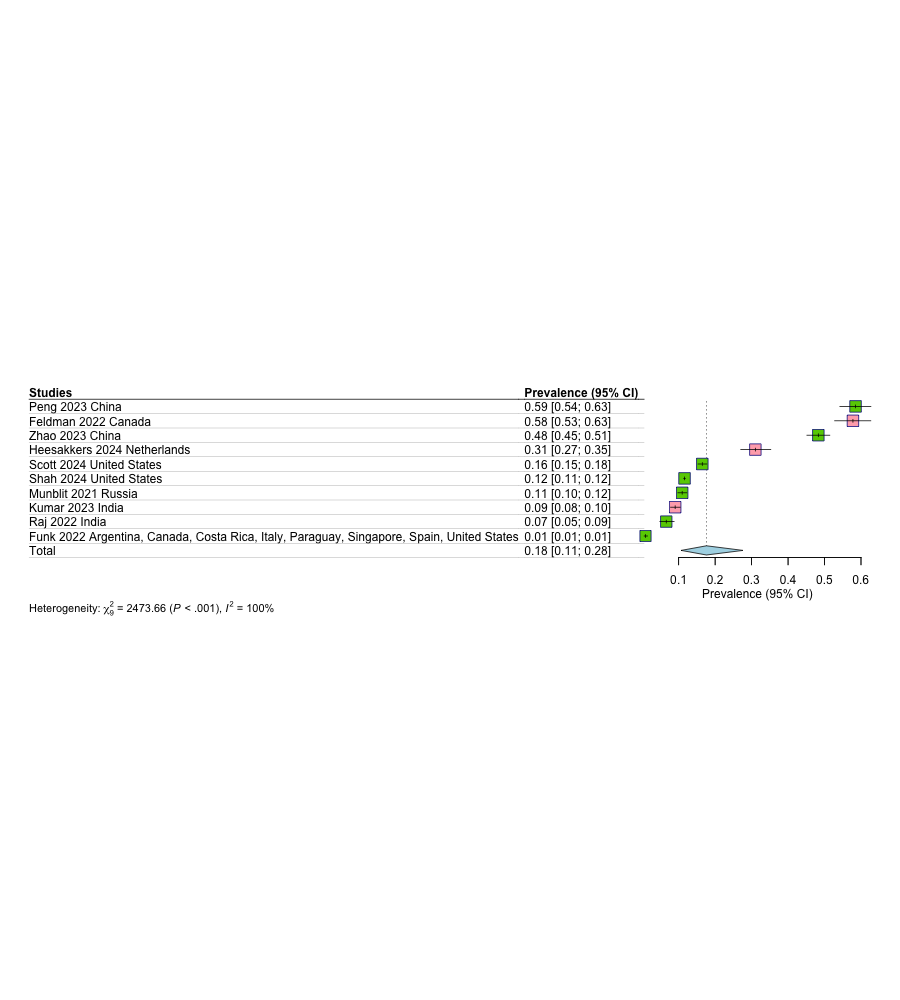

Psychological Follow-up

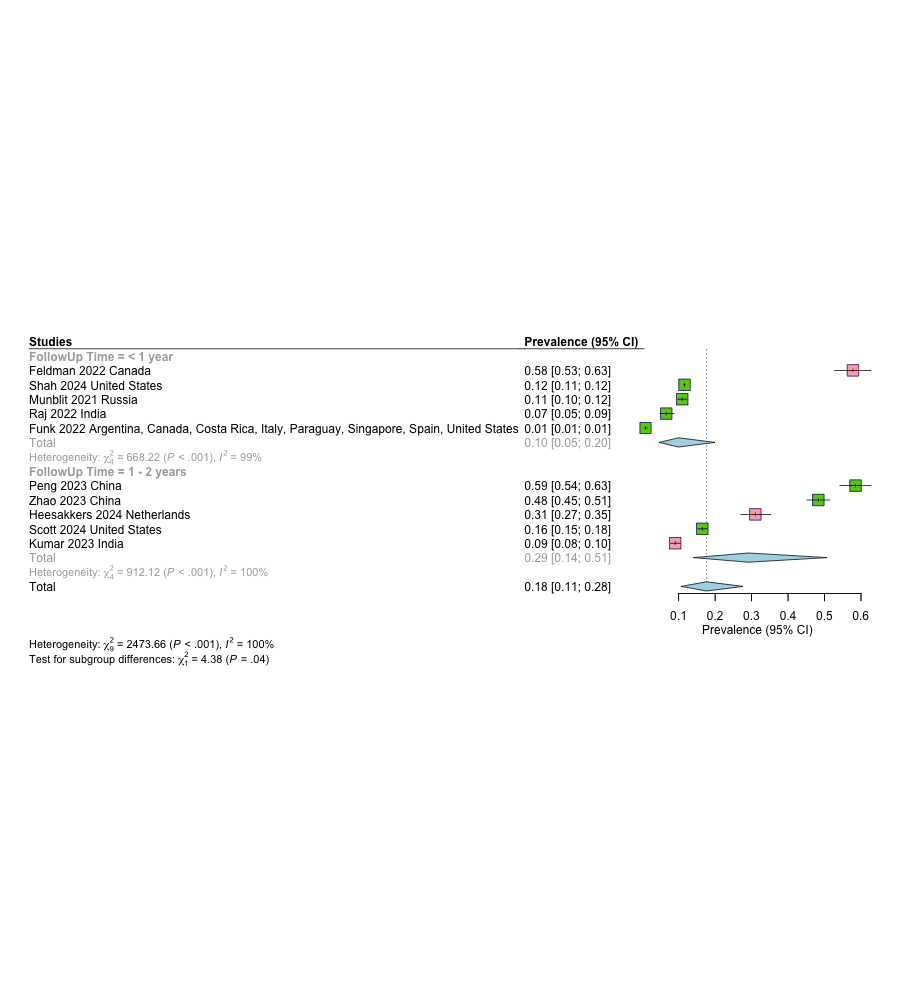

Anxiety

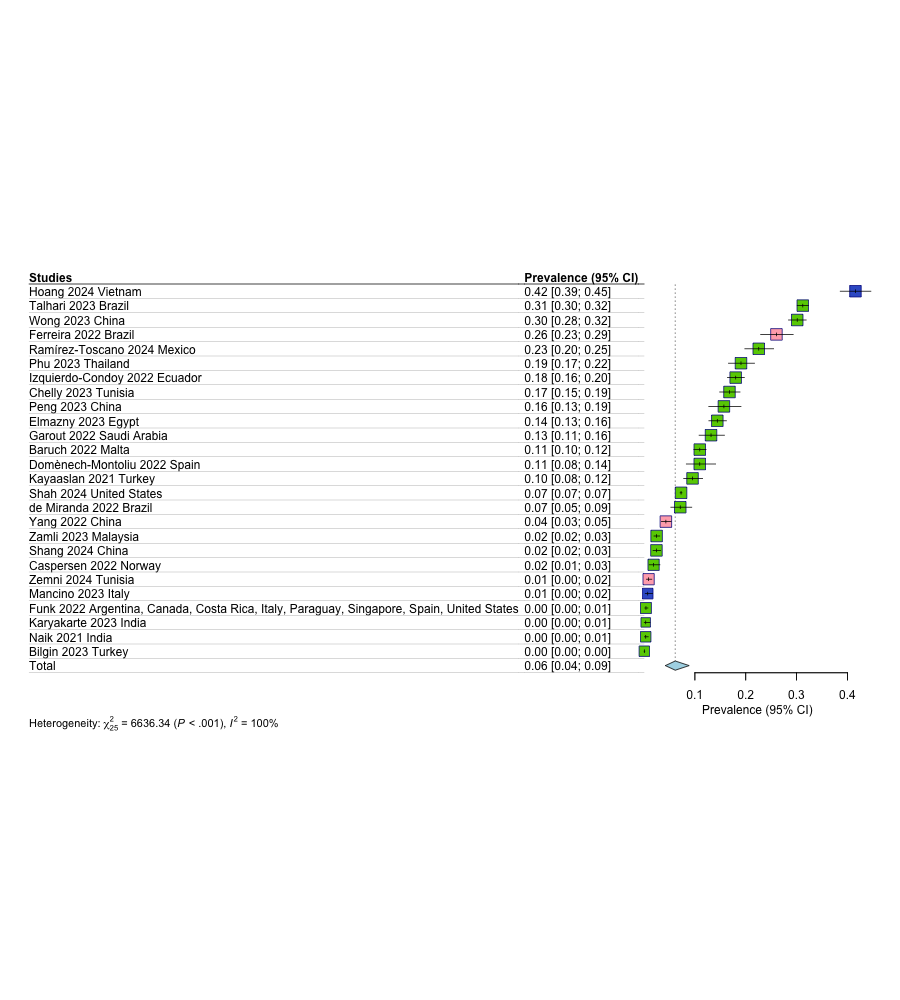

Depression
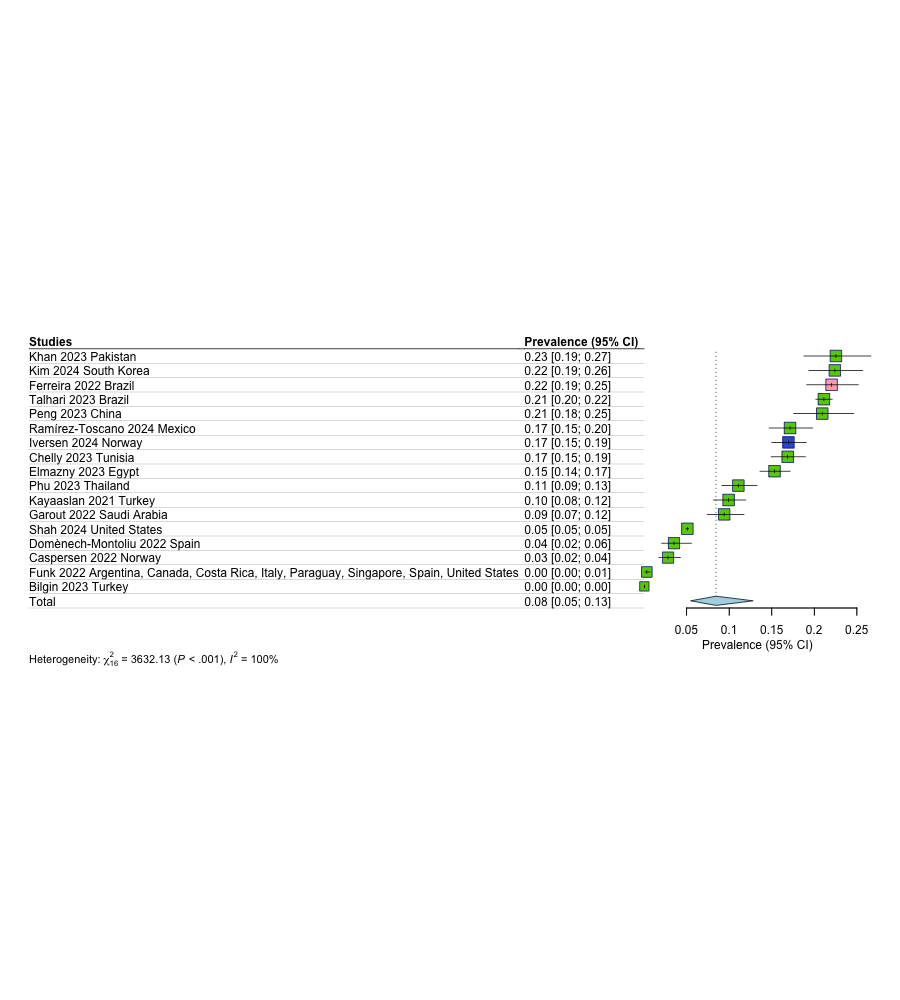

Insomnia

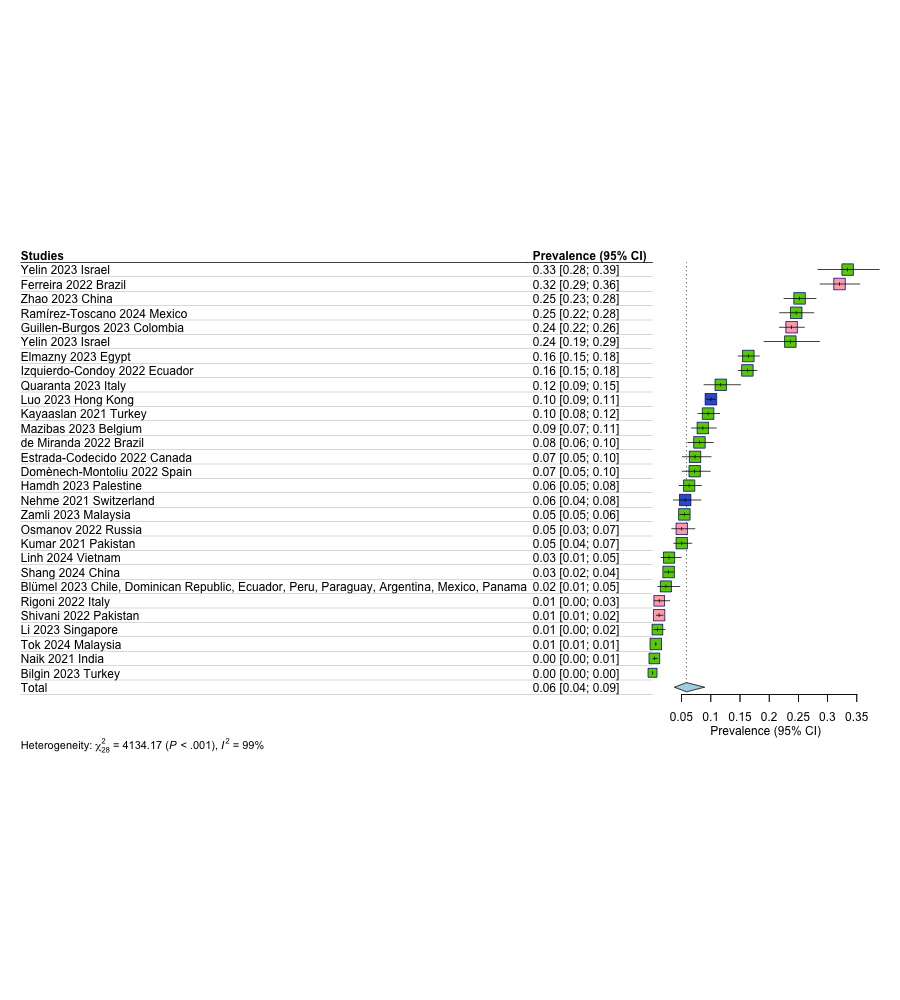

Mood swings

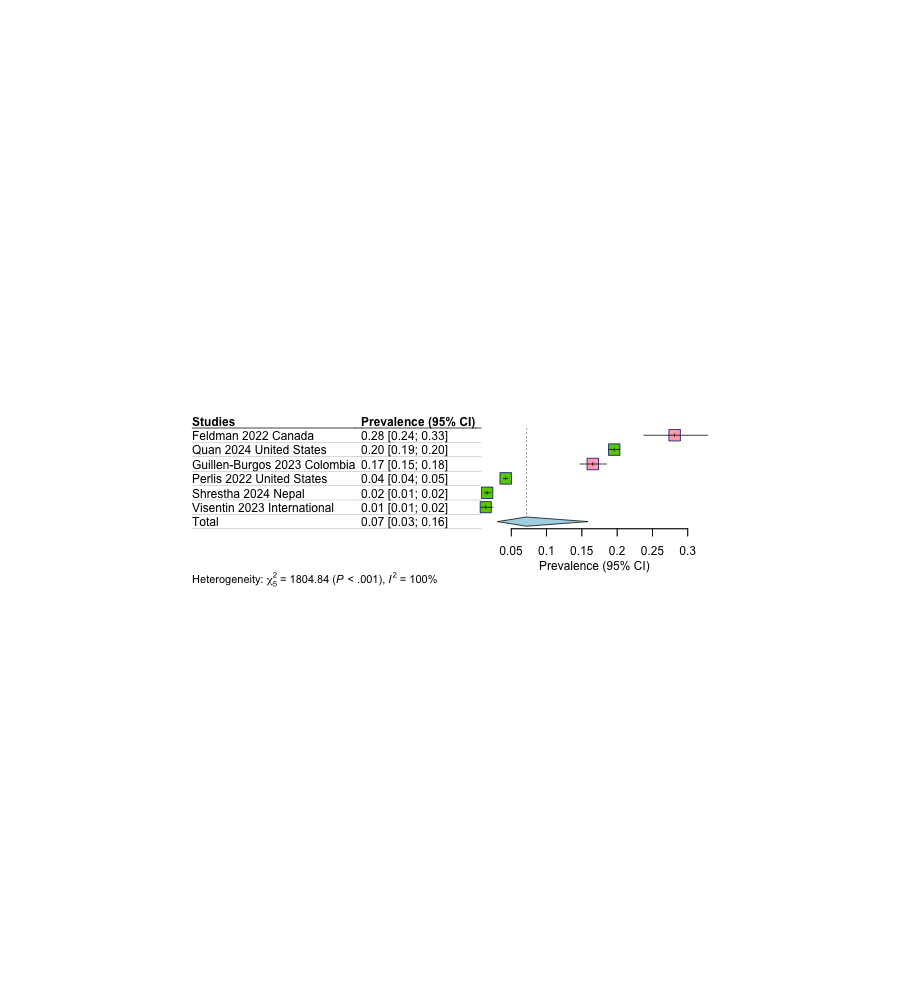

PTSD symptoms

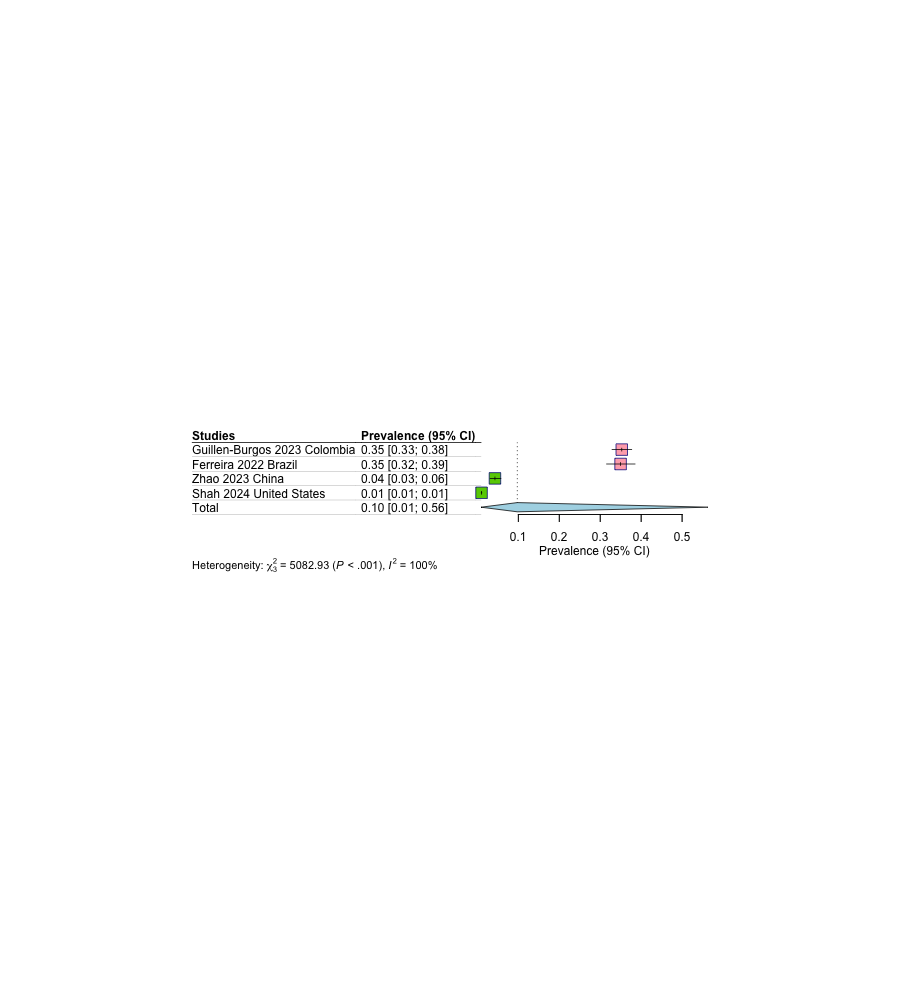

Cardiovascular

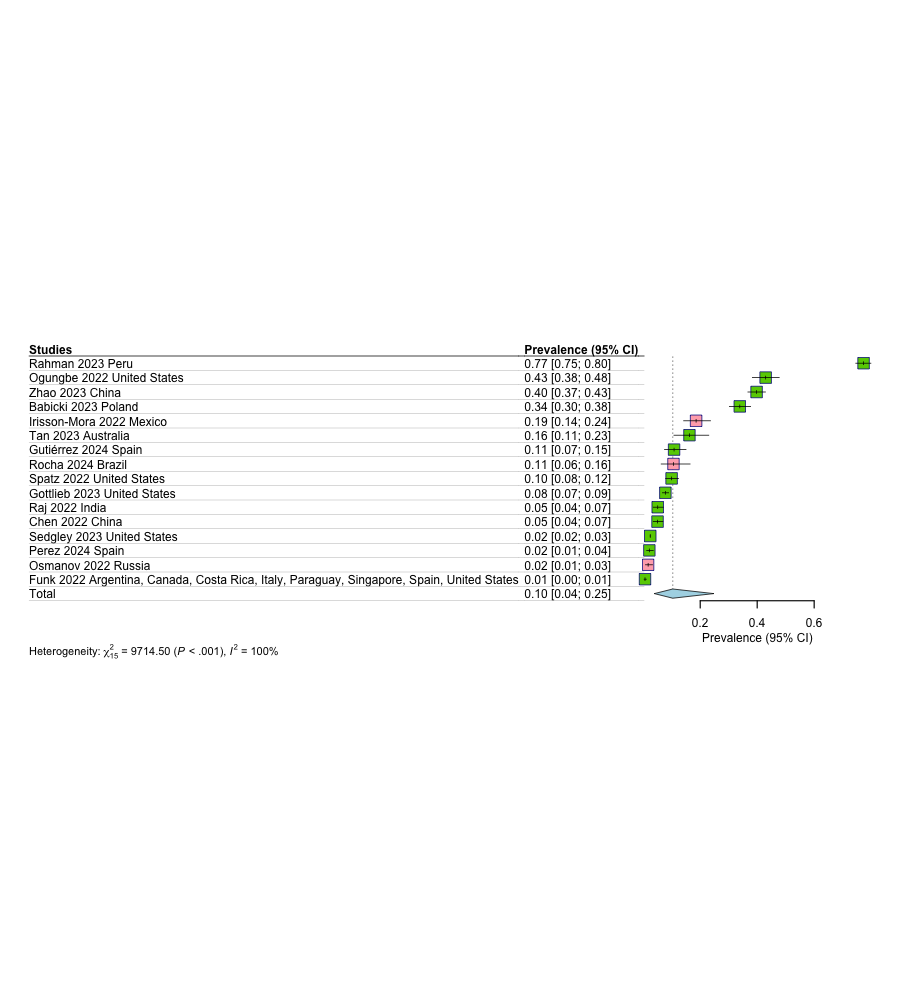

Cardiovascular Follow-up

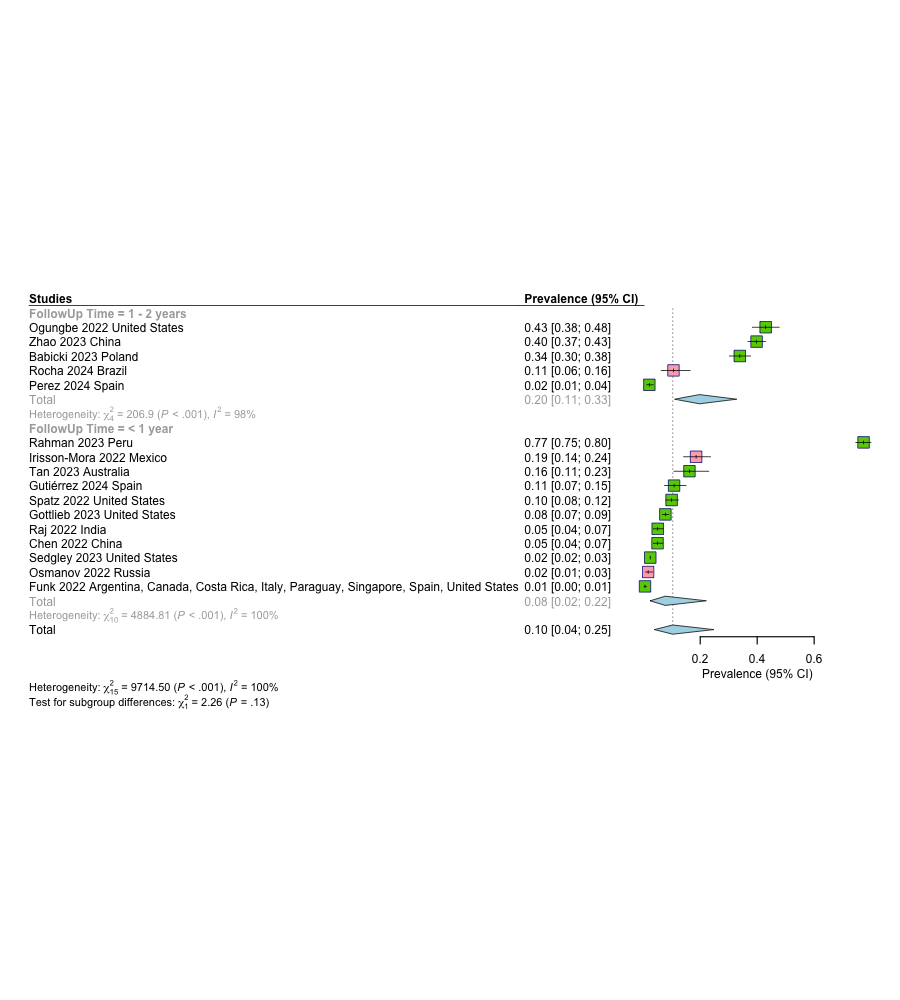

Arrhythmia

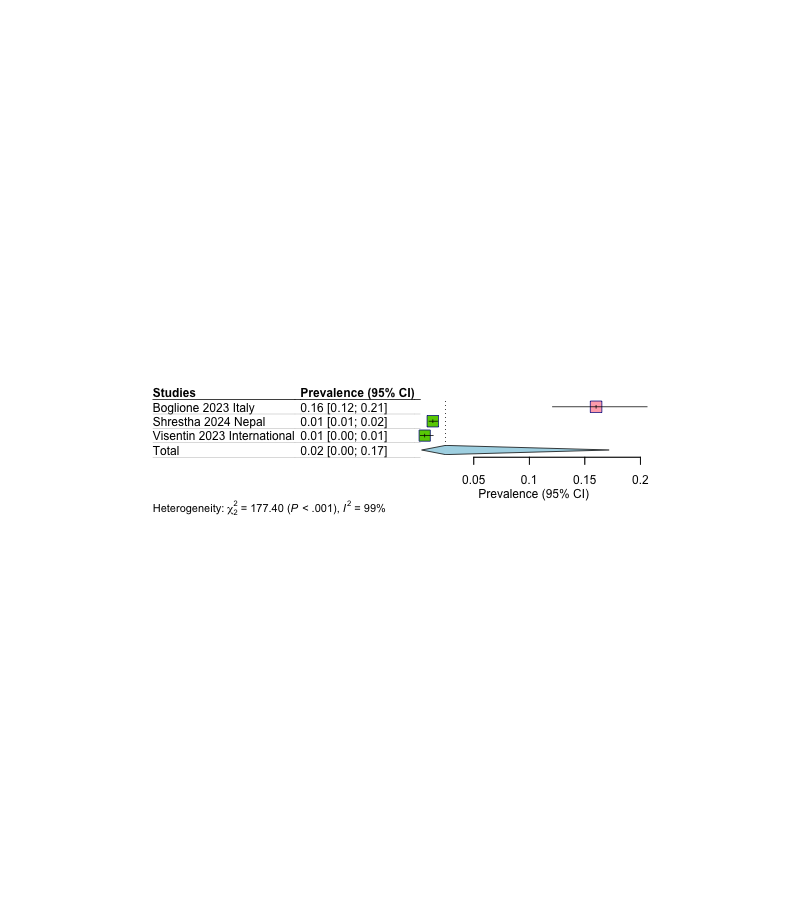

Cardiac impairment

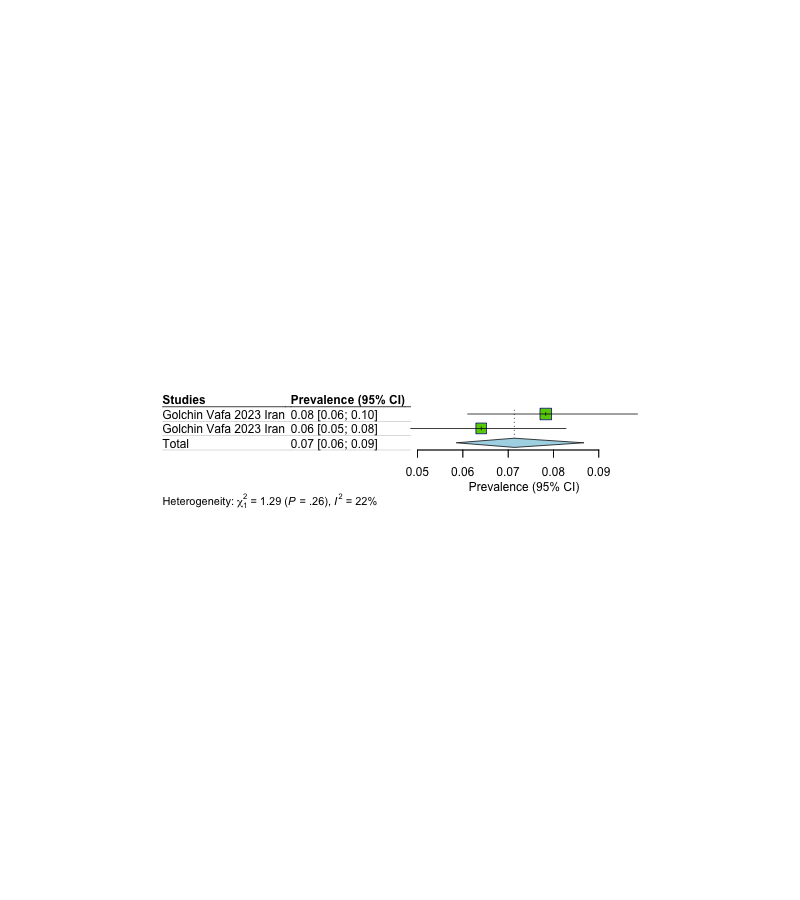

Hypertension

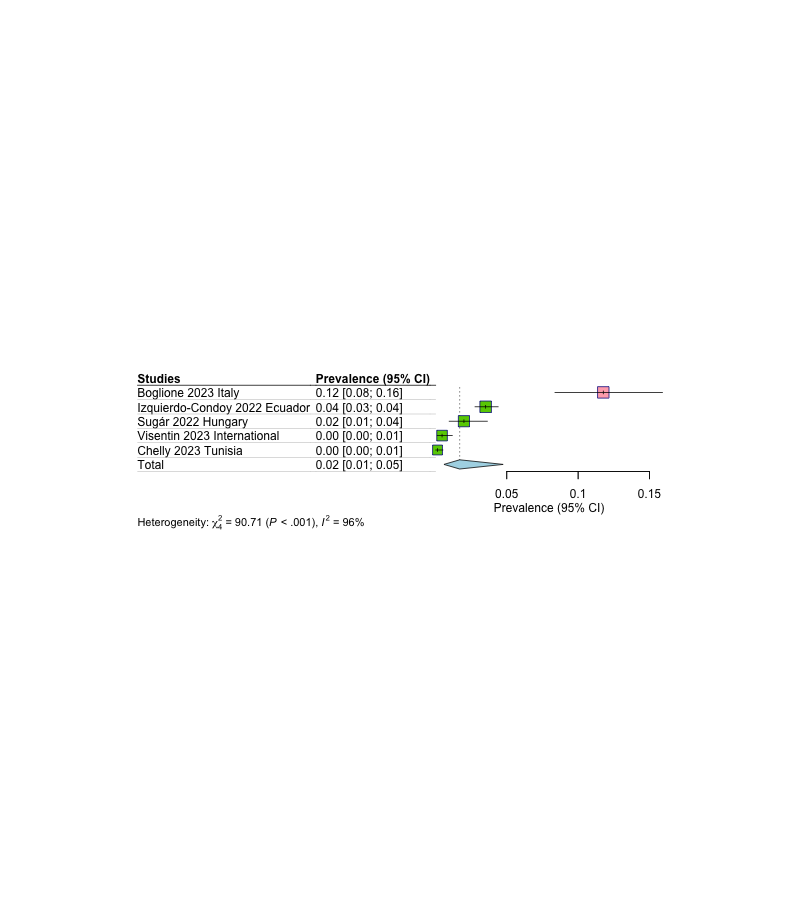

Palpitations

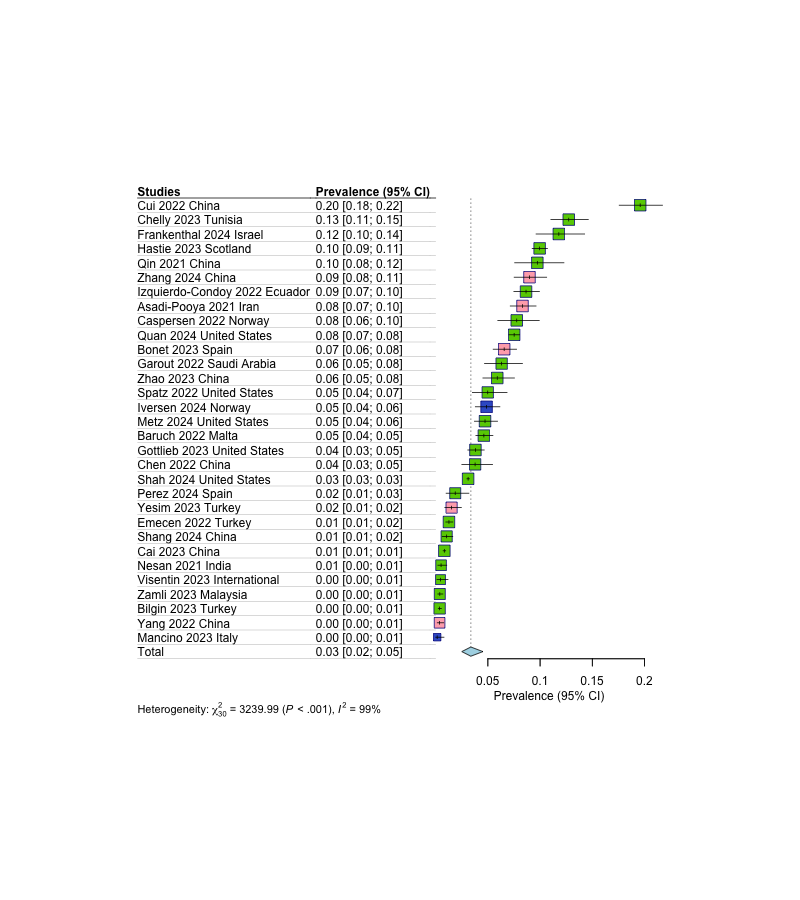

Tachycardia

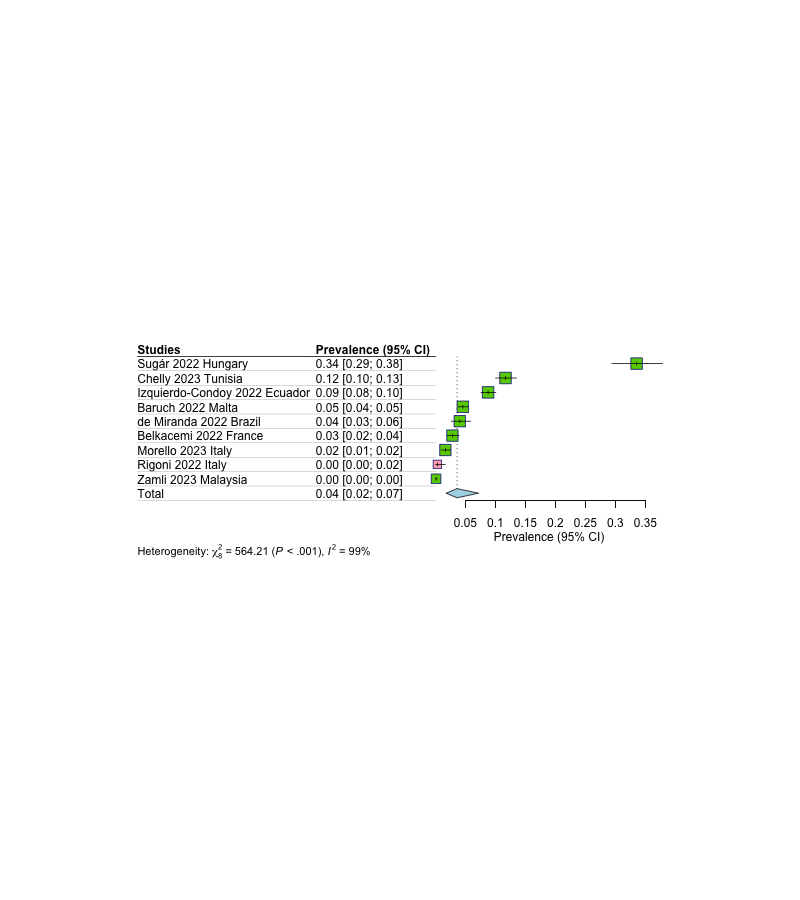

Respiratory Subtype

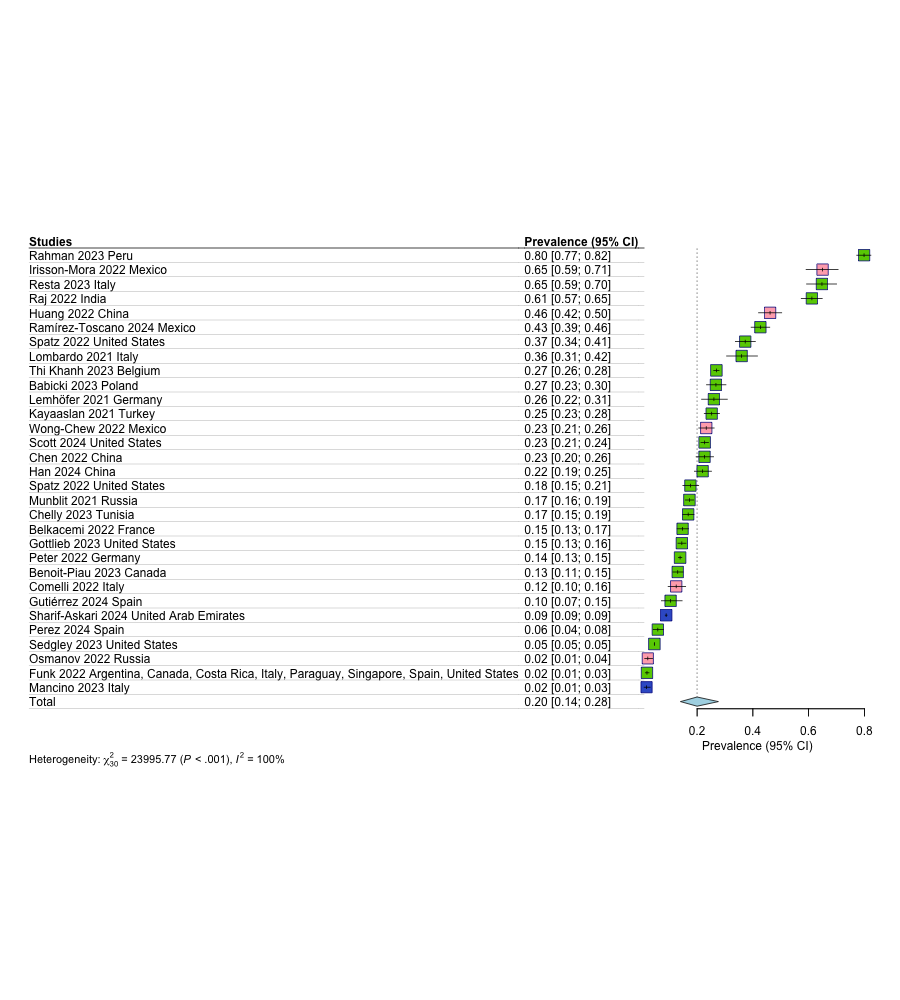

Respiratory Follow-up

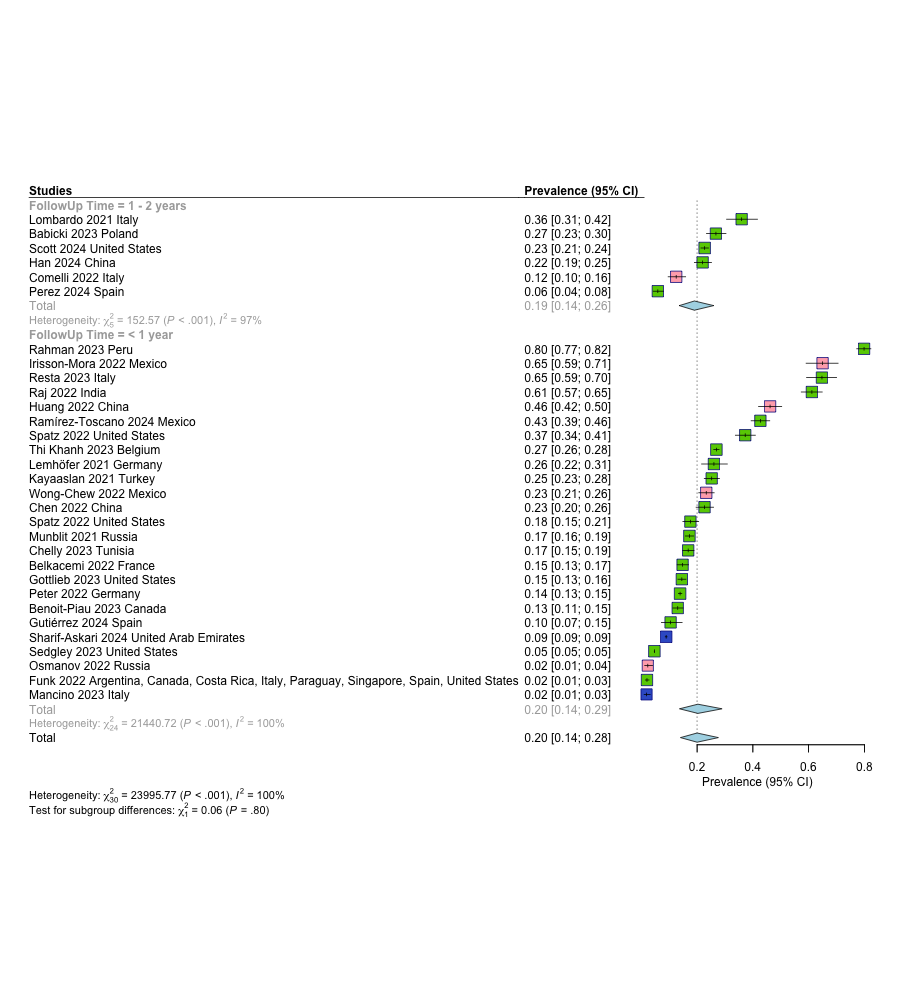

Breathlessness

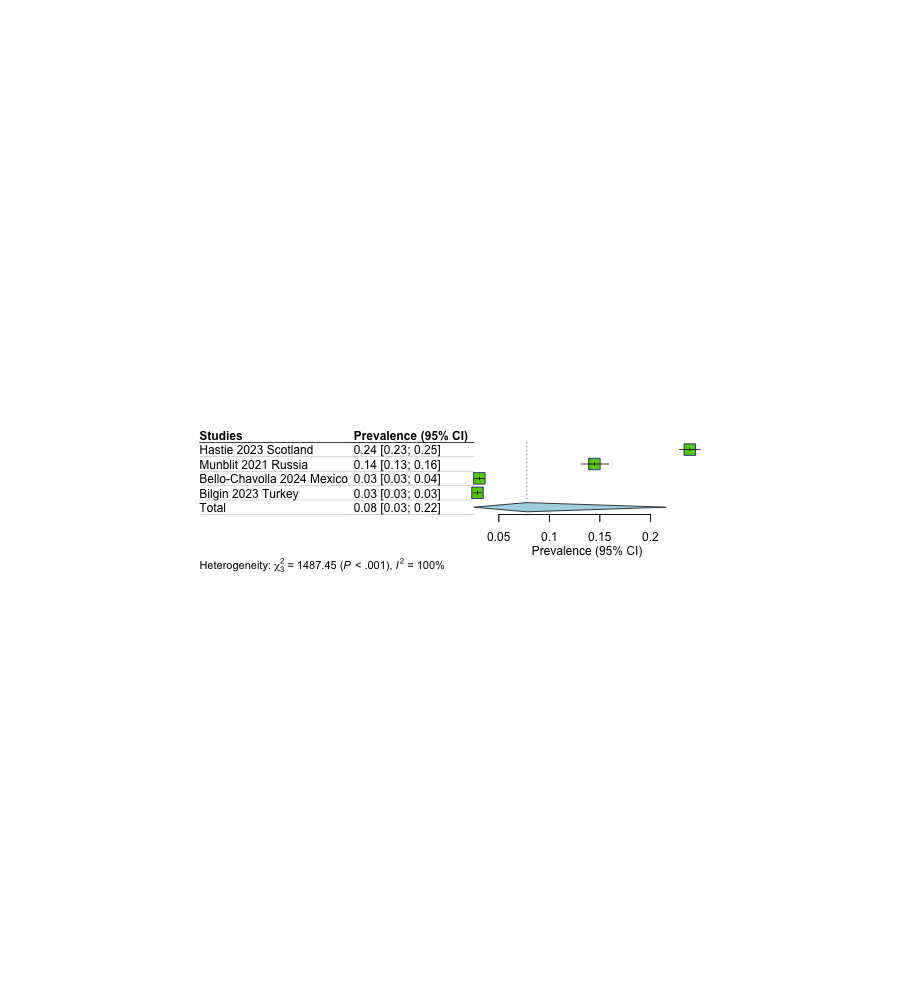

Chest pain or discomfort

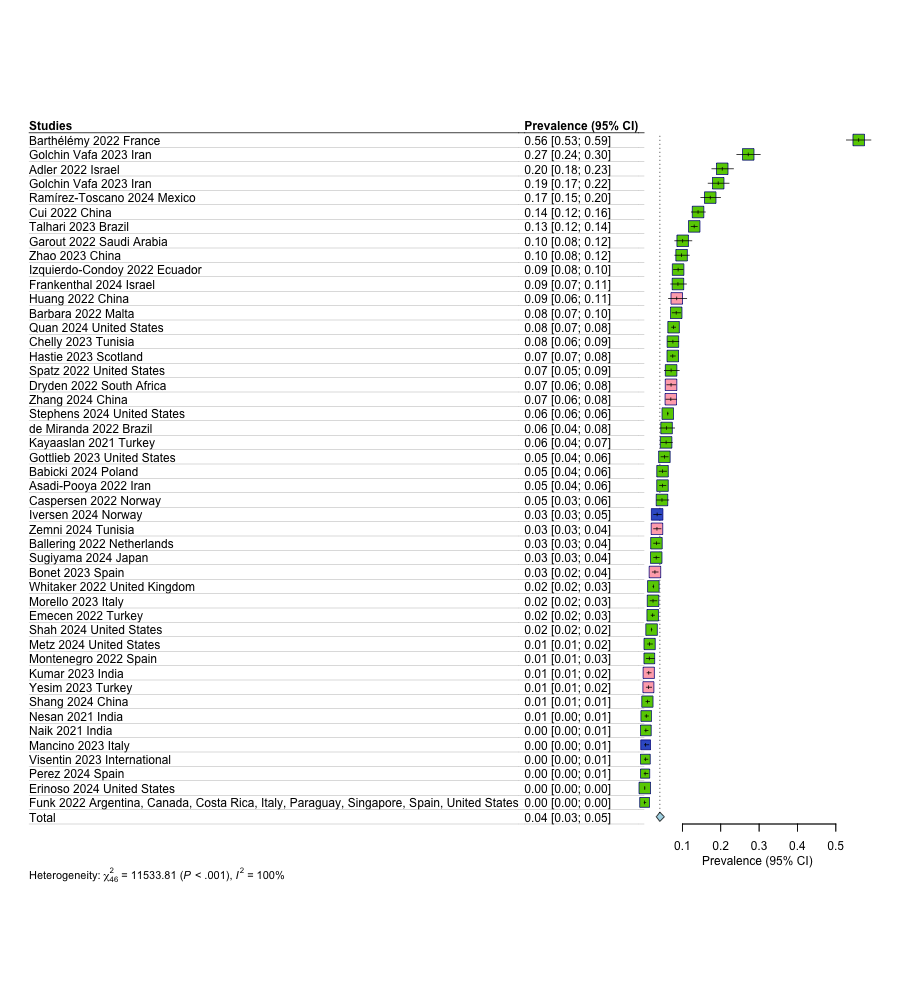

Chest tightness

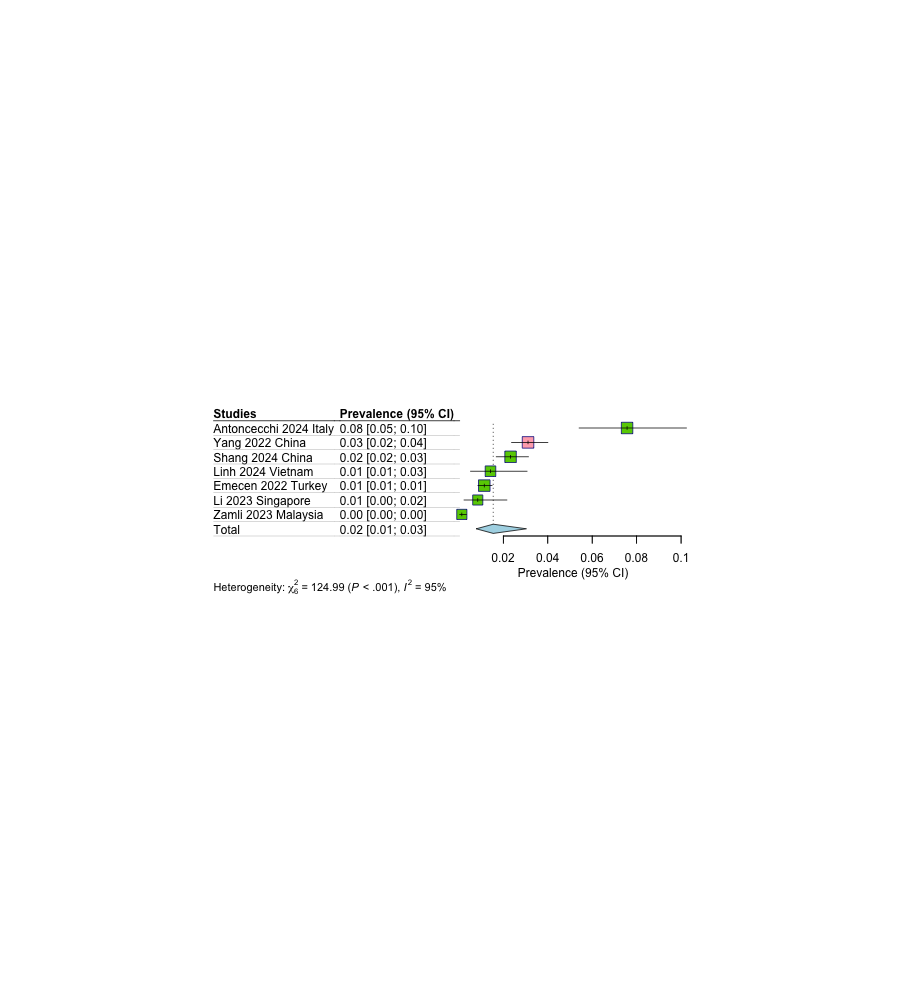

Cough/chronic cough

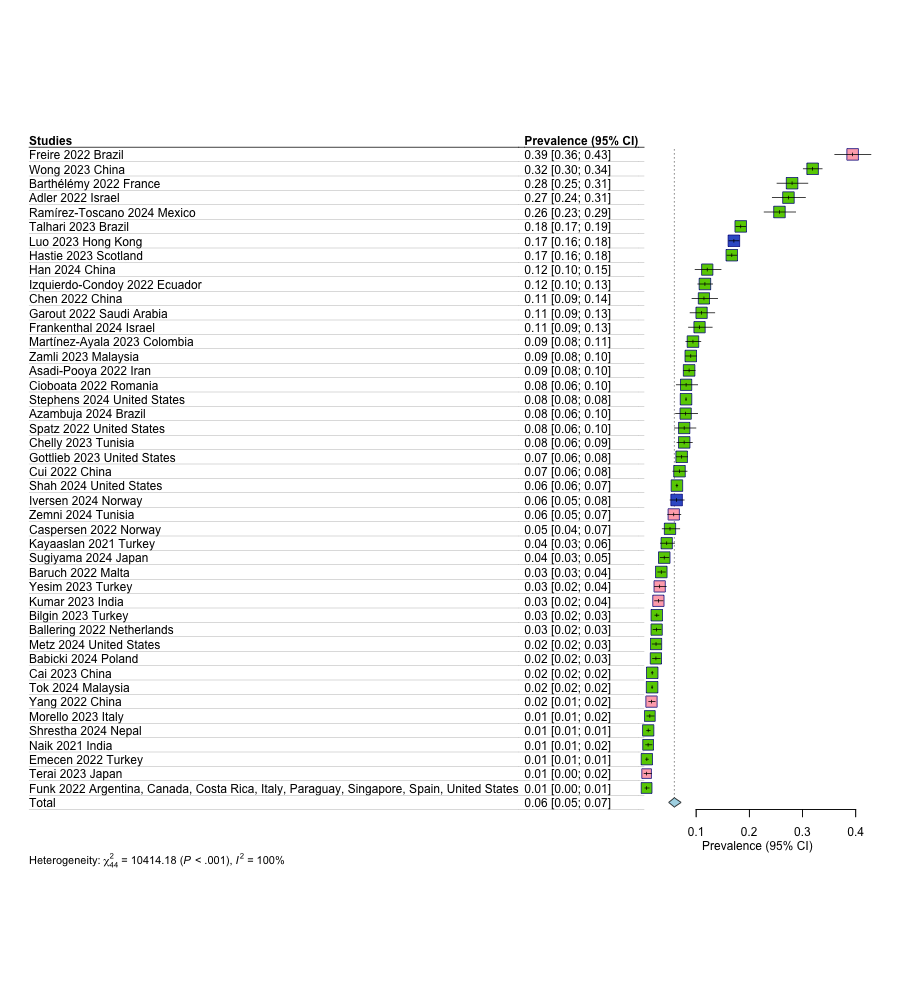

Dyspnea

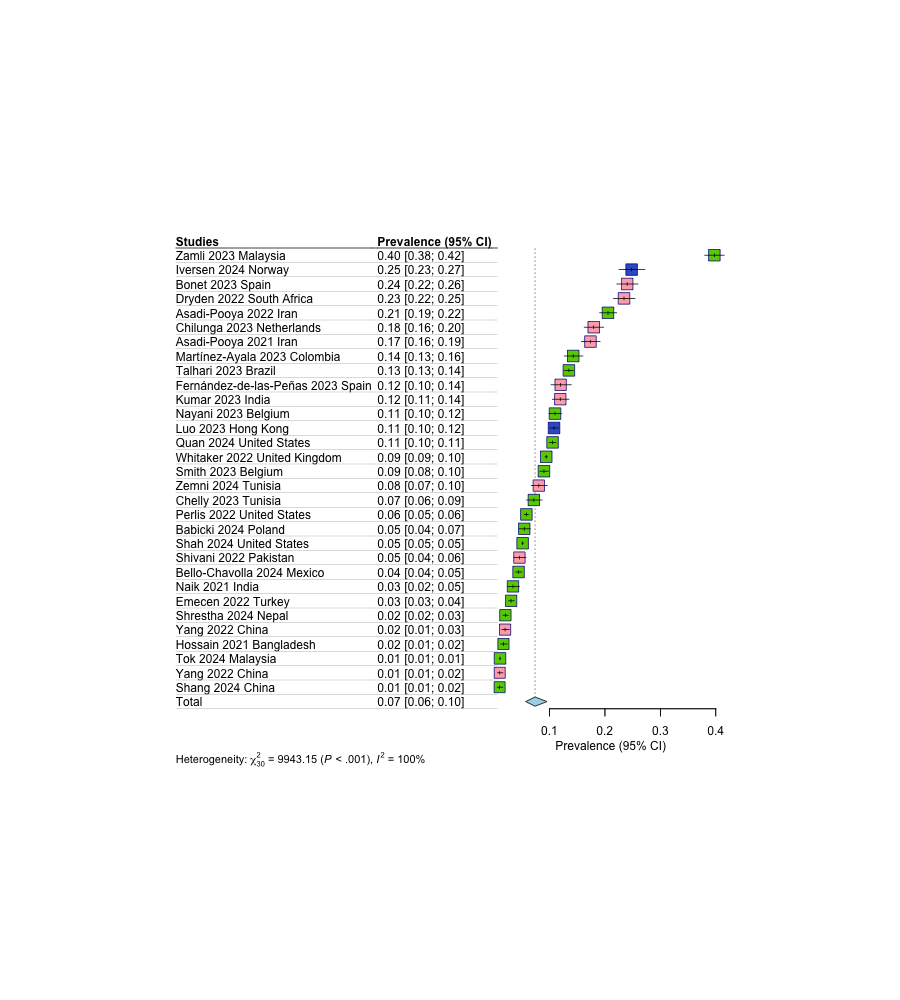

Nasal congestions

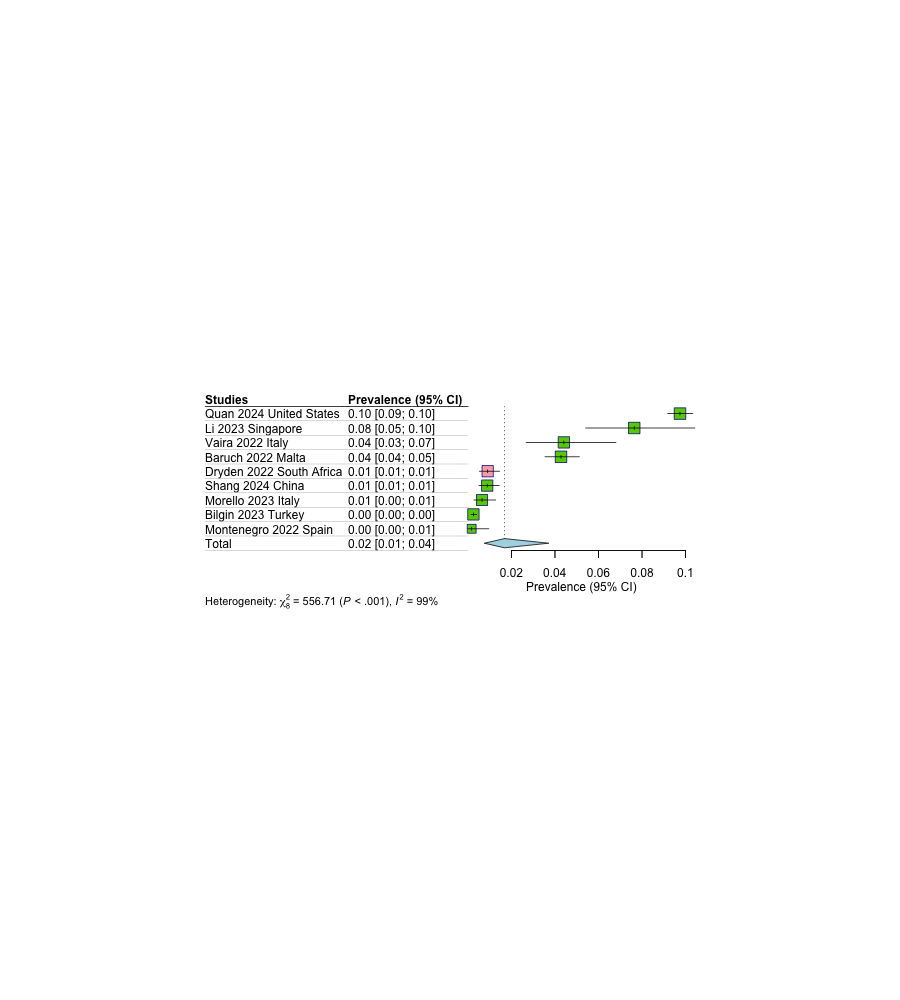

Gastrointestinal Subtype

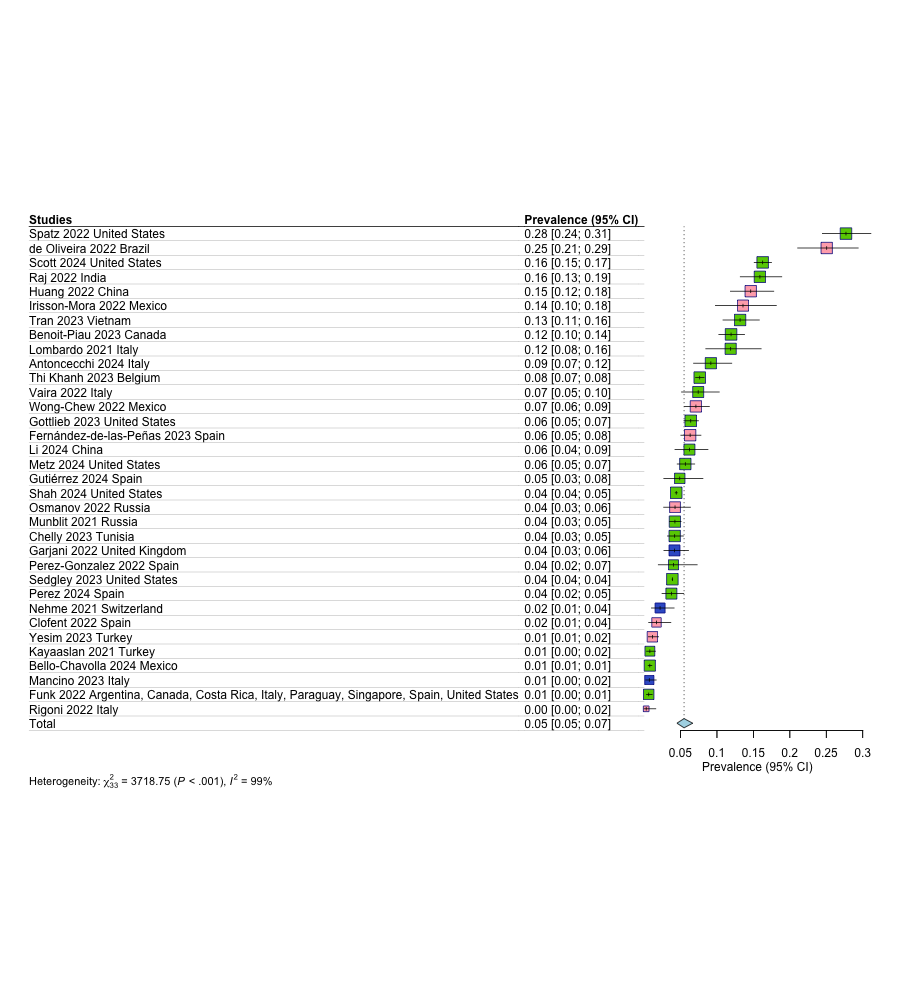

Gastrointestinal Follow-up

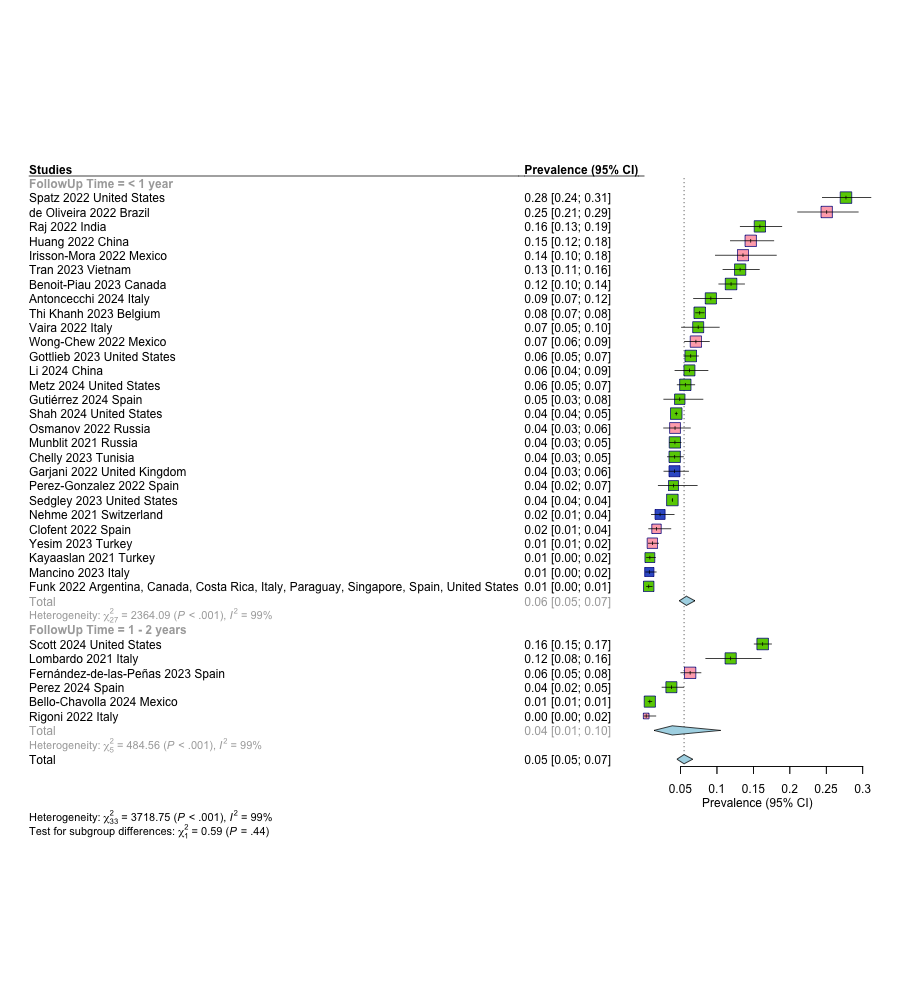

Abdominal pain

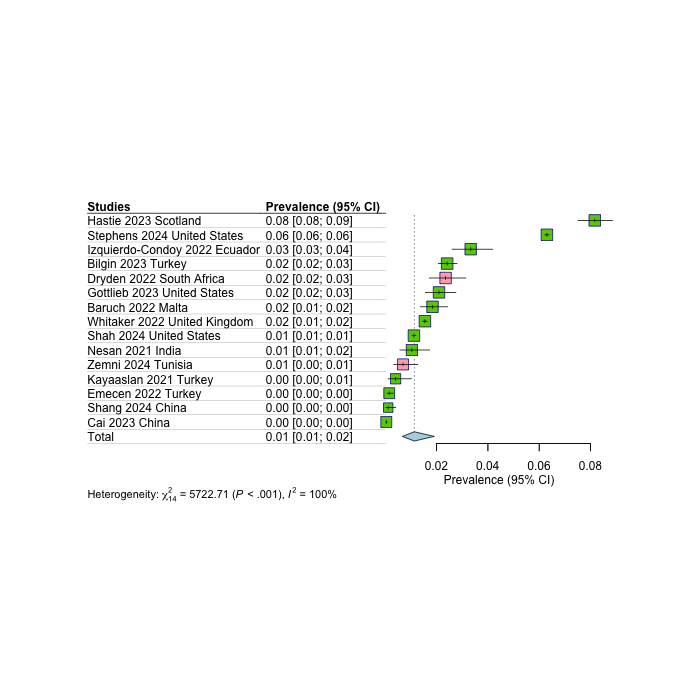

Constipation

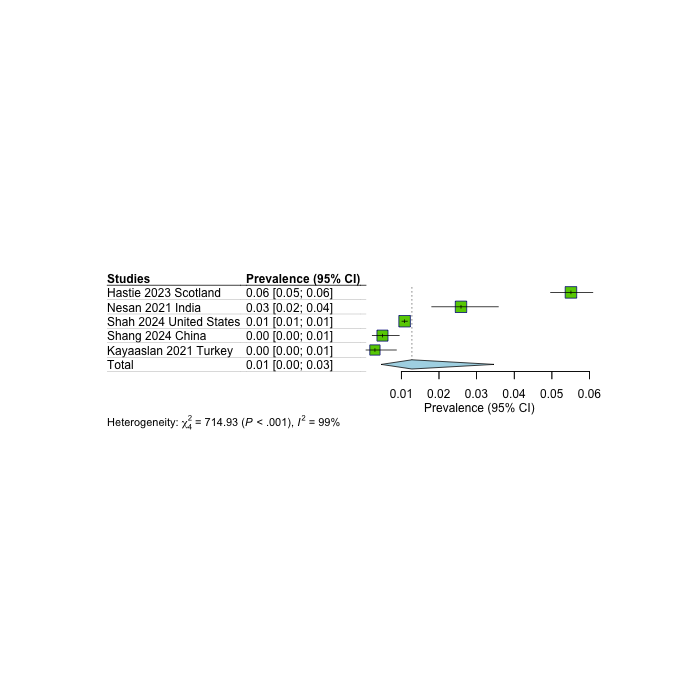

Diarrhea

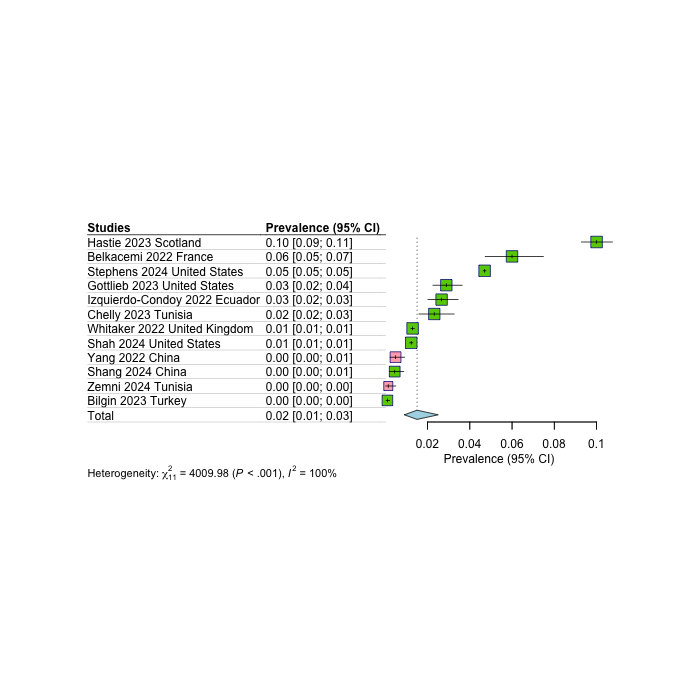

Stomach pain

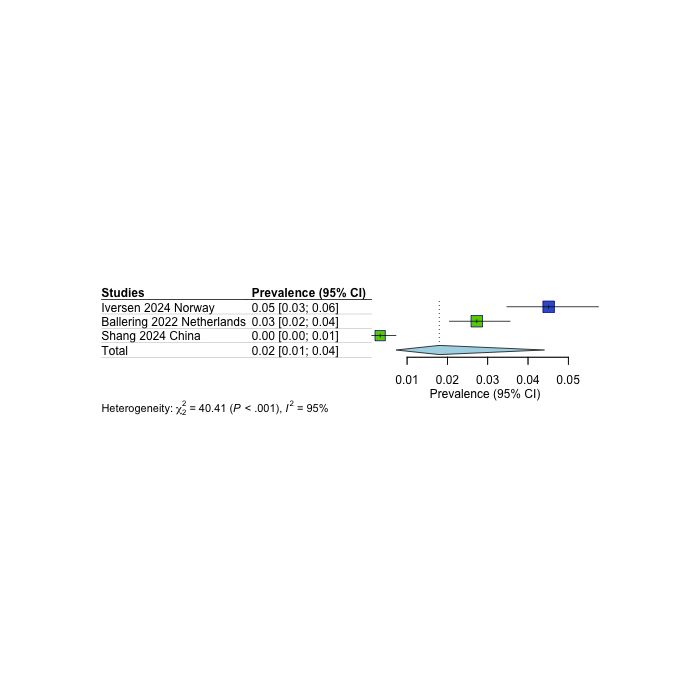

Musculoskeletal Subtype

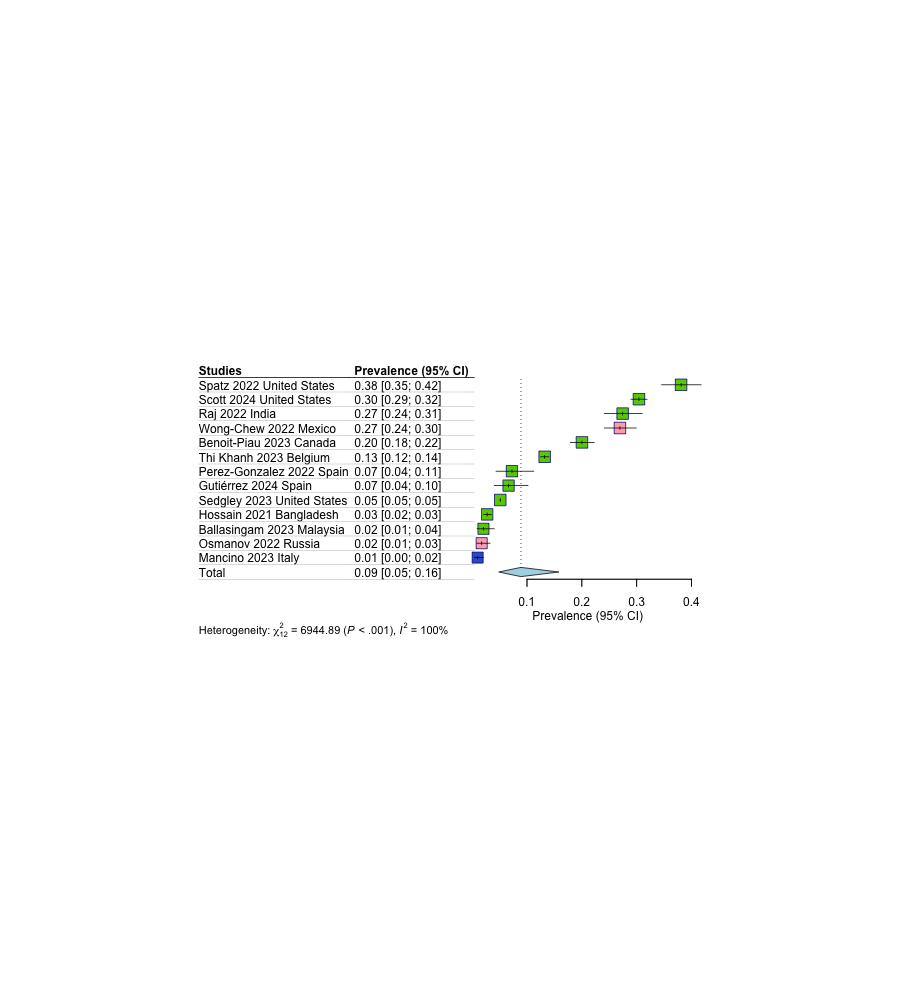

Musculoskeletal Follow-up

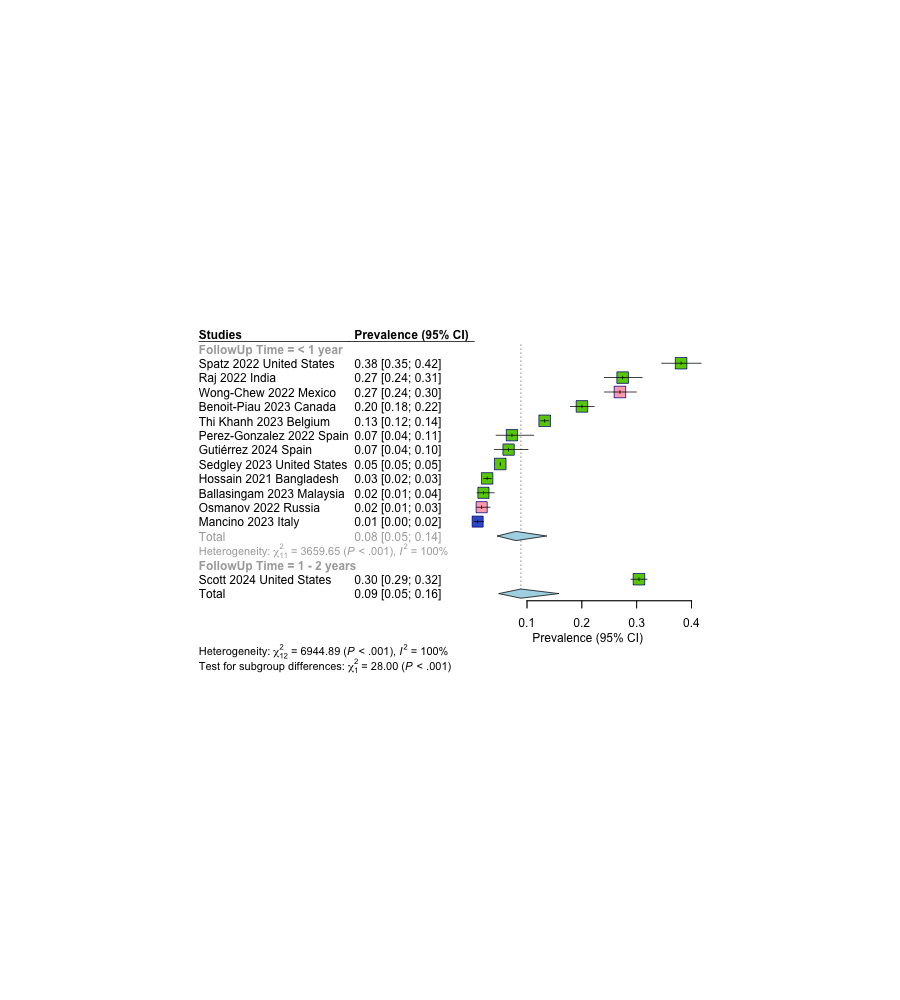

Joint pain

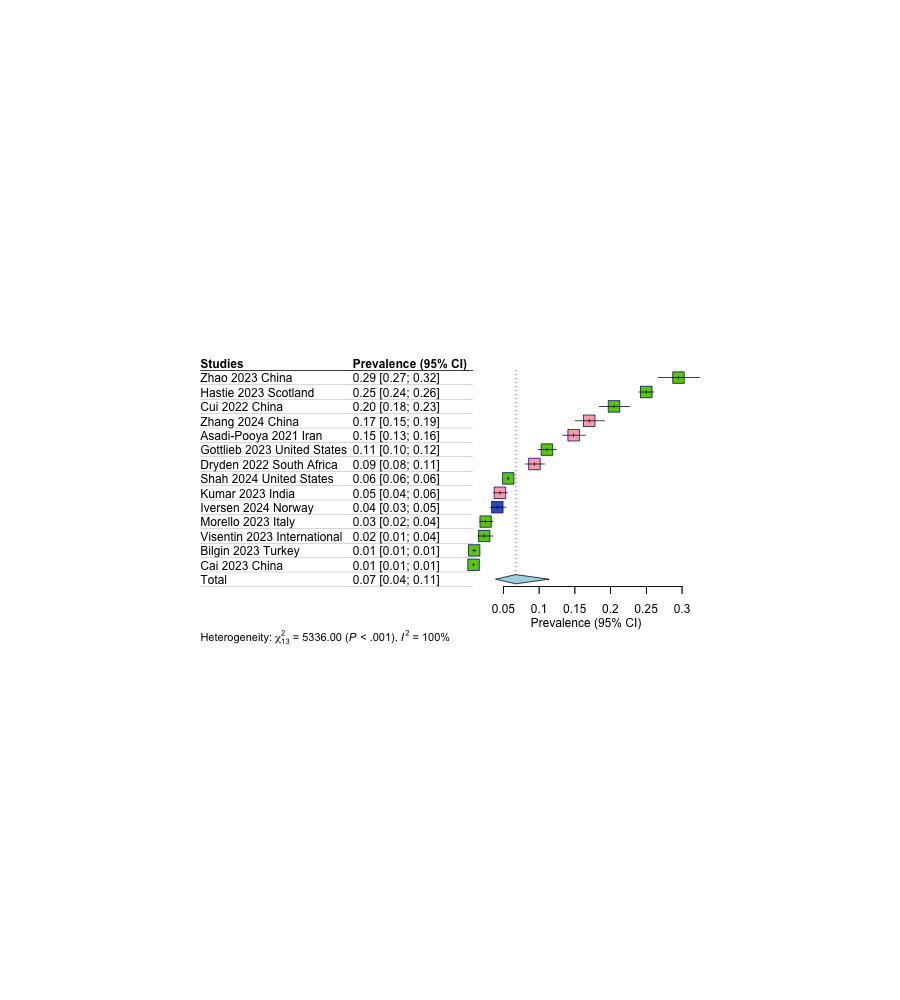

Muscle weakness

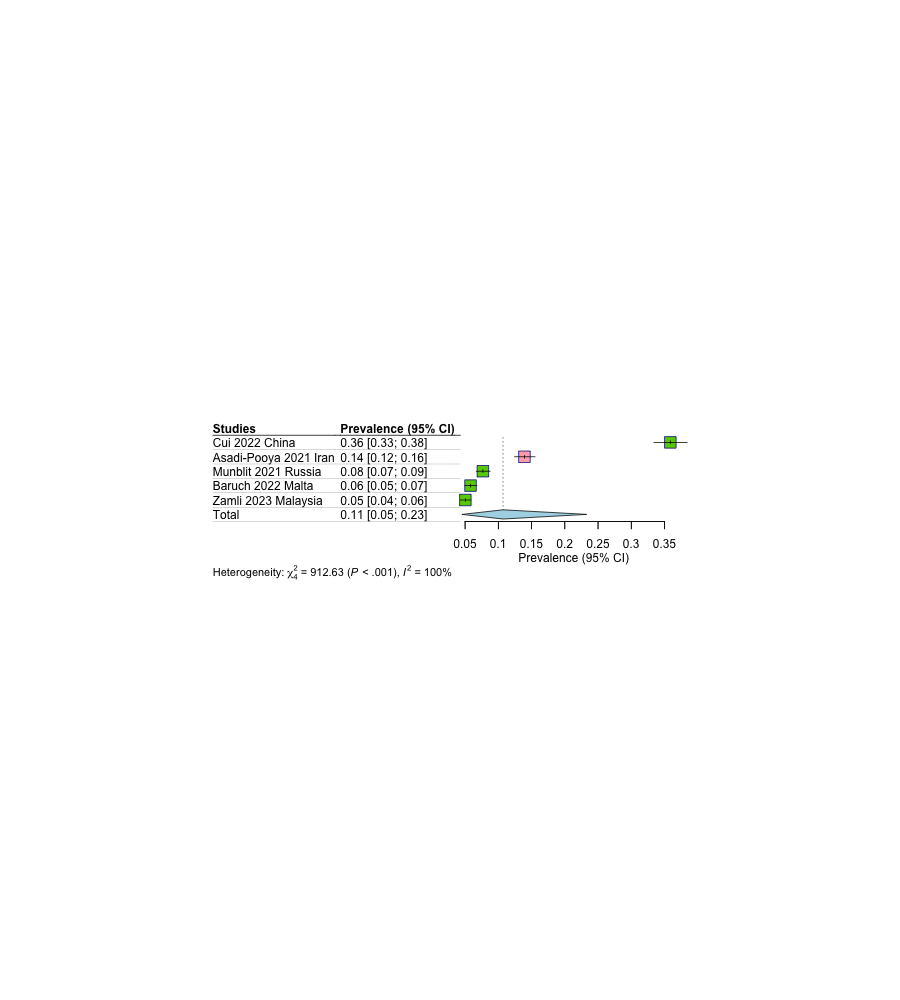

Myalgia

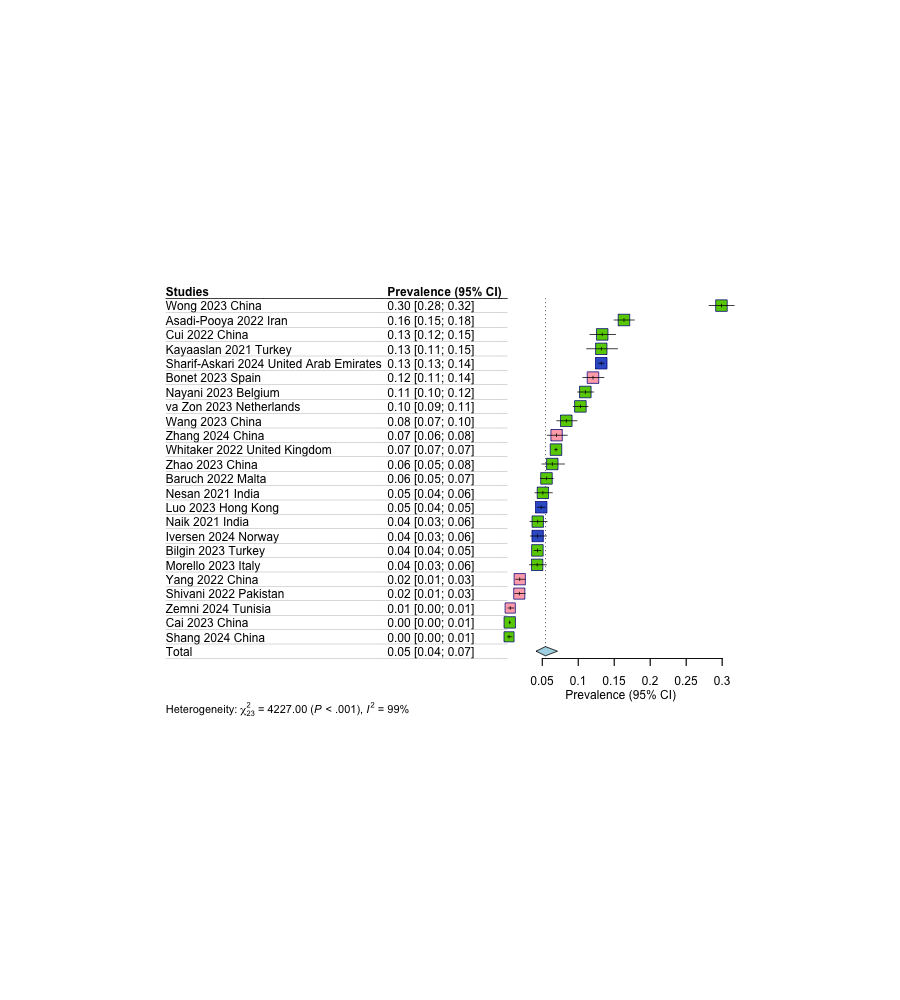

Swelling

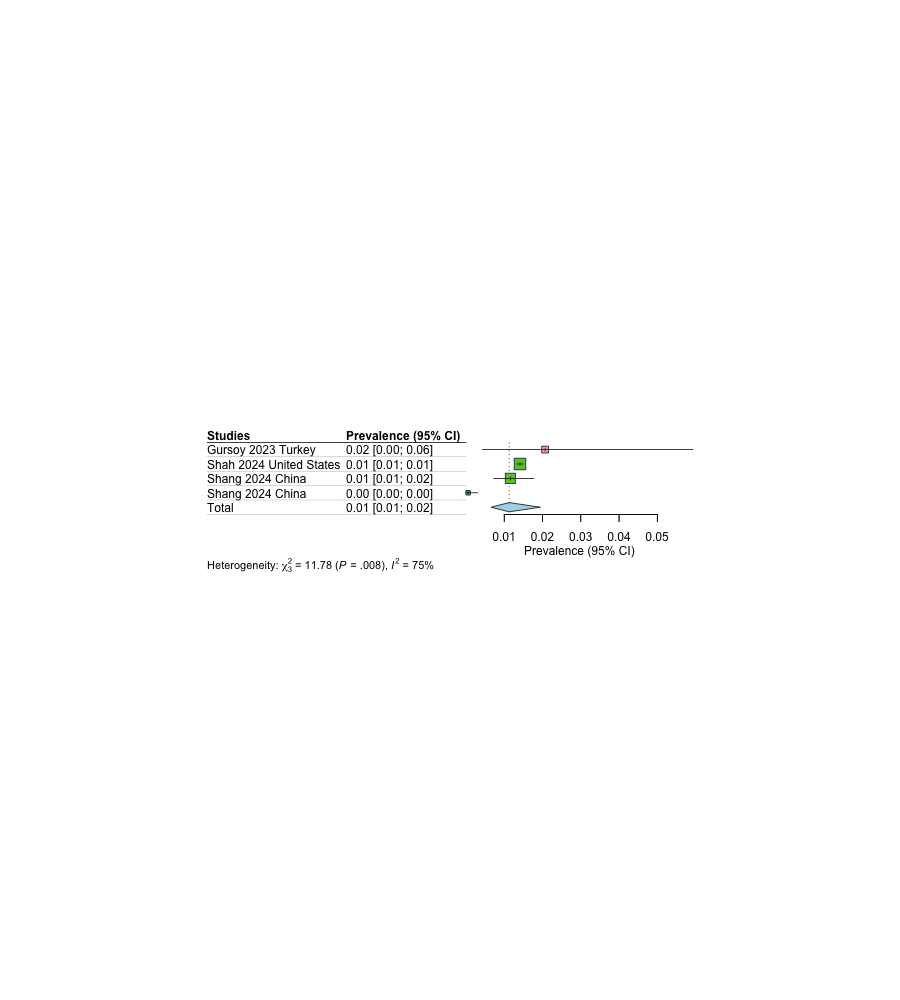

Dermatological Subtype

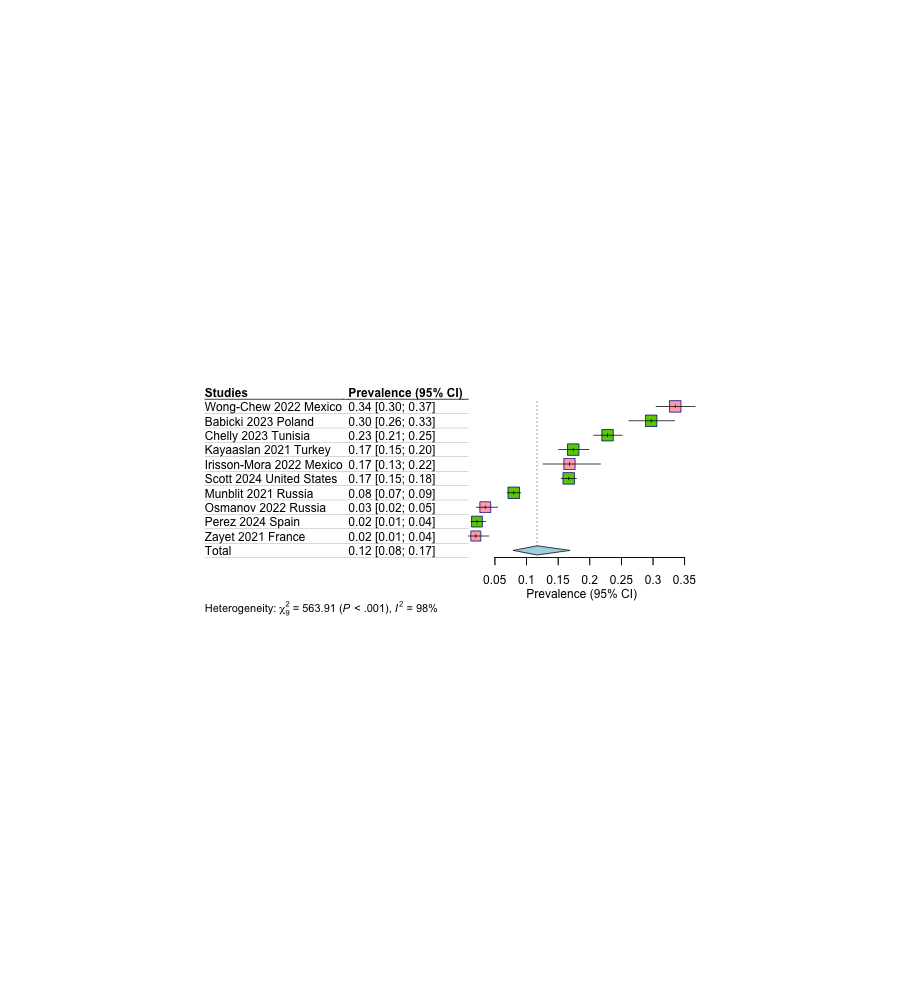

Dermatological Follow-up

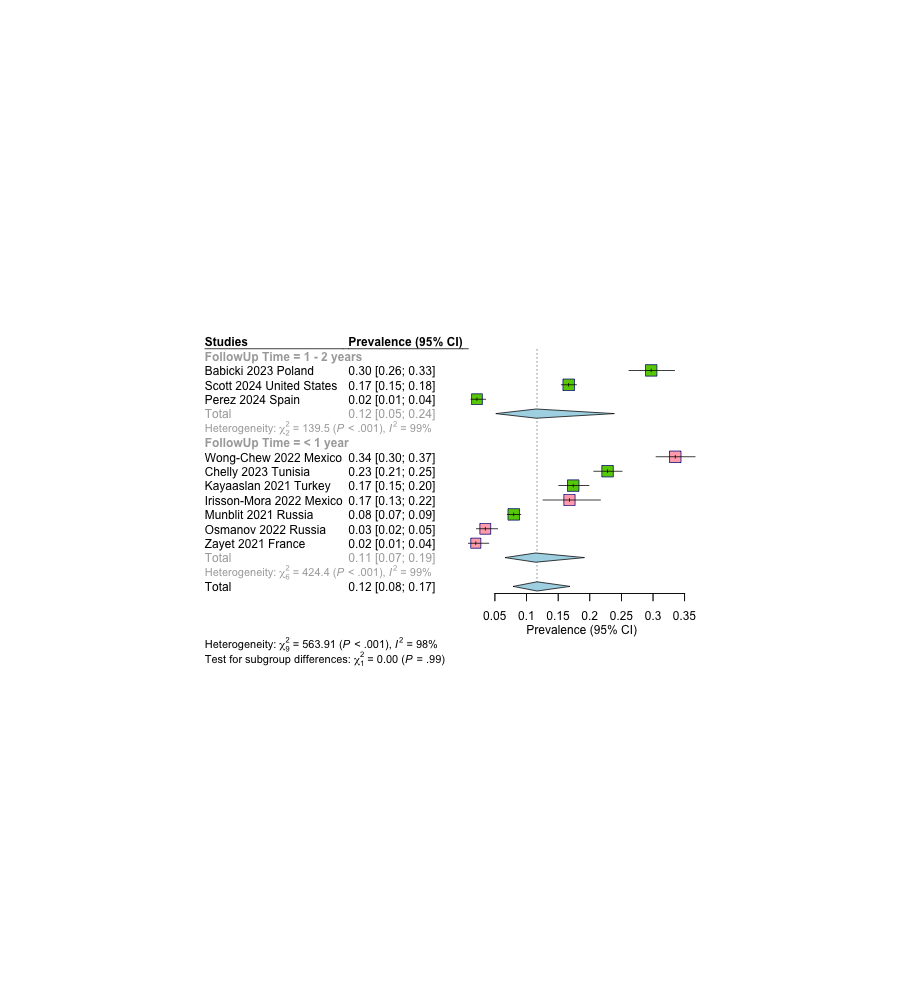

Hair loss

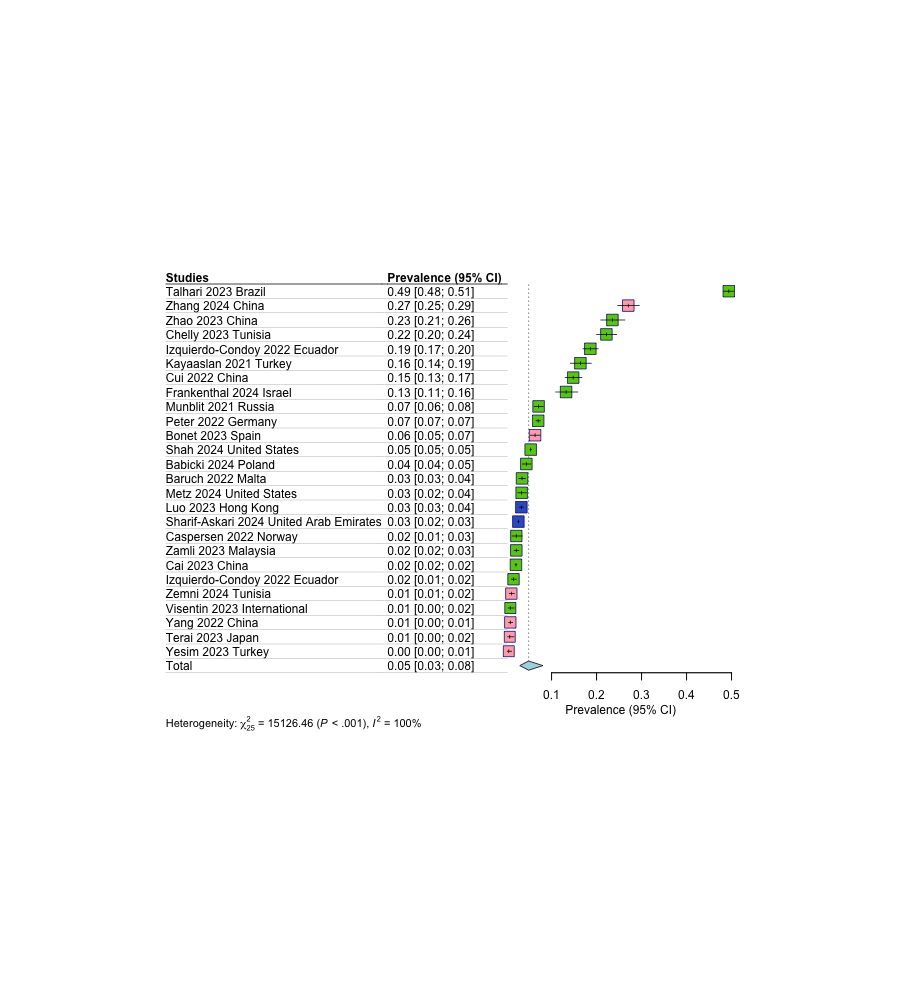

Skin rash

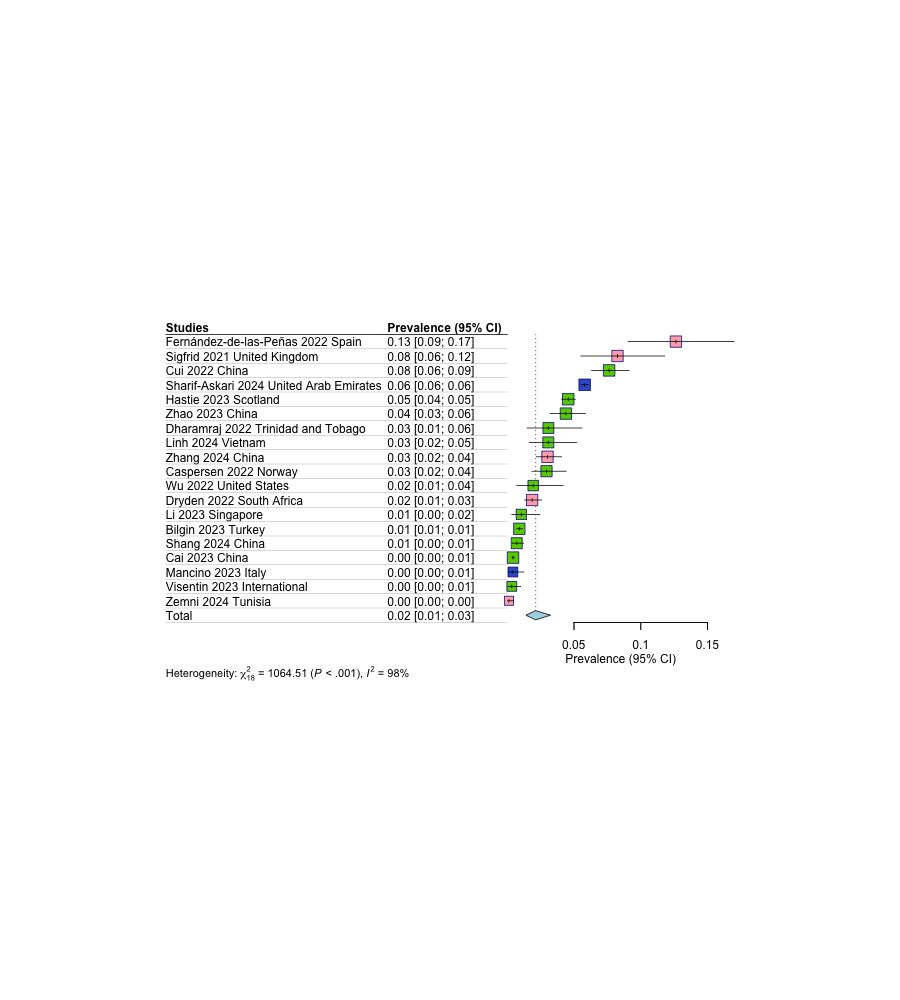

General Fatigue

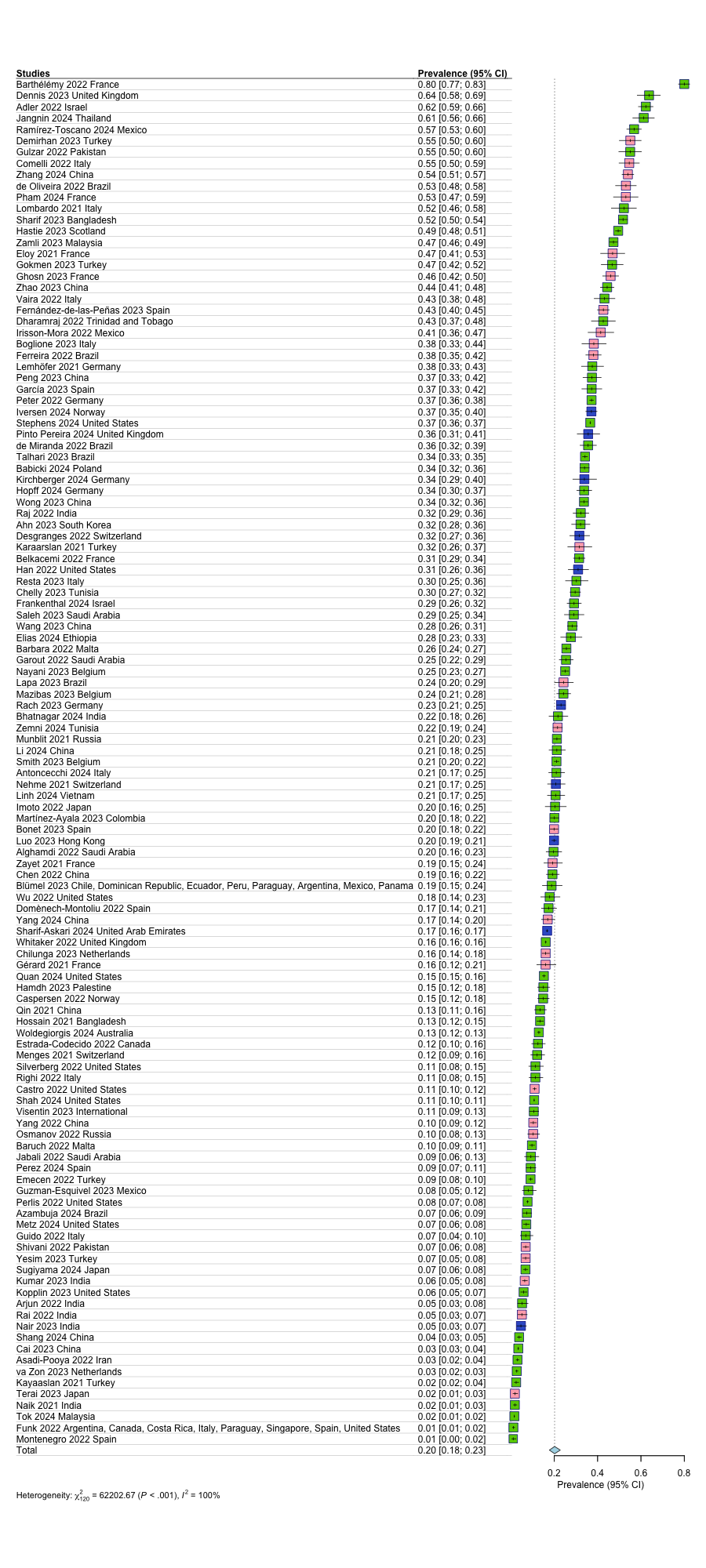

General Fatigue Follow-up

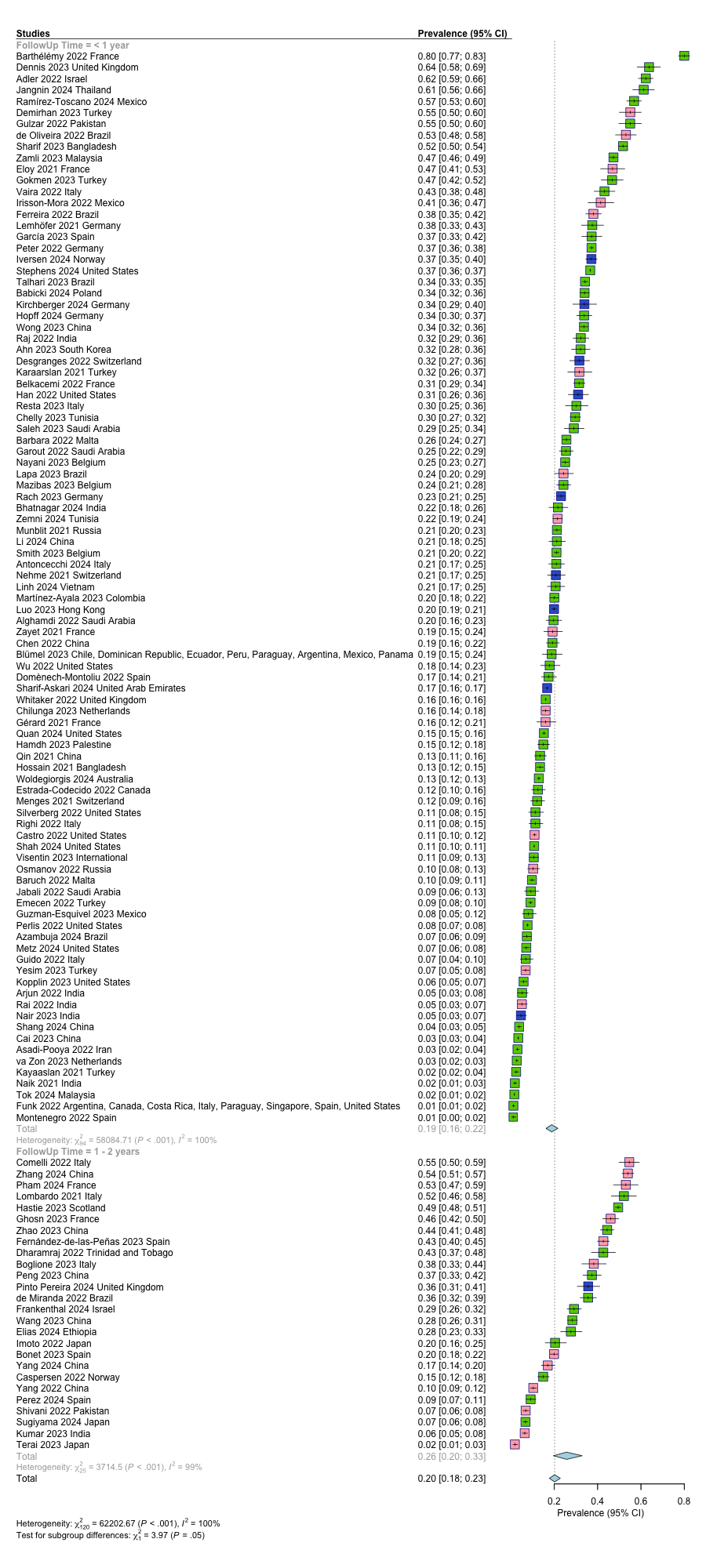

Appetite

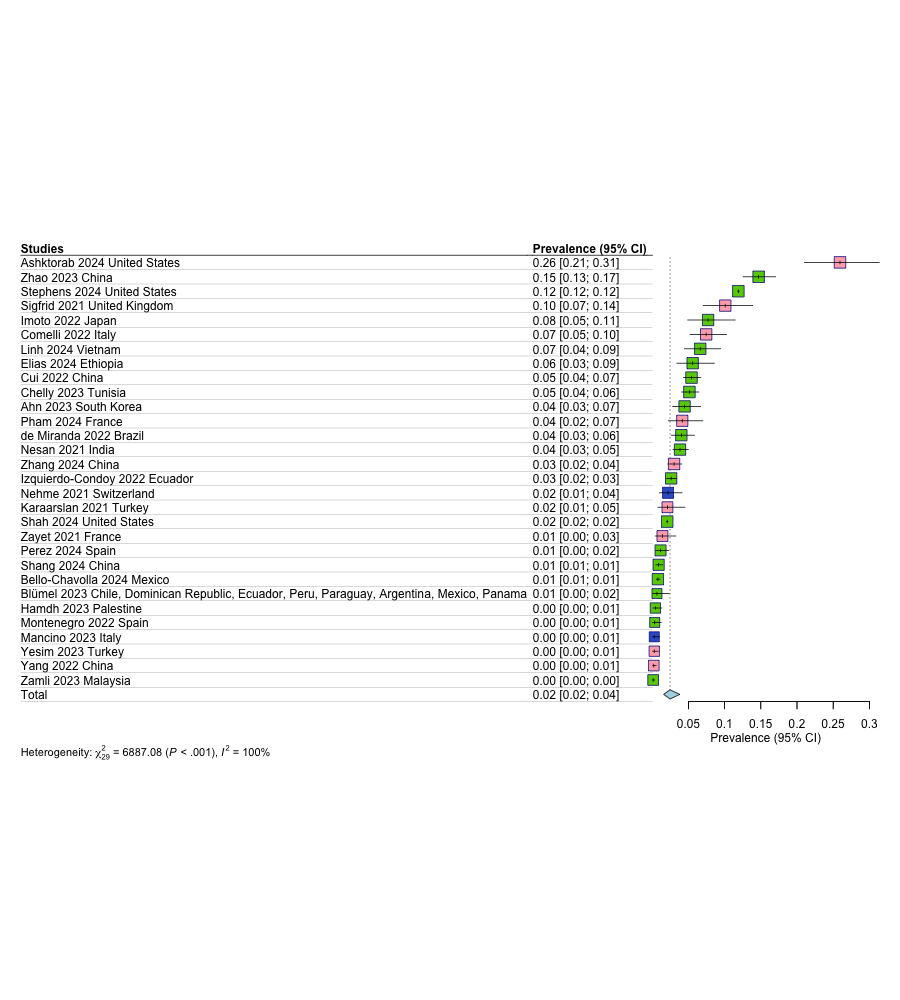

Dizziness

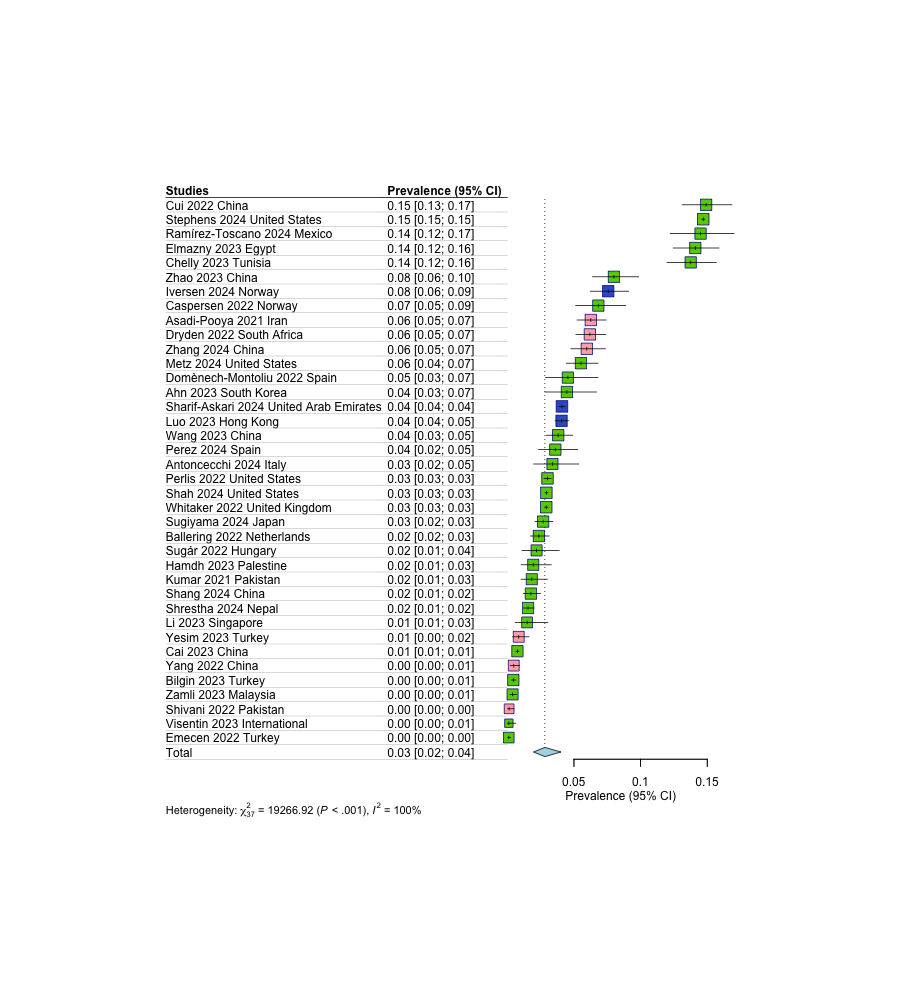

Fever

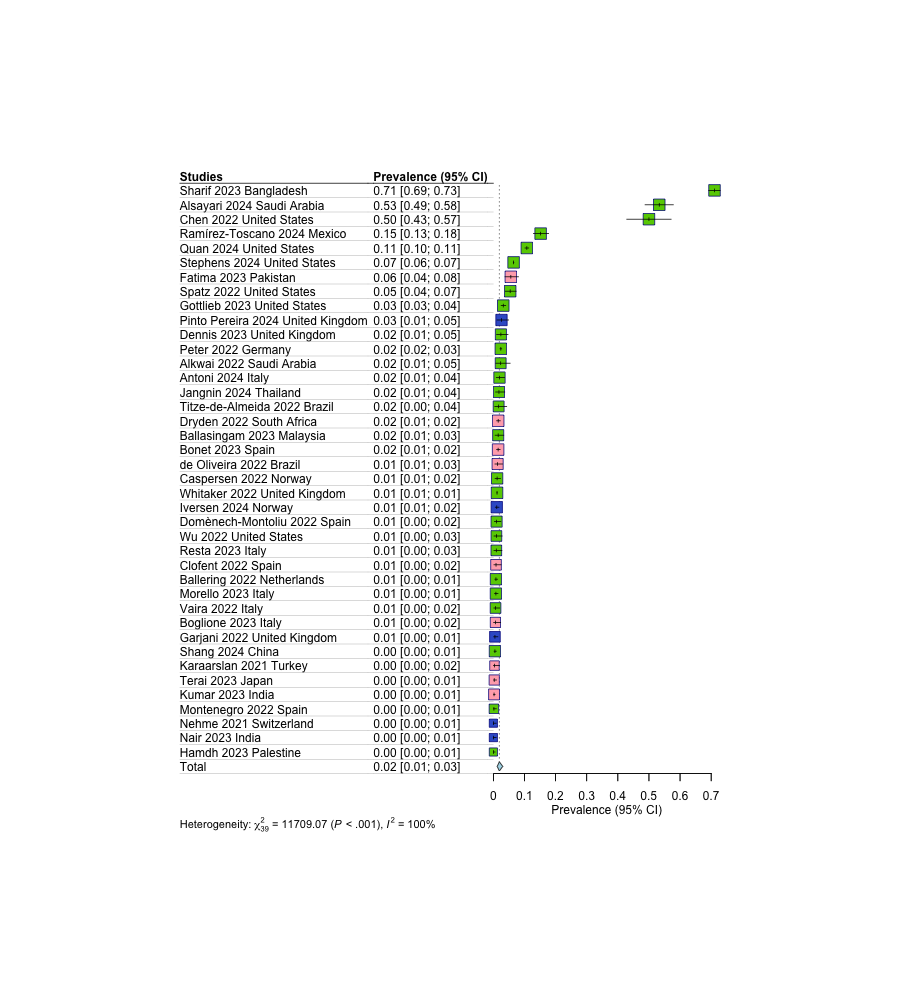

Sore Throat

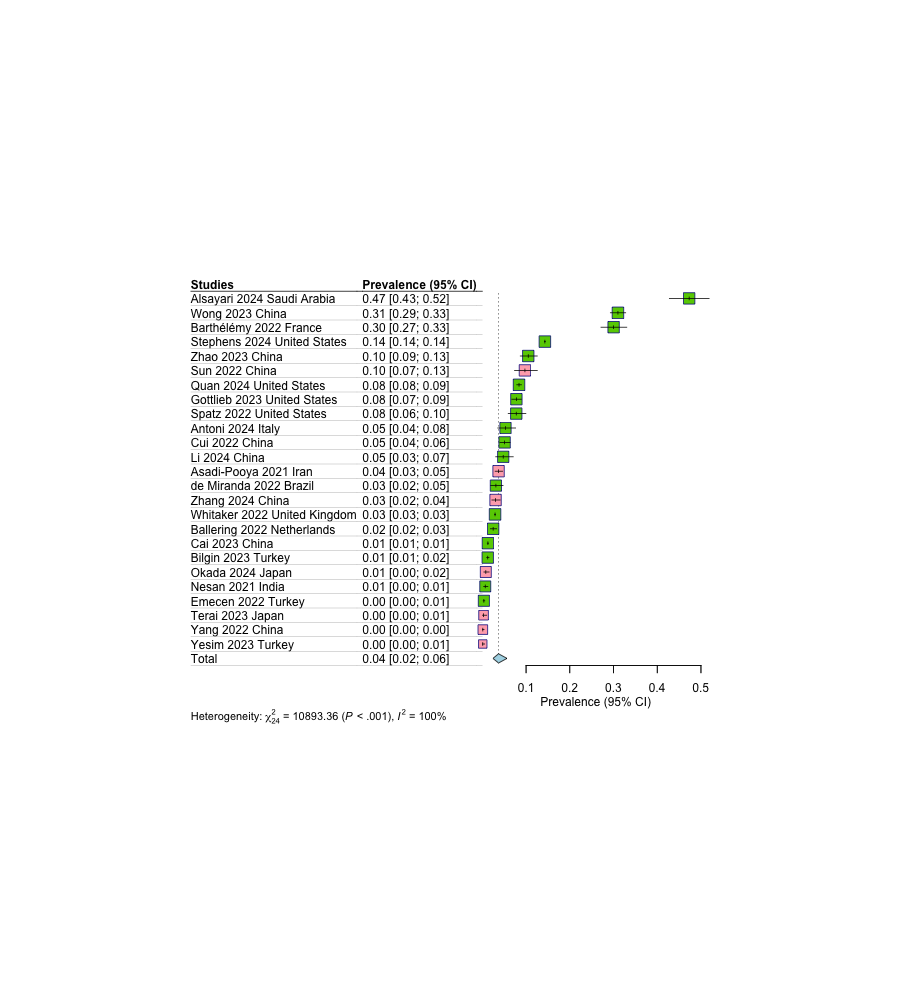

Sweats

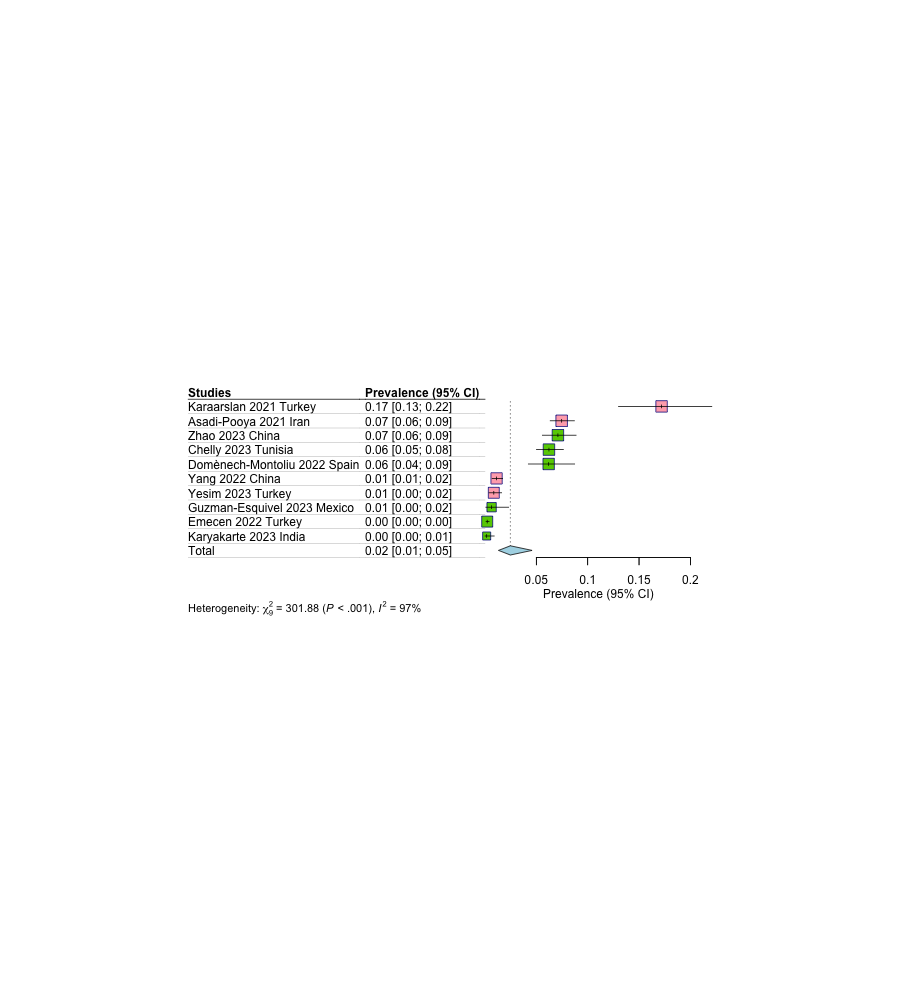

**eFigure 9.** Forest plots of risk factors for long COVID in COVID-19 positive individuals

| a) Age per year   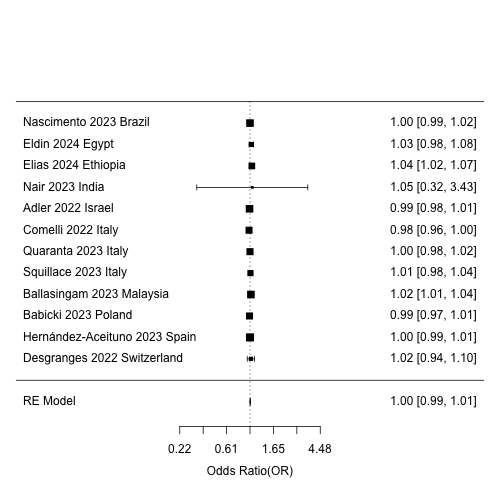 | b) At least one comorbidity   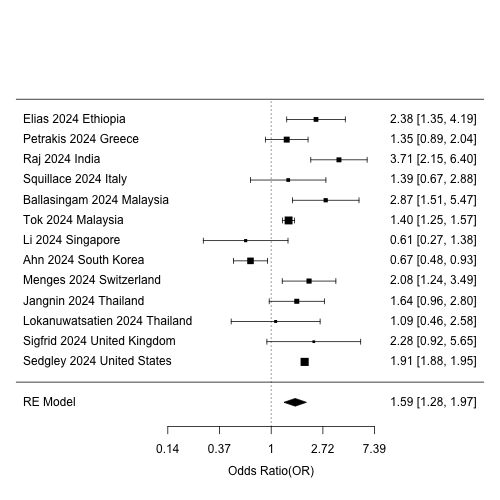 |
| --- | --- |

| c) Cardiovascular disease   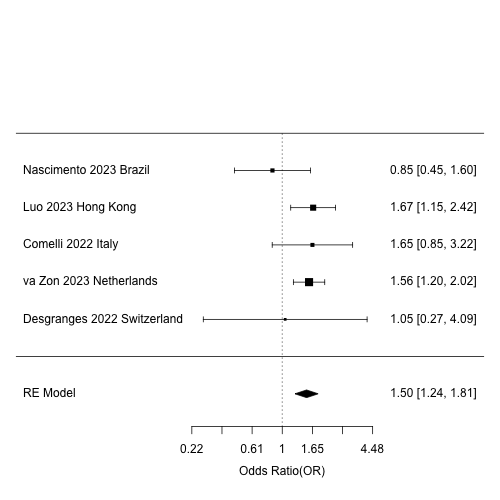 | d) Diabetes   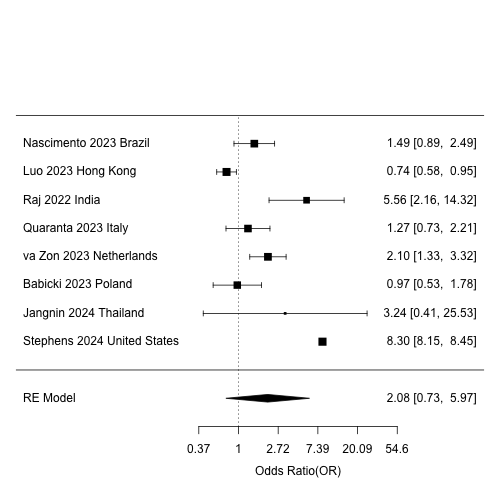 |
| --- | --- |

| e) Female sex   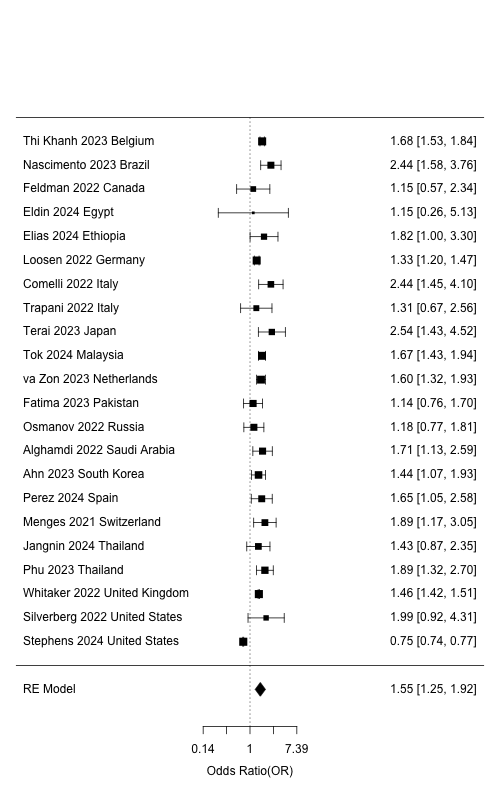 | f) History of Chronic Obstructive Pulmonary Disease (COPD)   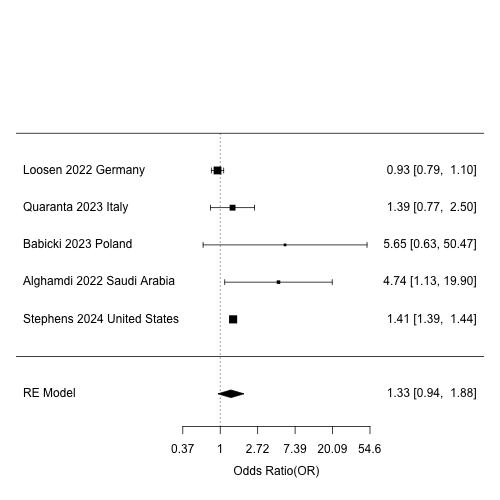 |
| --- | --- |
| g) Hypertension   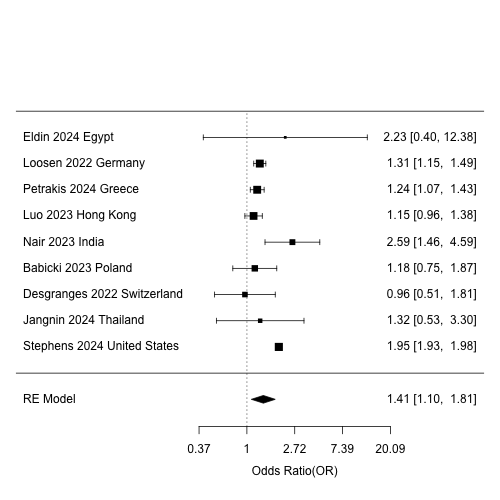 | h) Intensive care unit (ICU admission)   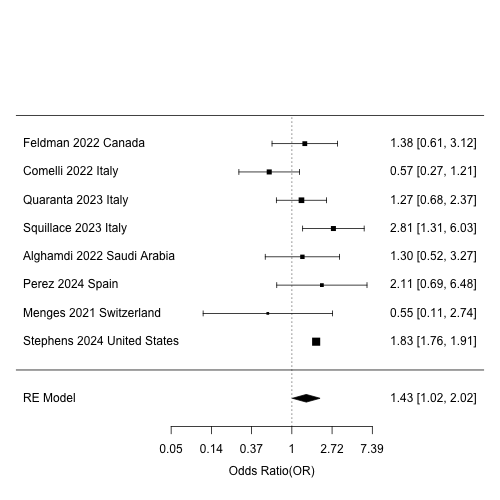 |

| i) Obesity   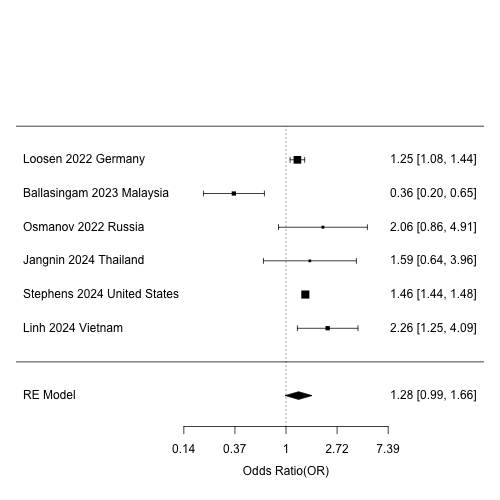 | j) Unvaccinated for COVID-19   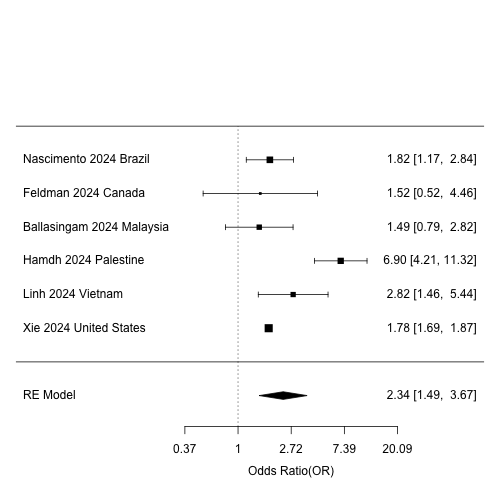 |
| --- | --- |
