## Supplementary material for "Global Prevalence of Long COVID, its Subtypes and Risk factors: An Updated Systematic Review and Meta-Analysis": eMethods 1a. Unregistered systematic review protocol

**Review question**

What is the global prevalence of long COVID and its subtypes and symptoms?

What are the risk factors for long COVID?

**Searches**

Searches for publications will be conducted in PubMed, Embase, and Web of Science Core Collection.

All searches will be conducted using symptoms keywords from long COVID subtypes by means of text word terms and additional MeSH terms.

**Search strategy**

Publication period: July 5, 2021 to May 29, 2024.

Supplementary search period: May 29, 2024 to July 23, 2024.

**Condition or domain being studied**

Post-Coronavirus Disease 2019 (COVID-19) condition or long COVID

**Participants/population**

Individuals who are diagnosed with COVID-19 are being studied by the review. We want to compare those who have long COVID and those who do not have long COVID.

**Interventions, exposure(s)**

The exposures would be long COVID symptoms occurred in individuals diagnosed with COVID-19.

**Comparator(s)/control**

The control would be individuals diagnosed with COVID-19 but do not have long COVID.

**Types of study to be included**

Cross-sectional, retrospective cohort, prospective cohort, ambidirectional cohort, case-control study

**Context**

Researchers are aware of long COVID as a complex multisystemic disease with numerous subtypes. There can be differences between prevalence of each subtype, and there can be updated risk factors for long COVID. Upon back ground research, there is no review concerning global prevalence of a well-curated list of long COVID subtypes with consideration of a large set of symptoms and risk factors for long COVID.

**Main outcome(s)**

Prevalence of long COVID, its subtypes and symptoms

Risk factors for long COVID

*Measures of effect*

Prevalence proportion and odds ratio/risk ratio

**Inclusion and exclusion criteria**

*Inclusion*

- Human study population with confirmed COVID-19 diagnosis through polymerase chain reaction (PCR) test, antibody test, or a clinical diagnosis
- Index date of first test/diagnosis, date of hospitalization, discharge date, date of clinical recovery/negative test, or date of symptom appearance
- Primary outcome must include prevalence, risk factors, duration, subtypes, or symptoms of long COVID.
- WHO definition of long COVID: continuation or development of new symptoms 3 months after the initial SARS-CoV-2 infection, with these symptoms lasting for at least 2 months with no other explanation

*Exclusion*

- Case studies, reviews, studies with imaging or molecular and/or cellular testing as primary results, and studies with only healthcare workers or residents of nursing homes and/or long-term care facilities
- Studies that estimated overall prevalence of long COVID did not meet the sample size threshold of 323, pre-calculated to ensure the included studies were adequately powered to achieve a margin of error of at least 0.05 with a priori’ prevalence of 30%.

**Data extraction (selection and coding)**

The studies will be screened based on the inclusion and exclusion criteria. Data extraction will be performed using the Covidence screening tool and spreadsheet. Two reviewers will independently screen records for inclusion in title and abstract screening and full-text screening. With any conflicts, two screeners will together conclude a decision.

Data about study design, sample size, methodology, prevalence estimates, variables of interests and risk factors will be extracted from study documents. Two people will also independently extract and check received data. Missing data will be reported for each study. Data will be stored in excel spreadsheets.

**Risk of bias (quality) assessment**

The risk of bias will be assessed based on the Joanna Briggs Institute (JBI) tool and further discussion. A funnel plot for examination of publication bias among included studies and Egger’s and Begg’s tests for funnel plot asymmetry will be shown.

**Strategy for data synthesis**

We included the reported measure from studies with no overlap between patient cohorts. If there were studies with overlapping cohorts, these studies can be included in the meta-analysis as long as they are reporting different outcomes. In the instance of overlapping cohorts on the same outcome measure, we chose one study, and following Chen et al. we preferred studies with the following characteristics: 1) being published more recently, 2) multicenter, 3) longer follow-up time, and 4) lower risk of bias.

Heterogeneity among studies was reflected by the

$$I^{2}$$

statistic, where

$$I^{2}$$

statistics between 70% and 100% indicated considerable heterogeneity. Random effects model with logit transformation and the DerSimonian-Laird estimator for between-study variance would be used to estimate pooled prevalence of long COVID and its subtypes and symptoms. Confidence intervals would be calculated by incorporating between-study variance obtained by the DerSimonian-Laird estimator. Odds ratio for individual studies would also be pooled using a random effects meta-analysis framework. All analysis would be conducted in **R** using packages **meta** and **metafor**.

**Type and method of review**

Meta-analysis, Systematic Review

**Actual start date**

May 6, 2024

**Anticipated completion date**

September, 2024

**Subject index terms**

COVID-19, Infectious Disease, Humans, Post-Acute COVID-19 Syndrome, Prevalence, Risk Factors, SARS-CoV-2
