## Supplementary material for "Global Prevalence of Long COVID, its Subtypes and Risk factors: An Updated Systematic Review and Meta-Analysis": eMethods 1b. Final search strategy

**eMethods 1**. Systematic review search strategies

PubMed

**Date searched:** 05/29/2024

**Number of results:** 5,117

**Date filter:** 07/05/2021 – present

Search blocks

1. "COVID-19 sequela*" OR (("COVID-19" OR "Sars-CoV-2" OR "2019 Novel Coronavirus" OR "2019-nCoV" OR "Coronavirus Disease 2019" OR "Coronavirus Disease-19" OR "SARS Coronavirus 2" OR "Severe Acute Respiratory Syndrome Coronavirus 2") AND sequela*) OR "post acute sequelae of Sars-CoV-2" OR ("PASC" AND ("COVID-19" OR "Sars-CoV-2" OR "2019 Novel Coronavirus" OR "2019-nCoV" OR "Coronavirus Disease 2019" OR "Coronavirus Disease 19" OR "SARS Coronavirus 2" OR "Severe Acute Respiratory Syndrome Coronavirus 2")) OR "post acute sequelae of COVID" OR (("post-intensive care syndrome" OR "postintensive care syndrome") AND ("COVID-19" OR "Sars-CoV-2" OR "2019 Novel Coronavirus" OR "2019-nCoV" OR "Coronavirus Disease 2019" OR "Coronavirus Disease-19" OR "SARS Coronavirus 2" OR "Severe Acute Respiratory Syndrome Coronavirus 2")) OR "post COVID condition*" OR "post COVID-19 condition*"[tw] OR "post COVID-19 condition*"[tw] OR ("PCC" AND ("COVID-19" OR "Sars-CoV-2" OR "2019 Novel Coronavirus" OR "2019-nCoV" OR "Coronavirus Disease 2019" OR "Coronavirus Disease-19" OR "SARS Coronavirus 2" OR "Severe Acute Respiratory Syndrome Coronavirus 2")) OR "convalescent COVID-19" OR "long haul COVID" OR "COVID long haul*" OR "long COVID" OR "long term COVID" OR "COVID-19 survivor*" OR "post COVID 19 symptom*" OR "chronic COVID syndrome" OR "post COVID syndrome" OR "post COVID-19 neurological syndrome" OR "post acute COVID-19" OR "post-acute COVID-19 syndrome"[mesh] OR "COVID-19 post-intensive care syndrome"[Supplementary Concept]

2. “prevalent”[tw] OR “prevalence”[tw] OR “prevalence”[mh] OR “occurrence”[tw] OR “occurrences”[tw] OR “duration”[tw] OR “durations”[tw] OR “length”[tw] OR “lengths”[tw] OR “risk factor”[tw] OR “risk factors”[tw] OR “risk factor”[title/abstract:~3] OR “risk factors”[title/abstract:~3] OR “Risk Factors”[mh] OR “predict”[tw] OR “prediction”[tw] OR “predictions”[tw] OR “predicting”[tw] OR “predictive”[tw] OR “predictor”[tw] OR “predictors”[tw] OR “symptom”[tw] OR “symptoms”[tw] OR “define”[tw] OR “defining”[tw] OR “definition”[tw] OR “definitions”[tw] OR “follow up”[tw] OR “follow-up”[tw] OR “followed up”[tw]

3. “Nervous System Diseases”[Mesh:NoExp] OR “Cerebrovascular Disorders”[Mesh:NoExp] OR “Brain Diseases”[Mesh:NoExp] OR “abnormal cerebrospinal fluid”[tw] OR (“abnormal”[tw] AND (“spinal fluid”[tw] OR “brain fluid”[tw])) OR “abnormal movement*”[tw] OR “dyskinesia”[tw] OR “dystonia”[tw] OR “dysautonomia”[tw] OR (“autonomic”[tw] AND (“dysfunction”[tw] OR “neuropathy”[tw])) OR “orthostatic hypotension”[tw] OR “blurred vision”[tw] OR “refractive error*”[tw] OR “brain hypometabolism”[tw] OR “brain glucose hypometabolism”[tw] OR “concentration*”[tw] OR “confusion*”[tw] OR “brain fog”[tw] OR “mental fog”[tw] OR “headache*”[tw] OR “cephalalgia”[tw] OR “migraine*”[tw] OR (“hearing”[tw] AND (“impairment”[tw] OR “loss”[tw]))OR “memory problem*”[tw] OR “memory loss”[tw] OR “amnesia”[tw] OR “amnestic syndrome”[tw] OR “dementia”[tw] OR “cellular dysregulation*”[tw] OR “neurogenesis”[tw] OR “neural dysregulation”[tw] OR (“myelin”[tw] AND (“loss”[tw] OR “dysregulation”[tw] OR “damage”[tw])) OR “demyelinating disease*”[tw] OR “nerve pain*”[tw] OR “neuralgia”[tw] OR “neuropathic pain”[tw] OR “neural inflammation*”[tw] OR “neuroimaging change*”[tw] OR “numbness”[tw] OR “tingling”[tw] OR “paresthesia”[tw] OR (“oxidative”[tw] AND (“stress”[tw] OR “damage”[tw])) OR “paralysis”[tw] OR “plegia”[tw] OR “paresis”[tw] OR “post-exertional malaise”[tw] OR “PEM”[tw] OR “chronic fatigue syndrome”[tw] OR “myalgic encephalomyelitis”[tw] OR “malaise”[tw] OR "grey matter reduction"[Title/Abstract:~3] OR "gray matter

reduction"[Title/Abstract:~3] OR "grey matter reductions"[Title/Abstract:~3] OR "gray matter reductions"[Title/Abstract:~3] OR “cortical atrophy”[tw] OR “seizure*”[tw] OR “sensorimotor symptom*”[tw] OR “sensory neuropathy”[tw] OR ((“balance”[tw] OR “gait”[tw]) AND

“problem*”[tw]) “serotonin”[tw] OR “serotonin reduction”[Title/Abstract:~3] OR “serotonin reductions”[Title/Abstract:~3] OR “sleep problem*”[tw] OR “sleep-wake disorder*”[tw] OR “narcolepsy”[tw] OR “smell”[tw] OR “anosmia”[tw] OR “hyposmia”[tw] OR “taste”[tw] OR “ageusia” OR “hypogeusia”[tw] OR “stroke*”[tw] OR “brain attack*”[tw] OR “tinnitus”[tw] OR “tremor*”[tw] OR “trembling”[tw]

4. “Anxiety Disorders”[Mesh:NoExp] OR “Depression”[Mesh:NoExp] OR “Mental Disorders”[Mesh:NoExp] OR “Mental Health”[Mesh] OR “anxiety”[tw] OR “anxiety disorder*”[tw] OR “anxious”[tw] OR “generalized anxiety disorder*”[tw] OR ((“panic”[tw] OR “anxiety”[tw]) AND “attack*”[tw]) OR “social anxiety disorder*”[tw] OR “fear”[tw] OR “phobia*”[tw] OR “attention deficit*”[tw] OR “hyperactive”[tw] OR “attention-deficit hyperactivity disorder”[tw] OR “ADHD”[tw] OR “inattention”[tw] OR “impulsivity”[tw] OR “delirium”[tw] OR “delusion*”[tw] OR “instability”[tw] OR “depress*”[tw] OR “insomnia”[tw] OR “agrypnia”[tw] OR “irritability”[tw] OR “dysregulation*”[tw] OR “post-traumatic stress disorder”[tw] OR “PTSD”[tw] OR “psychiatric disorder*”[tw] OR “sleep disturbance*”[tw] OR “psychomotor”[tw] OR “addiction*”[tw] OR “dependence”[tw] OR “substance abuse”[tw] OR “trauma”[tw] OR “stress”[tw] OR “mental disorder*”[tw]

5. “Cardiovascular System”[Mesh:NoExp] OR “Cardiovascular Abnormalities”[Mesh:NoExp] OR “anemia”[tw] OR “iron deficiency”[tw] OR “hemoglobin*”[tw] OR “anaemia”[tw] OR “arrhythmia*”[tw] OR “dysrhythmia”[tw] OR ((“abnormal”[tw] OR “irregular”[tw]) AND “heartbeat*”[tw]) OR “bleeding”[tw] OR “hemorrhage*”[tw] OR “damaged blood vessel*”[tw] OR “blood cell alteration”[Title/Abstract:~3] OR “poikilocytosis”[tw] OR “anisocytosis”[tw] OR “bradycardia”[tw] OR (“cardiac”[tw] AND (“impairment”[tw] OR “problem*”[tw] OR “arrest”[tw] OR “insufficiency”[tw] OR “abnormal*”[tw])) OR “heart failure*”[tw] OR “chest pain*”[tw] OR “angina”[tw] OR “clotting”[tw] OR “coagulation”[tw] OR “embolism”[tw] OR “thromboembolism”[tw] OR “coronary atherosclerosis”[tw] OR “atherosclerosis”[tw] OR “coronary artery disease*”[tw] OR “coronary heart disease*”[tw] OR “ischemia”[tw] OR “deep vein thrombosis”[tw] OR “endothelial inflammation”[tw] OR “endothelial dysfunction”[tw] OR “microthrombosis”[tw] OR “heart attack*”[tw] OR “hypertension”[tw] OR “blood pressure”[tw] OR “microangiopathy”[tw] OR “myocardial infarction”[tw] OR “myocardial inflammation”[tw] OR “palpitation*”[tw] OR “postural orthostatic tachycardia syndrome”[tw] OR “POTS”[tw] OR “tachycardia”[tw] OR “tachyarrhythmia”[tw]

6. “Cough”[Mesh] OR “Lung”[Mesh:NoExp] OR “Respiratory System”[Mesh:NoExp] OR “Respiratory System Abnormalities”[Mesh:NoExp] OR “Respiratory Tract Infections”[Mesh:NoExp] OR (“abnormal”[tw] AND (“gas exchange*”[tw] OR “respiration*”[tw])) OR “hyperventilation”[tw] OR “hyper-ventilation”[tw] OR “diffusion*”[tw] OR “asthma”[tw] OR “pleurisy”[tw] OR “pleura”[tw] OR “pleuritis”[tw] OR “chest pain*”[tw] OR “chest discomfort”[tw] OR “chronic obstructive pulmonary disease”[tw] OR “COPD”[tw] OR “emphysema”[tw] OR “bronchitis”[tw] OR “inflammatory lung disease”[tw] OR “cough”[tw] OR “chronic cough”[tw] OR “hyperpnea”[tw] OR “breathlessness”[tw] OR “dyspnea”[tw] OR “shortness of breath”[tw] OR “hypoxaemia”[tw] OR “hypoxia”[tw] OR “blood oxygen”[tw] OR “diffusion capacity”[tw] OR “lower respiratory disease*”[tw] OR “respiratory tract infection*” OR “pneumonia”[tw] OR “abscess”[tw] OR “nasal congestion”[tw] OR “rhinitis”[tw] OR “rhinorrhea”[tw] OR “sinus infection*”[tw] OR “nasal inflammation”[tw] OR “pulmonary embolism”[tw] OR “sleep apnea”[tw] OR “obstructive sleep apnea”[tw] OR “sleep disordered breathing”[tw] OR “airway resistance”[tw]

7. “Gastrointestinal Diseases”[Mesh:NoExp] OR “Gastrointestinal Microbiome”[Mesh] OR “abdominal pain”[tw] OR “stomachache*”[tw] OR “stomach ache*”[tw] OR “visceral pain”[tw] OR “peritoneal pain”[tw] OR “dyschezia”[tw] OR “dyssynergia”[tw] OR “constipation”[tw] OR “diarrhea”[tw] OR “gastroenteritis”[tw] OR “dry mouth”[tw] OR “xerostomia”[tw] OR “parageusia”[tw] OR “dysgeusia”[tw] OR “dyspepsia”[tw] OR “indigestion”[tw] OR “upset stomach”[tw] OR “achalasia”[tw] OR “esophagitis”[tw] OR “esophageal disorder*”[tw] OR “gastritis”[tw] OR “duodenitis”[tw] OR “gastroesophageal reflux disease”[tw] OR “GERD” OR “acid reflux”[tw] OR “heartburn*”[tw] OR (“acid”[tw] AND (“regurgitation”[tw] OR “indigestion”[tw])) OR “gastrointestinal disorder*”[tw] OR “digestive disorder*” OR “hemorrhoid*”[tw] OR “gastrointestinal

disturbance*”[tw] OR ((“gut”[tw] OR “microbiome”[tw] OR “gastrointestinal”[tw] OR “intestinal”[tw]) AND “dysbiosis”[tw]) OR “mouth pain”[tw] OR “orofacial pain”[tw] OR “emesis”[tw] OR “qualm”[tw] OR “nausea”[tw] OR “vomiting”[tw] OR “toothache*”[tw] OR “dental pain”[tw] OR “tooth decay”[tw] OR (“dental”[tw] AND (“caries”[tw] OR “cavity”[tw]))

8. “Kidney Diseases”[Mesh:NoExp] OR “Urologic Diseases”[Mesh:NoExp] OR (“bladder”[tw] AND (“problem*”[tw] OR “irritation*”[tw] OR “inflammation”[tw] OR “pain”)) OR “interstitial cystitis”[tw] OR “urinary incontinence”[tw] OR “cystitis”[tw] OR “chronic kidney disease”[tw] OR “renal disease*”[tw] OR “nephropathy”[tw] OR ((“kidney”[tw] OR “renal”[tw]) AND (“impairment”[tw] OR “insufficiency”[tw] OR “dysfunction”[tw])) OR “nephritis”[tw] OR “nephrosis”[tw] OR “hypovolaemia”[tw] OR “hyperosmolar”[tw] OR “hyponatremia”[tw] OR “dehydration”[tw] OR “fluid disorders”[Title/Abstract:~3] OR “fluid disorder”[Title/Abstract:~3] OR “electrolyte imbalance”[tw] OR “fluid imbalance”[tw] OR “diabetes insipidus”[tw] OR “kidney injury”[tw] OR “kidney injuries”[tw] OR “renal failure*”[tw]

9. “Musculoskeletal Abnormalities”[Mesh:NoExp] OR “Musculoskeletal Diseases”[Mesh:NoExp] OR “Musculoskeletal Pain”[Mesh] OR “connective tissue disease”[tw] OR “collagenosis”[tw] OR “ligaments “[tw] OR “muscle fascia*”[tw] OR “collagen”[tw] OR “joint pain”[tw] OR “arthralgia”[tw] OR “polyarthralgia”[tw] OR “arthritis”[tw] OR “articular pain*”[tw] OR (“joint”[tw] AND (“pain”[tw] OR “stiffness”[tw])) OR “myasthenia”[tw] OR “asthenia”[tw] OR “muscle weakness*”[tw] OR “musculoskeletal pain”[tw] OR “lower back pain”[tw] OR “muscle ache*”[tw] OR “myalgia”[tw] OR “osteoarthritis”[tw] OR “osteoarthrosis”[tw] OR “degenerative joint disease”[tw] OR “spondylitis”[tw] OR “spondyloarthritis”[tw] OR “spondylopathies”[tw] OR “tendon disorder*”[tw] OR “synovial disorder*”[tw] OR “tendinopathy”[tw] OR “tenosynovitis”[tw] OR “tendinosis”[tw] OR “tendinitis”[tw]

10. “Skin Diseases”[Mesh:NoExp] OR “hair loss”[tw] OR “alopecia”[tw] OR “skin color*”[tw] OR “pigmentation alteration”[tw] OR “skin discoloration*”[tw] OR “hyperpigmentation”[tw] OR “hypopigmentation”[tw] OR “skin sensitivity”[tw] OR “allodynia”[tw] OR “skin pain”[tw] OR “skin rash”[tw] OR “skin change*”[tw] OR “dermatitis”[tw] OR “eczema”[tw] OR “exanthem”[tw] OR “skin inflammation”[tw]

11. “Diabetes Mellitus”[Mesh:NoExp] OR “Endocrine System Diseases”[Mesh:NoExp] OR “Metabolic Syndrome”[Mesh:NoExp] OR “diabetes”[tw] OR “diabetes mellitus”[tw] OR “dyslipidemia”[tw] OR “lipid disorder*”[tw] OR “abnormal lipid level*”[tw] OR “hypercholesterolemia”[tw] OR “hypocholesterolemia”[tw] OR “cholesterol level”[tw] OR “hormonal disorder*”[tw] OR “endocrine disorder*”[tw] OR “hormonal imbalance”[tw] OR “hyperlipidaemia”[tw] OR “metabolic disorder*”[tw] OR “mitochondrial dysfunction”[tw] OR “impaired metabolism”[tw] OR “metabolic syndrome”[tw] OR “insulin resistance syndrome”[tw] OR “dysmetabolic syndrome”[tw] OR “weight gain”[tw] OR “obesity”[tw] OR “pancreas injury”[tw] OR “pancreatitis”[tw] OR “pancreatic trauma”[tw] OR “reproduct*”[tw] OR “hormonal change*”[tw] OR “hormone deficit*”[tw] OR “thirst”[tw] OR “polydipsia”[tw]

12. “Autoimmune Diseases”[Mesh:NoExp] OR “Fatigue”[Mesh:NoExp] OR

“Inflammation”[Mesh:NoExp] OR “appetite”[tw] OR “anorexia”[tw] OR “autoimmunity”[tw] OR (“immune”[tw] AND (“dysregulation”[tw] OR “reactivity”[tw])) OR “autoimmune”[tw] OR “dizziness”[tw] OR “vertigo”[tw] OR “vertiginous”[tw] OR “lightheadedness”[tw] OR “tiredness”[tw] OR “fatigue”[tw] OR “lassitude”[tw] OR “drowsiness”[tw] OR “sleepiness”[tw] OR “pyrexia”[tw] OR “hyperpyrexia”[tw] OR “fever”[tw] OR “sweats”[tw] OR “diaphoresis”[tw] OR “perspiration”[tw] OR “shivering”[tw] OR “chills”[tw] OR “immunosupress*”[tw] OR “immunocompromised”[tw] OR “sore throat”[tw] OR “pharyngodynia”[tw] OR “pharyngitis”[tw] OR “itchy throat”[tw] OR “throat pain”[tw] OR “swelling”[tw] OR “edema”[tw] OR “anasarca”[tw] OR “distress”[tw] OR “unspecified pain*”[tw] OR (“viral”[tw] AND (“persistence”[tw] OR “reactivation”[tw] OR “inflammation”[tw]))

1 AND 2 AND (3 OR 4 OR 5 OR 6 OR 7 OR 8 OR 9 OR 10 OR 11 OR 12)

Embase

**Date searched:** 05/29/2024

**Number of results:** 3,136

**Date filter:** 07/05/2021 - present

**Other filters applied:** Embase ONLY,

Search blocks

1. 'covid-19 sequela*' OR (('covid 19' OR 'sars cov 2' OR '2019 novel coronavirus' OR '2019 ncov' OR 'coronavirus disease 2019' OR 'coronavirus disease-19' OR 'sars coronavirus 2' OR 'severe acute respiratory syndrome coronavirus 2') AND sequela*) OR 'post acute sequelae of sars-cov 2' OR (pasc AND ('covid 19' OR 'sars cov 2' OR '2019 novel coronavirus' OR '2019 ncov' OR

'coronavirus disease 2019' OR 'coronavirus disease-19' OR 'sars coronavirus 2' OR 'severe acute respiratory syndrome coronavirus 2')) OR 'post acute sequelae of covid' OR (('post-intensive care syndrome' OR 'postintensive care syndrome') AND ('covid 19' OR 'sars cov 2' OR '2019 novel coronavirus' OR '2019 ncov' OR 'coronavirus disease 2019' OR 'coronavirus disease-19' OR 'sars coronavirus 2' OR 'severe acute respiratory syndrome coronavirus 2')) OR 'post covid condition*' OR 'post-COVID-19 condition*' OR 'post COVID-19 condition*' OR (pcc AND ('covid 19' OR 'sars cov 2' OR '2019 novel coronavirus' OR '2019 ncov' OR 'coronavirus disease 2019' OR 'coronavirus disease-19' OR 'sars coronavirus 2' OR 'severe acute respiratory syndrome coronavirus 2')) OR 'convalescent covid-19' OR 'long haul covid' OR 'covid long haul*' OR 'long covid' OR 'long term covid' OR 'covid-19 survivor*' OR 'post covid-19 symptom*' OR 'chronic covid syndrome' OR 'post covid syndrome' OR 'post covid-19 neurological syndrome' OR 'post acute covid-19' OR 'long covid'/de OR 'post intensive care syndrome'/exp

2. prevalent:ti,ab,kw OR prevalence:ti,ab,kw OR 'prevalence'/exp OR 'prevalence' OR occurrence:ti,ab,kw OR occurrences:ti,ab,kw OR duration:ti,ab,kw OR durations:ti,ab,kw OR length:ti,ab,kw OR lengths:ti,ab,kw OR 'risk factor'/exp OR ((risk NEAR/3 factor*):ti,ab,kw) OR 'risk factor'/exp OR predict:ti,ab,kw OR prediction:ti,ab,kw OR predictions:ti,ab,kw OR predicting:ti,ab,kw OR predictive:ti,ab,kw OR predictor:ti,ab,kw OR predictors:ti,ab,kw OR define:ti,ab,kw OR defining:ti,ab,kw OR definition:ti,ab,kw OR definitions:ti,ab,kw OR 'follow up':ti,ab,kw OR 'followed up':ti,ab,kw

3. 'neurologic disease'/de OR 'cerebrovascular disease'/de OR 'brain disease'/de OR 'abnormal cerebrospinal fluid':ti,ab,kw OR (abnormal AND ('spinal fluid' OR 'brain fluid')):ti,ab,kw OR 'abnormal movement*':ti,ab,kw OR dyskinesia:ti,ab,kw OR dystonia:ti,ab,kw OR dysautonomia:ti,ab,kw OR (autonomic AND (dysfunction OR neuropathy)):ti,ab,kw OR 'orthostatic hypotension':ti,ab,kw OR 'blurred vision':ti,ab,kw OR 'refractive error*':ti,ab,kw OR 'brain hypometabolism':ti,ab,kw OR 'brain glucose hypometabolism':ti,ab,kw OR concentration*:ti,ab,kw OR confusion*:ti,ab,kw OR 'brain fog':ti,ab,kw OR 'mental fog':ti,ab,kw OR headache*:ti,ab,kw OR cephalalgia:ti,ab,kw OR migraine*:ti,ab,kw OR (hearing AND (impairment OR loss)):ti,ab,kw OR ‘memory problem*':ti,ab,kw OR 'memory loss':ti,ab,kw OR amnesia:ti,ab,kw OR 'amnestic syndrome':ti,ab,kw OR dementia:ti,ab,kw OR 'cellular dysregulation*':ti,ab,kw OR neurogenesis:ti,ab,kw OR 'neural dysregulation':ti,ab,kw OR (myelin AND (loss OR dysregulation OR damage)):ti,ab,kw OR 'demyelinating disease*':ti,ab,kw OR 'nerve pain*':ti,ab,kw OR neuralgia:ti,ab,kw OR 'neuropathic pain':ti,ab,kw OR 'neural inflammation*':ti,ab,kw OR 'neuroimaging change*':ti,ab,kw OR numbness:ti,ab,kw OR tingling:ti,ab,kw OR paresthesia:ti,ab,kw OR (oxidative AND (stress OR damage)):ti,ab,kw OR paralysis:ti,ab,kw OR plegia:ti,ab,kw OR paresis:ti,ab,kw OR 'post-exertional malaise':ti,ab,kw OR PEM:ti,ab,kw OR 'post-exertional autoimmune exhaustion':ti,ab,kw OR 'chronic fatigue syndrome':ti,ab,kw OR 'myalgic encephalomyelitis':ti,ab,kw OR malaise:ti,ab,kw OR ((‘grey matter’ NEAR/3 reduction):ti,ab,kw) OR ((‘gray matter’ NEAR/3 reduction):ti,ab,kw) OR ((‘gray matter’ NEAR/3 reductions):ti,ab,kw) OR ((‘grey matter’ NEAR/3 reductions):ti,ab,kw) OR 'cortical atrophy':ti,ab,kw OR seizure*:ti,ab,kw OR 'sensorimotor symptom*':ti,ab,kw OR 'sensory neuropathy':ti,ab,kw OR ((balance OR gait) AND problem*):ti,ab,kw serotonin:ti,ab,kw OR ((‘serotonin’ NEAR/3 reduction):ti,ab,kw) OR ((‘serotonin’ NEAR/3 reductions):ti,ab,kw) OR 'sleep problem*':ti,ab,kw

OR 'sleep-wake disorder*':ti,ab,kw OR narcolepsy:ti,ab,kw OR smell:ti,ab,kw OR anosmia:ti,ab,kw OR hyposmia:ti,ab,kw OR taste:ti,ab,kw OR ageusia:ti,ab,kw OR hypogeusia:ti,ab,kw OR stroke*:ti,ab,kw OR 'brain attack*':ti,ab,kw OR tinnitus:ti,ab,kw OR tremor*:ti,ab,kw OR trembling:ti,ab,kw

4. 'anxiety disorder'/de OR depression/de OR 'mental disease'/de OR 'mental health'/exp OR anxiety:ti,ab,kw OR 'anxiety disorder*':ti,ab,kw OR anxious:ti,ab,kw OR 'generalized anxiety disorder*':ti,ab,kw OR ((panic OR anxiety) AND attack*):ti,ab,kw OR 'social anxiety disorder*':ti,ab,kw OR fear:ti,ab,kw OR phobia*:ti,ab,kw OR 'attention deficit*':ti,ab,kw OR hyperactive:ti,ab,kw OR 'attention-deficit hyperactivity disorder':ti,ab,kw OR ADHD:ti,ab,kw OR inattention:ti,ab,kw OR impulsivity:ti,ab,kw OR delirium:ti,ab,kw OR delusion*:ti,ab,kw OR instability:ti,ab,kw OR depress*:ti,ab,kw OR insomnia:ti,ab,kw OR agrypnia:ti,ab,kw OR irritability:ti,ab,kw OR dysregulation*:ti,ab,kw OR 'post-traumatic stress disorder':ti,ab,kw OR PTSD:ti,ab,kw OR 'psychiatric disorder*':ti,ab,kw OR 'sleep disturbance*':ti,ab,kw OR psychomotor:ti,ab,kw OR addiction*:ti,ab,kw OR dependence:ti,ab,kw OR 'substance abuse':ti,ab,kw OR trauma:ti,ab,kw OR stress:ti,ab,kw OR 'mental disorder*':ti,ab,kw

5. 'cardiovascular system'/de OR 'cardiovascular disease'/de OR anemia:ti,ab,kw OR 'iron deficiency':ti,ab,kw OR hemoglobin*:ti,ab,kw OR anaemia:ti,ab,kw OR arrhythmia*:ti,ab,kw OR dysrhythmia:ti,ab,kw OR ((abnormal OR irregular) AND heartbeat*):ti,ab,kw OR bleeding:ti,ab,kw OR hemorrhage*:ti,ab,kw OR 'damaged blood vessel*':ti,ab,kw OR ('blood cell’ NEAR/3 alteration):ti,ab,kw OR poikilocytosis:ti,ab,kw OR 'anisocytosis’ OR ‘bradycardia':ti,ab,kw OR (cardiac AND (impairment OR problem*:ti,ab,kw OR arrest:ti,ab,kw OR insufficiency OR abnormal*)):ti,ab,kw OR 'heart failure*':ti,ab,kw OR 'chest pain*':ti,ab,kw OR angina:ti,ab,kw OR clotting:ti,ab,kw OR coagulation:ti,ab,kw OR embolism:ti,ab,kw OR thromboembolism:ti,ab,kw OR 'coronary atherosclerosis':ti,ab,kw OR atherosclerosis:ti,ab,kw OR 'coronary artery disease*':ti,ab,kw OR 'coronary heart disease*':ti,ab,kw OR ischemia:ti,ab,kw OR 'deep vein thrombosis':ti,ab,kw OR 'endothelial inflammation':ti,ab,kw OR 'endothelial dysfunction':ti,ab,kw OR microthrombosis:ti,ab,kw OR 'heart attack*':ti,ab,kw OR hypertension:ti,ab,kw OR 'blood pressure':ti,ab,kw OR microangiopathy:ti,ab,kw OR 'myocardial infarction':ti,ab,kw OR 'myocardial inflammation':ti,ab,kw OR palpitation*:ti,ab,kw OR postural:ti,ab,kw OR ‘orthostatic tachycardia syndrome':ti,ab,kw OR POTS:ti,ab,kw OR tachycardia:ti,ab,kw OR tachyarrhythmia:ti,ab,kw

6. coughing/de OR lung/de OR 'respiratory system'/de OR 'respiratory system abnormalities'/de OR 'respiratory tract disease'/de OR (abnormal AND ('gas exchange*’ OR respiration*)) OR hyperventilation:ti,ab,kw OR hyper-ventilation:ti,ab,kw OR diffusion*:ti,ab,kw OR asthma:ti,ab,kw OR pleurisy:ti,ab,kw OR pleura:ti,ab,kw OR pleuritis:ti,ab,kw OR 'chest pain*':ti,ab,kw OR 'chest discomfort':ti,ab,kw OR 'chronic obstructive pulmonary disease':ti,ab,kw OR COPD:ti,ab,kw OR emphysema:ti,ab,kw OR bronchitis:ti,ab,kw OR 'inflammatory lung disease':ti,ab,kw OR cough:ti,ab,kw OR 'chronic cough':ti,ab,kw OR hyperpnea:ti,ab,kw OR breathlessness:ti,ab,kw OR dyspnea:ti,ab,kw OR 'shortness of breath':ti,ab,kw OR hypoxaemia:ti,ab,kw OR hypoxia:ti,ab,kw OR 'blood oxygen':ti,ab,kw OR 'diffusion capacity':ti,ab,kw OR 'lower respiratory disease*':ti,ab,kw OR 'respiratory tract infection*':ti,ab,kw OR pneumonia:ti,ab,kw OR abscess:ti,ab,kw OR 'nasal congestion':ti,ab,kw OR rhinitis:ti,ab,kw OR rhinorrhea:ti,ab,kw OR 'sinus infection*':ti,ab,kw OR 'nasal inflammation':ti,ab,kw OR 'pulmonary embolism':ti,ab,kw OR 'sleep apnea':ti,ab,kw OR 'obstructive sleep apnea':ti,ab,kw OR 'sleep disordered breathing':ti,ab,kw OR 'airway resistance':ti,ab,kw

7. 'gastrointestinal disease'/de OR 'intestine flora'/exp OR 'abdominal pain’:ti,ab,kw OR ‘stomachache*':ti,ab,kw OR 'stomach ache*':ti,ab,kw OR 'visceral pain':ti,ab,kw OR 'peritoneal pain':ti,ab,kw OR dyschezia:ti,ab,kw OR dyssynergia:ti,ab,kw OR constipation:ti,ab,kw OR diarrhea:ti,ab,kw OR gastroenteritis:ti,ab,kw OR 'dry mouth':ti,ab,kw OR xerostomia:ti,ab,kw OR parageusia:ti,ab,kw OR dysgeusia:ti,ab,kw OR dyspepsia:ti,ab,kw OR indigestion:ti,ab,kw OR 'upset stomach':ti,ab,kw OR achalasia:ti,ab,kw OR esophagitis:ti,ab,kw OR 'esophageal disorder*':ti,ab,kw OR gastritis:ti,ab,kw OR duodenitis:ti,ab,kw OR 'gastroesophageal reflux disease':ti,ab,kw OR GERD:ti,ab,kw OR 'acid reflux':ti,ab,kw OR heartburn*:ti,ab,kw OR (acid

AND (regurgitation OR indigestion)):ti,ab,kw OR 'gastrointestinal disorder*':ti,ab,kw OR 'digestive disorder*':ti,ab,kw OR hemorrhoid*:ti,ab,kw OR 'gastrointestinal disturbance*':ti,ab,kw OR ((gut OR microbiome OR gastrointestinal OR intestinal) AND dysbiosis):ti,ab,kw OR 'mouth pain':ti,ab,kw OR 'orofacial pain':ti,ab,kw OR emesis:ti,ab,kw OR qualm:ti,ab,kw OR nausea:ti,ab,kw OR vomiting:ti,ab,kw OR toothache*:ti,ab,kw OR 'dental pain':ti,ab,kw OR 'tooth decay':ti,ab,kw OR (dental AND (caries OR cavity)):ti,ab,kw

8. 'kidney disease'/de OR 'urinary tract disease'/de OR (bladder AND (problem*:ti,ab,kw OR irritation* OR inflammation OR pain)):ti,ab,kw OR 'irritated bladder':ti,ab,kw OR 'interstitial cystitis':ti,ab,kw OR 'urinary incontinence':ti,ab,kw OR cystitis:ti,ab,kw OR 'chronic kidney disease':ti,ab,kw OR 'renal disease*':ti,ab,kw OR nephropathy:ti,ab,kw OR ((kidney OR renal)

AND (impairment OR insufficiency OR dysfunction)):ti,ab,kw OR nephritis:ti,ab,kw OR nephrosis:ti,ab,kw OR hypovolaemia:ti,ab,kw OR hyperosmolar:ti,ab,kw OR

hyponatremia:ti,ab,kw OR dehydration:ti,ab,kw OR ((fluid NEAR/3 disorders):ti,ab,kw) OR ((fluid NEAR/3 disorder):ti,ab,kw) OR 'electrolyte imbalance':ti,ab,kw OR 'fluid imbalance':ti,ab,kw OR 'diabetes insipidus':ti,ab,kw OR 'kidney injury':ti,ab,kw OR 'kidney injuries':ti,ab,kw OR 'renal failure*':ti,ab,kw

9. 'musculoskeletal system malformation'/de OR 'musculoskeletal disease'/de OR 'musculoskeletal pain'/exp OR 'connective tissue disease':ti,ab,kw OR collagenosis:ti,ab,kw OR ligaments:ti,ab,kw OR 'muscle fascia*':ti,ab,kw OR collagen:ti,ab,kw OR 'joint pain':ti,ab,kw OR arthralgia OR polyarthralgia:ti,ab,kw OR arthritis:ti,ab,kw OR 'articular pain*':ti,ab,kw OR (joint AND (pain OR stiffness)):ti,ab,kw OR myasthenia:ti,ab,kw OR asthenia:ti,ab,kw OR 'muscle weakness*':ti,ab,kw OR 'musculoskeletal pain':ti,ab,kw OR 'lower back pain':ti,ab,kw OR 'muscle ache*':ti,ab,kw OR myalgia:ti,ab,kw OR osteoarthritis:ti,ab,kw OR osteoarthrosis:ti,ab,kw OR 'degenerative joint disease':ti,ab,kw OR spondylitis:ti,ab,kw OR spondyloarthritis:ti,ab,kw OR

spondylopathies:ti,ab,kw OR 'tendon disorder*':ti,ab,kw OR 'synovial disorder*':ti,ab,kw OR tendinopathy:ti,ab,kw OR tenosynovitis:ti,ab,kw OR tendinosis:ti,ab,kw OR tendinitis:ti,ab,kw

10. 'skin disease'/de OR 'hair loss':ti,ab,kw OR alopecia:ti,ab,kw OR 'skin color*':ti,ab,kw OR 'pigmentation alteration':ti,ab,kw OR 'skin discoloration*':ti,ab,kw OR hyperpigmentation:ti,ab,kw OR hypopigmentation:ti,ab,kw OR 'skin sensitivity':ti,ab,kw OR allodynia:ti,ab,kw OR 'skin pain':ti,ab,kw OR 'skin rash':ti,ab,kw OR 'skin change*':ti,ab,kw OR dermatitis:ti,ab,kw OR eczema:ti,ab,kw OR exanthem:ti,ab,kw OR 'skin inflammation':ti,ab,kw

11. 'diabetes mellitus'/de OR 'endocrine disease'/de OR 'metabolic syndrome X'/de OR diabetes:ti,ab,kw OR 'diabetes mellitus':ti,ab,kw OR dyslipidemia:ti,ab,kw OR 'lipid disorder*':ti,ab,kw OR 'abnormal lipid level*’:ti,ab,kw OR hypercholesterolemia:ti,ab,kw OR hypocholesterolemia:ti,ab,kw OR 'cholesterol level':ti,ab,kw OR 'hormonal disorder*':ti,ab,kw OR 'endocrine disorder*':ti,ab,kw OR 'hormonal imbalance':ti,ab,kw OR hyperlipidaemia:ti,ab,kw OR 'metabolic disorder*':ti,ab,kw OR 'mitochondrial dysfunction':ti,ab,kw OR 'impaired metabolism':ti,ab,kw OR 'metabolic syndrome':ti,ab,kw OR 'insulin resistance syndrome':ti,ab,kw OR 'dysmetabolic syndrome':ti,ab,kw OR 'weight gain':ti,ab,kw OR obesity:ti,ab,kw OR 'pancreas injury':ti,ab,kw OR pancreatitis:ti,ab,kw OR 'pancreatic trauma':ti,ab,kw OR reproduct*:ti,ab,kw OR 'hormonal change*':ti,ab,kw OR 'hormone deficit*':ti,ab,kw OR thirst:ti,ab,kw OR polydipsia:ti,ab,kw

12. 'autoimmune disease'/de OR fatigue/de OR inflammation/de OR appetite:ti,ab,kw OR anorexia:ti,ab,kw OR autoimmunity:ti,ab,kw OR (immune AND (dysregulation OR reactivity)):ti,ab,kw OR autoimmune:ti,ab,kw OR dizziness:ti,ab,kw OR vertigo:ti,ab,kw OR vertiginous:ti,ab,kw OR lightheadedness:ti,ab,kw OR tiredness:ti,ab,kw OR fatigue:ti,ab,kw OR lassitude:ti,ab,kw OR drowsiness:ti,ab,kw OR sleepiness:ti,ab,kw OR pyrexia:ti,ab,kw OR hyperpyrexia:ti,ab,kw OR fever:ti,ab,kw OR sweats:ti,ab,kw OR diaphoresis:ti,ab,kw OR perspiration:ti,ab,kw OR shivering:ti,ab,kw OR chills:ti,ab,kw OR immunosupress*:ti,ab,kw OR immunocompromised:ti,ab,kw OR 'sore throat':ti,ab,kw OR pharyngodynia:ti,ab,kw OR pharyngitis:ti,ab,kw OR 'itchy throat':ti,ab,kw OR 'throat pain':ti,ab,kw OR swelling:ti,ab,kw OR

edema:ti,ab,kw OR anasarca:ti,ab,kw OR distress:ti,ab,kw OR 'unspecified pain*':ti,ab,kw OR (viral AND (persistence OR reactivation OR inflammation)):ti,ab,kw

1 AND 2 AND (3 OR 4 OR 5 OR 6 OR 7 OR 8 OR 9 OR 10 OR 11 OR 12)

Web of Science Core Collection

**Date searched:** 05/29/2024

**Number of results:** 5,161

**Date filter:** 07/05/2021 - present

**Other filters applied:** Search in TOPIC field

Search blocks

1. "COVID-19 sequela*" OR ((COVID-19 OR Sars-CoV-2 OR "2019 Novel Coronavirus" OR 2019- nCoV OR "Coronavirus Disease 2019" OR "Coronavirus Disease-19" OR "SARS Coronavirus 2" OR "Severe Acute Respiratory Syndrome Coronavirus 2" ) AND sequela* ) OR "post acute sequelae of Sars-CoV-2" OR (PASC AND (COVID-19 OR Sars-CoV-2 OR "2019 Novel Coronavirus" OR 2019-nCoV OR "Coronavirus Disease 2019" OR "Coronavirus Disease-19" OR "SARS Coronavirus 2" OR "Severe Acute Respiratory Syndrome Coronavirus 2" )) OR "post acute sequelae of COVID" OR (("post-intensive care syndrome" OR "postintensive care syndrome" ) AND (COVID-19 OR Sars-CoV-2 OR "2019 Novel Coronavirus" OR 2019-nCoV OR "Coronavirus Disease 2019" OR "Coronavirus Disease-19" OR "SARS Coronavirus 2" OR "Severe Acute Respiratory Syndrome Coronavirus 2" )) OR "post COVID condition*" OR "post COVID-19 condition*" OR "post COVID-19 condition*" OR (PCC AND (COVID-19 OR Sars-CoV 2 OR "2019 Novel Coronavirus" OR 2019-nCoV OR "Coronavirus Disease 2019" OR "Coronavirus Disease-19" OR "SARS Coronavirus 2" OR "Severe Acute Respiratory Syndrome Coronavirus 2" )) OR "convalescent COVID-19" OR "long haul COVID" OR "COVID long haul*" OR "long COVID" OR "long term COVID" OR "COVID-19 survivor*" OR "post COVID-19 symptom*" OR "chronic COVID syndrome" OR "post COVID syndrome" OR "post COVID-19 neurological syndrome" OR "post acute COVID-19" OR "post-acute COVID-19 syndrome" OR "COVID-19 post-intensive care syndrome"

2. prevalent OR prevalence OR prevalence OR occurrence OR occurrences OR duration OR durations OR length OR lengths OR "risk factor" OR "risk factors" OR "risk NEAR/3 factor” OR "risk NEAR/3 factors” OR "Risk Factors" OR predict OR prediction OR predictions OR predicting OR predictive OR predictor OR predictors OR symptom OR symptoms OR define OR defining OR definition OR definitions OR "follow up" OR follow-up OR "followed up"

3. "Nervous System Diseases" OR "Cerebrovascular Disorders" OR "Brain Diseases" OR "abnormal cerebrospinal fluid" OR (abnormal AND ("spinal fluid" OR "brain fluid")) OR "abnormal movement*" OR dyskinesia OR dystonia OR dysautonomia OR (autonomic AND (dysfunction OR neuropathy)) OR "orthostatic hypotension" OR "blurred vision" OR "refractive error*" OR "brain hypometabolism" OR "brain glucose hypometabolism" OR concentration* OR confusion* OR "brain fog" OR "mental fog" OR headache* OR cephalalgia OR migraine* OR (hearing AND (impairment OR loss)) OR “memory problem*” OR "memory loss" OR amnesia OR "amnestic syndrome" OR dementia OR "cellular dysregulation*" OR neurogenesis OR "neural dysregulation" OR (myelin AND (loss OR dysregulation OR damage)) OR "demyelinating disease*" OR "nerve pain*" OR neuralgia OR "neuropathic pain" OR "neural inflammation*" OR "neuroimaging change*" OR numbness OR tingling OR paresthesia OR (oxidative AND (stress OR damage)) OR paralysis OR plegia OR paresis OR "post-exertional malaise" OR PEM OR "post-exertional autoimmune exhaustion" OR "chronic fatigue syndrome" OR "myalgic encephalomyelitis" OR malaise OR "grey matter NEAR/3 reduction" OR "gray matter NEAR/3

reduction" OR "grey matter NEAR/3 reductions" OR "gray matter NEAR/3 reductions" OR "cortical atrophy" OR seizure* OR "sensorimotor symptom*" OR "sensory neuropathy" OR ((balance OR gait) AND problem*) OR serotonin OR "serotonin NEAR/3 reduction" OR "serotonin NEAR/3 reductions" OR "sleep problem*" OR "sleep-wake disorder*" OR narcolepsy OR smell OR anosmia OR hyposmia OR taste OR ageusia OR hypogeusia OR stroke* OR "brain attack*" OR tinnitus OR tremor* OR trembling

4. "Anxiety Disorders" OR Depression OR "Mental Disorders" OR "Mental Health" OR anxiety OR "anxiety disorder*" OR anxious OR "generalized anxiety disorder*" OR ((panic OR anxiety) AND attack*) OR "social anxiety disorder*" OR fear OR phobia* OR "attention deficit*" OR hyperactive OR "attention-deficit hyperactivity disorder" OR ADHD OR inattention OR impulsivity OR delirium OR delusion* OR instability OR depress* OR insomnia OR agrypnia OR irritability OR dysregulation OR "post-traumatic stress disorder" OR PTSD OR "psychiatric disorder*" OR "sleep disturbance*" OR psychomotor OR addiction* OR dependence OR "substance abuse" OR trauma OR stress OR "mental disorder*"

5. "Cardiovascular System" OR "Cardiovascular Abnormalities" OR anemia OR "iron deficiency" OR hemoglobin* OR anaemia OR arrhythmia* OR dysrhythmia OR ((abnormal OR irregular) AND heartbeat*) OR bleeding OR hemorrhage* OR "damaged blood vessel*" OR "blood cell NEAR/3 alteration" OR poikilocytosis OR anisocytosis OR bradycardia OR (cardiac AND (impairment OR problem* OR arrest OR insufficiency OR abnormal*)) OR "heart failure*" OR "chest pain*" OR angina OR clotting OR coagulation OR embolism OR thromboembolism OR "coronary atherosclerosis" OR atherosclerosis OR "coronary artery disease*" OR "coronary heart disease*" OR ischemia OR "deep vein thrombosis" OR "endothelial inflammation" OR "endothelial dysfunction" OR microthrombosis OR "heart attack*" OR hypertension OR "blood pressure" OR microangiopathy OR "myocardial infarction" OR "myocardial inflammation" OR palpitation* OR "postural orthostatic tachycardia syndrome" OR POTS OR tachycardia OR tachyarrhythmia

6. Cough OR Lung OR "Respiratory System" OR "Respiratory System Abnormalities" OR "Respiratory Tract Infections" OR (abnormal AND ("gas exchange*" OR respiration*)) OR hyperventilation OR hyper-ventilation OR diffusion* OR asthma OR pleurisy OR pleura OR pleuritis OR "chest pain*" OR "chest discomfort" OR "chronic obstructive pulmonary disease" OR COPD OR emphysema OR bronchitis OR "inflammatory lung disease" OR cough OR "chronic cough" OR hyperpnea OR breathlessness OR dyspnea OR "shortness of breath" OR hypoxaemia OR hypoxia OR "blood oxygen" OR "diffusion capacity" OR "lower respiratory disease*" OR "respiratory tract infection*" OR pneumonia OR abscess OR "nasal congestion" OR rhinitis OR rhinorrhea OR "sinus infection*" OR "nasal inflammation" OR "pulmonary embolism" OR "sleep apnea" OR "obstructive sleep apnea" OR "sleep disordered breathing" OR "airway resistance"

7. "Gastrointestinal Diseases" OR "Gastrointestinal Microbiome" OR "abdominal pain” “stomachache*” OR "stomach ache*" OR "visceral pain" OR "peritoneal pain" OR dyschezia OR dyssynergia OR constipation OR diarrhea OR gastroenteritis OR "dry mouth" OR xerostomia OR parageusia OR dysgeusia OR dyspepsia OR indigestion OR "upset stomach" OR achalasia OR esophagitis OR "esophageal disorder*" OR gastritis OR duodenitis OR "gastroesophageal reflux disease" OR GERD OR "acid reflux" OR heartburn* OR (acid AND (regurgitation OR indigestion)) OR "gastrointestinal disorder*" OR "digestive disorder*" OR hemorrhoid* OR "gastrointestinal disturbance*" OR ((gut OR microbiome OR gastrointestinal OR intestinal) AND dysbiosis) OR "mouth pain" OR "orofacial pain" OR emesis OR qualm OR nausea OR vomiting OR toothache* OR "dental pain" OR "tooth decay" OR (dental AND (caries OR cavity))

8. "Kidney Diseases" OR "Urologic Diseases" OR (bladder AND (problem* OR irritation* OR inflammation OR pain)) OR "irritated bladder" OR "interstitial cystitis" OR "urinary incontinence" OR cystitis OR "chronic kidney disease" OR "renal disease*" OR nephropathy OR ((kidney OR renal) AND (impairment OR insufficiency OR dysfunction)) OR nephritis OR nephrosis OR hypovolaemia OR hyperosmolar OR hyponatremia OR dehydration OR "fluid NEAR/3 disorder"

OR "fluid NEAR/3 disorders" OR "electrolyte imbalance" OR "fluid imbalance" OR "diabetes insipidus" OR "kidney injury" OR "kidney injuries" OR "renal failure*"

9. "Musculoskeletal Abnormalities" OR "Musculoskeletal Diseases" OR "Musculoskeletal Pain" OR "connective tissue disease" OR collagenosis OR ligaments OR "muscle fascia*" OR collagen OR "joint pain" OR arthralgia OR polyarthralgia OR arthritis OR "articular pain*" OR (joint AND (pain OR stiffness)) OR myasthenia OR asthenia OR "muscle weakness*" OR "musculoskeletal pain" OR "lower back pain" OR "muscle ache*" OR myalgia OR osteoarthritis OR osteoarthrosis OR "degenerative joint disease" OR spondylitis OR spondyloarthritis OR spondylopathies OR "tendon disorder*" OR "synovial disorder*" OR tendinopathy OR tenosynovitis OR tendinosis OR tendinitis

10. "Skin Diseases" OR "hair loss" OR alopecia OR "skin color*" OR "pigmentation alteration" OR "skin discoloration*" OR hyperpigmentation OR hypopigmentation OR "skin sensitivity" OR allodynia OR "skin pain" OR "skin rash" OR "skin change*" OR dermatitis OR eczema OR exanthem OR "skin inflammation"

11. "Diabetes Mellitus" OR "Endocrine System Diseases" OR "Metabolic Syndrome" OR diabetes OR "diabetes mellitus" OR dyslipidemia OR "lipid disorder*" OR "abnormal lipid level*" OR hypercholesterolemia OR hypocholesterolemia OR "cholesterol level" OR "hormonal disorder*" OR "endocrine disorder*" OR "hormonal imbalance" OR hyperlipidaemia OR "metabolic disorder*" OR "mitochondrial dysfunction" OR "impaired metabolism" OR "metabolic syndrome" OR "insulin resistance syndrome" OR "dysmetabolic syndrome" OR "weight gain" OR obesity OR "pancreas injury" OR pancreatitis OR "pancreatic trauma" OR "reproduct*” OR "hormonal change*" OR "hormone deficit*" OR thirst OR polydipsia

12. "Autoimmune Diseases" OR Fatigue OR Inflammation OR appetite OR anorexia OR autoimmunity OR (immune AND (dysregulation OR reactivity)) OR autoimmune OR dizziness OR vertigo OR vertiginous OR lightheadedness OR tiredness OR fatigue OR lassitude OR drowsiness OR sleepiness OR pyrexia OR hyperpyrexia OR fever OR sweats OR diaphoresis OR perspiration OR shivering OR chills OR immunosupress* OR immunocompromised OR "sore throat" OR pharyngodynia OR pharyngitis OR "itchy throat" OR "throat pain" OR swelling OR edema OR anasarca OR distress OR "unspecified pain*" OR (viral AND (persistence OR reactivation OR inflammation))

1 AND 2 AND (3 OR 4 OR 5 OR 6 OR 7 OR 8 OR 9 OR 10 OR 11 OR 12)
