## Supplementary material for "Global Prevalence of Long COVID, its Subtypes and Risk factors: An Updated Systematic Review and Meta-Analysis": eMethods 1c. Long COVID subtypes justification

**Long COVID Subtypes**, [link to spreadsheet for subtypes and symptoms/conditions](https://docs.google.com/spreadsheets/d/1TPMlDcZqZ3YB07h8cU_ivdSca_YzGH5VBKHeshVdnGI/edit?usp=sharing)

**ICD-11**, [International Classification of Disease 11th Revision](https://icd.who.int/browse/2024-01/mms/en)

**Publications** supporting each subtype

| **Subtype 1:**  Neurological or nervous system related | **Subtype 2:**  Psychological |
| --- | --- |
| **ICD-11:**  **06** Mental, behavior, or neurodevelopmental disorders  **07** Sleep-wake disorders  **09** Diseases of the nervous system | **ICD-11:**  **06** Mental, behavior, or neurodevelopmental disorders |
| **Summarization:**  Persistent neurological symptoms are found in patients with Long COVID, and ongoing brain damage is found in COVID-19 patients. These suggest that Long COVID can lead to long-term brain injury. Neurological conditions such as functional neurological disorders, changes in brain physiology, and various related symptoms constitute a subtype of nervous system diseases or disorders for Long COVID.  **Publications:**  Long COVID: major findings, mechanisms and recommendations (2023), [*Nature Reviews Microbiology*](https://www.nature.com/articles/s41579-022-00846-2)  Development of a Definition of Postacute Sequelae of SARS-CoV-2 Infection (2023), [*JAMA Network*](https://jamanetwork.com/journals/jama/fullarticle/2805540), [Supplementary Materials](https://cdn.jamanetwork.com/ama/content_public/journal/jama/939166/joi230062supp3_prod_1712695240.88184.pdf?Expires=1719182313&Signature=1DWb1eL9fHoDGO~Lajw5Y1vRNcYVX66275UgIbyhUjNHBvihGpYvk3BwSZ1zT45umhUYazDXV5H2wOvdPNyeVfPwJHvIzedWmin63WyDW9p~mtx3JaRUc5oJ3807SSIjOtN5sFtde0fboaLxMjjJCOn~Exkf2fOpGhZpGcnBnaXdEzimHjAB9TlbwteUnJJsHiZwUuADuW4YVXlSYG9sASDUMzPo2A-u1EFCM3bnDwUDc-Go48i1d9G0NDpcX8poU5WW5xBH8zOSyB1R228hA2ZL-AA7Nv-sXZ46ZGmDyC7PFXwzOGuAV59TjlAdHMy-Jz6QuwZpZUgyswmDcalmJQ__&Key-Pair-Id=APKAIE5G5CRDK6RD3PGA)  Defining the Subtypes of Long COVID and Risk Factors for Prolonged Disease: Population-Based Case-Crossover Study (2024), [*JMIR Publications*](https://publichealth.jmir.org/2024/1/e49841), [Suppl. Materials](https://docs.google.com/document/d/1Wte1k_dqrU9_YGrrS-j__qAoq8STjoKo/edit?usp=drive_link&ouid=107389695956544702385&rtpof=true&sd=true)  Prevalence and Trajectories of Post-COVID-19 Neurological Manifestations: A Systematic Review and Meta-Analysis (2024), [*Neuroepidemiology*](https://karger.com/ned/article/58/2/120/895963/Prevalence-and-Trajectories-of-Post-COVID-19)  Psychiatric and neurological complications of long COVID (2022), [*Journal of Psychiatric Research*](https://www.sciencedirect.com/science/article/pii/S0022395622005982)  Solving the puzzle of long Covid (2024), [*Science*](https://www.science.org/doi/10.1126/science.adl0867)  High-dimensional characterization of post-acute sequelae of COVID-19 (2021), [*Nature*](https://www.nature.com/articles/s41586-021-03553-9)  Data-driven identification of post-acute SARS-CoV-2 infection subphenotypes (2022), [*Nature Medicine*](https://www.nature.com/articles/s41591-022-02116-3) | **Summarization:**  Although nervous system disorders contribute to psychological symptoms, the neurological subtype of Long COVID focuses on conditions from the brain, spinal cord, and nerves, whereas the psychological subtype of Long COVID focuses on mental or behavior processes.  **Publications:**  Development of a Definition of Postacute Sequelae of SARS-CoV-2 Infection (2023), [*JAMA Network*](https://jamanetwork.com/journals/jama/fullarticle/2805540), [Supplementary Materials](https://cdn.jamanetwork.com/ama/content_public/journal/jama/939166/joi230062supp3_prod_1712695240.88184.pdf?Expires=1719182313&Signature=1DWb1eL9fHoDGO~Lajw5Y1vRNcYVX66275UgIbyhUjNHBvihGpYvk3BwSZ1zT45umhUYazDXV5H2wOvdPNyeVfPwJHvIzedWmin63WyDW9p~mtx3JaRUc5oJ3807SSIjOtN5sFtde0fboaLxMjjJCOn~Exkf2fOpGhZpGcnBnaXdEzimHjAB9TlbwteUnJJsHiZwUuADuW4YVXlSYG9sASDUMzPo2A-u1EFCM3bnDwUDc-Go48i1d9G0NDpcX8poU5WW5xBH8zOSyB1R228hA2ZL-AA7Nv-sXZ46ZGmDyC7PFXwzOGuAV59TjlAdHMy-Jz6QuwZpZUgyswmDcalmJQ__&Key-Pair-Id=APKAIE5G5CRDK6RD3PGA)  Defining the Subtypes of Long COVID and Risk Factors for Prolonged Disease: Population-Based Case-Crossover Study (2024), [*JMIR Publications*](https://publichealth.jmir.org/2024/1/e49841), [Suppl. Materials](https://docs.google.com/document/d/1Wte1k_dqrU9_YGrrS-j__qAoq8STjoKo/edit?usp=drive_link&ouid=107389695956544702385&rtpof=true&sd=true)  Psychiatric and neurological complications of long COVID (2022), [*Journal of Psychiatric Research*](https://www.sciencedirect.com/science/article/pii/S0022395622005982)  High-dimensional characterization of post-acute sequelae of COVID-19 (2021), [*Nature*](https://www.nature.com/articles/s41586-021-03553-9)  Data-driven identification of post-acute SARS-CoV-2 infection subphenotypes (2022), [*Nature Medicine*](https://www.nature.com/articles/s41591-022-02116-3)  Acute and postacute sequelae associated with SARS-CoV-2 reinfection (2022), [*Nature Medicine*](https://www.nature.com/articles/s41591-022-02051-3)  Prevalence of mental health conditions and brain fog in people with long COVID: A systematic review and meta-analysis (2024), [*General Hospital Psychiatry*](https://www.sciencedirect.com/science/article/pii/S0163834324000392) |

| **Subtype 3:**  Cardiovascular or circulatory | **Subtype 4:**  Respiratory |
| --- | --- |
| **ICD-11:**  **03** Diseases of the blood or blood-forming organs  **11** Diseases of the circulatory system | **ICD-11:**  **12** Diseases of the respiratory system |
| **Summarization:**  Observational studies of COVID-19 survivors indicate cardiovascular manifestations in Long COVID. Cardiovascular or circulatory subtype for Long COVID characterizes symptoms and conditions related to the heart, blood, and blood vessels.  **Publications:**  Long COVID: major findings, mechanisms and recommendations (2023), [*Nature Reviews Microbiology*](https://www.nature.com/articles/s41579-022-00846-2)  Development of a Definition of Postacute Sequelae of SARS-CoV-2 Infection (2023), [*JAMA Network*](https://jamanetwork.com/journals/jama/fullarticle/2805540), [Supplementary Materials](https://cdn.jamanetwork.com/ama/content_public/journal/jama/939166/joi230062supp3_prod_1712695240.88184.pdf?Expires=1719182313&Signature=1DWb1eL9fHoDGO~Lajw5Y1vRNcYVX66275UgIbyhUjNHBvihGpYvk3BwSZ1zT45umhUYazDXV5H2wOvdPNyeVfPwJHvIzedWmin63WyDW9p~mtx3JaRUc5oJ3807SSIjOtN5sFtde0fboaLxMjjJCOn~Exkf2fOpGhZpGcnBnaXdEzimHjAB9TlbwteUnJJsHiZwUuADuW4YVXlSYG9sASDUMzPo2A-u1EFCM3bnDwUDc-Go48i1d9G0NDpcX8poU5WW5xBH8zOSyB1R228hA2ZL-AA7Nv-sXZ46ZGmDyC7PFXwzOGuAV59TjlAdHMy-Jz6QuwZpZUgyswmDcalmJQ__&Key-Pair-Id=APKAIE5G5CRDK6RD3PGA)  Defining the Subtypes of Long COVID and Risk Factors for Prolonged Disease: Population-Based Case-Crossover Study (2024), [*JMIR Publications*](https://publichealth.jmir.org/2024/1/e49841), [Suppl. Materials](https://docs.google.com/document/d/1Wte1k_dqrU9_YGrrS-j__qAoq8STjoKo/edit?usp=drive_link&ouid=107389695956544702385&rtpof=true&sd=true)  Solving the puzzle of long Covid (2024), [*Science*](https://www.science.org/doi/10.1126/science.adl0867)  High-dimensional characterization of post-acute sequelae of COVID-19 (2021), [*Nature*](https://www.nature.com/articles/s41586-021-03553-9)  Data-driven identification of post-acute SARS-CoV-2 infection subphenotypes (2022), [*Nature Medicine*](https://www.nature.com/articles/s41591-022-02116-3)  Acute and postacute sequelae associated with SARS-CoV-2 reinfection (2022), [*Nature Medicine*](https://www.nature.com/articles/s41591-022-02051-3)  Cardiovascular Manifestations of the Long COVID Syndrome, [*Cardiology In Review*](https://journals.lww.com/cardiologyinreview/fulltext/9900/cardiovascular_manifestations_of_the_long_covid.97.aspx) | **Summarization:**  Individuals with Long COVID can experience ongoing pulmonary dysfunction with difficulties regaining normal lung function. Therefore, respiratory conditions are frequently explored as a group of Long COVID symptoms. Respiratory subtype includes issues with the lungs, nose, mouth, throat, and airways.  **Publications:**  Long COVID: major findings, mechanisms and recommendations (2023), [*Nature Reviews Microbiology*](https://www.nature.com/articles/s41579-022-00846-2)  Development of a Definition of Postacute Sequelae of SARS-CoV-2 Infection (2023), [*JAMA Network*](https://jamanetwork.com/journals/jama/fullarticle/2805540), [Supplementary Materials](https://cdn.jamanetwork.com/ama/content_public/journal/jama/939166/joi230062supp3_prod_1712695240.88184.pdf?Expires=1719182313&Signature=1DWb1eL9fHoDGO~Lajw5Y1vRNcYVX66275UgIbyhUjNHBvihGpYvk3BwSZ1zT45umhUYazDXV5H2wOvdPNyeVfPwJHvIzedWmin63WyDW9p~mtx3JaRUc5oJ3807SSIjOtN5sFtde0fboaLxMjjJCOn~Exkf2fOpGhZpGcnBnaXdEzimHjAB9TlbwteUnJJsHiZwUuADuW4YVXlSYG9sASDUMzPo2A-u1EFCM3bnDwUDc-Go48i1d9G0NDpcX8poU5WW5xBH8zOSyB1R228hA2ZL-AA7Nv-sXZ46ZGmDyC7PFXwzOGuAV59TjlAdHMy-Jz6QuwZpZUgyswmDcalmJQ__&Key-Pair-Id=APKAIE5G5CRDK6RD3PGA)  Defining the Subtypes of Long COVID and Risk Factors for Prolonged Disease: Population-Based Case-Crossover Study (2024), [*JMIR Publications*](https://publichealth.jmir.org/2024/1/e49841), [Suppl. Materials](https://docs.google.com/document/d/1Wte1k_dqrU9_YGrrS-j__qAoq8STjoKo/edit?usp=drive_link&ouid=107389695956544702385&rtpof=true&sd=true)  High-dimensional characterization of post-acute sequelae of COVID-19 (2021), [*Nature*](https://www.nature.com/articles/s41586-021-03553-9)  Data-driven identification of post-acute SARS-CoV-2 infection subphenotypes (2022), [*Nature Medicine*](https://www.nature.com/articles/s41591-022-02116-3)  Acute and postacute sequelae associated with SARS-CoV-2 reinfection (2022), [*Nature Medicine*](https://www.nature.com/articles/s41591-022-02051-3) |

| **Subtype 5:**  Gastrointestinal | **Subtype 6:**  Renal |
| --- | --- |
| **ICD-11:**  **13** Diseases of the digestive system | **ICD-11:**  **16** Diseases of the genitourinary system |
| **Summarization:**  There is emerging evidence for digestive problems, such as dyspepsia, diarrhea, constipation, in Long COVID. By classifying gastrointestinal symptoms and conditions as a subtype of Long COVID, prevalence and risk factors can be accessed for this subtype that includes issues with the mouth and teeth, esophagus, stomach, and intestines.  **Publications:**  Long COVID: major findings, mechanisms and recommendations (2023), [*Nature Reviews Microbiology*](https://www.nature.com/articles/s41579-022-00846-2)  Development of a Definition of Postacute Sequelae of SARS-CoV-2 Infection (2023), [*JAMA Network*](https://jamanetwork.com/journals/jama/fullarticle/2805540), [Supplementary Materials](https://cdn.jamanetwork.com/ama/content_public/journal/jama/939166/joi230062supp3_prod_1712695240.88184.pdf?Expires=1719182313&Signature=1DWb1eL9fHoDGO~Lajw5Y1vRNcYVX66275UgIbyhUjNHBvihGpYvk3BwSZ1zT45umhUYazDXV5H2wOvdPNyeVfPwJHvIzedWmin63WyDW9p~mtx3JaRUc5oJ3807SSIjOtN5sFtde0fboaLxMjjJCOn~Exkf2fOpGhZpGcnBnaXdEzimHjAB9TlbwteUnJJsHiZwUuADuW4YVXlSYG9sASDUMzPo2A-u1EFCM3bnDwUDc-Go48i1d9G0NDpcX8poU5WW5xBH8zOSyB1R228hA2ZL-AA7Nv-sXZ46ZGmDyC7PFXwzOGuAV59TjlAdHMy-Jz6QuwZpZUgyswmDcalmJQ__&Key-Pair-Id=APKAIE5G5CRDK6RD3PGA)  Defining the Subtypes of Long COVID and Risk Factors for Prolonged Disease: Population-Based Case-Crossover Study (2024), [*JMIR Publications*](https://publichealth.jmir.org/2024/1/e49841), [Suppl. Materials](https://docs.google.com/document/d/1Wte1k_dqrU9_YGrrS-j__qAoq8STjoKo/edit?usp=drive_link&ouid=107389695956544702385&rtpof=true&sd=true)  Solving the puzzle of long Covid (2024), [*Science*](https://www.science.org/doi/10.1126/science.adl0867)  High-dimensional characterization of post-acute sequelae of COVID-19 (2021), [*Nature*](https://www.nature.com/articles/s41586-021-03553-9)  Data-driven identification of post-acute SARS-CoV-2 infection subphenotypes (2022), [*Nature Medicine*](https://www.nature.com/articles/s41591-022-02116-3)  Acute and postacute sequelae associated with SARS-CoV-2 reinfection (2022), [*Nature Medicine*](https://www.nature.com/articles/s41591-022-02051-3) | **Summarization:**  The impact of COVID-19 on kidneys remains equivocal. Publications that examined Long COVID and kidney functions grouped kidney conditions based on Common Terminology Criteria for Adverse Events and medical specialty. Having renal or kidney conditions as a subtype of Long COVID would allow us to examine worldwide prevalence and risk factors, especially diabetes and kidney disease in a patient's past health history.  **Publications:**  Development of a Definition of Postacute Sequelae of SARS-CoV-2 Infection (2023), [*JAMA Network*](https://jamanetwork.com/journals/jama/fullarticle/2805540), [Supplementary Materials](https://cdn.jamanetwork.com/ama/content_public/journal/jama/939166/joi230062supp3_prod_1712695240.88184.pdf?Expires=1719182313&Signature=1DWb1eL9fHoDGO~Lajw5Y1vRNcYVX66275UgIbyhUjNHBvihGpYvk3BwSZ1zT45umhUYazDXV5H2wOvdPNyeVfPwJHvIzedWmin63WyDW9p~mtx3JaRUc5oJ3807SSIjOtN5sFtde0fboaLxMjjJCOn~Exkf2fOpGhZpGcnBnaXdEzimHjAB9TlbwteUnJJsHiZwUuADuW4YVXlSYG9sASDUMzPo2A-u1EFCM3bnDwUDc-Go48i1d9G0NDpcX8poU5WW5xBH8zOSyB1R228hA2ZL-AA7Nv-sXZ46ZGmDyC7PFXwzOGuAV59TjlAdHMy-Jz6QuwZpZUgyswmDcalmJQ__&Key-Pair-Id=APKAIE5G5CRDK6RD3PGA)  Defining the Subtypes of Long COVID and Risk Factors for Prolonged Disease: Population-Based Case-Crossover Study (2024), [*JMIR Publications*](https://publichealth.jmir.org/2024/1/e49841), [Suppl. Materials](https://docs.google.com/document/d/1Wte1k_dqrU9_YGrrS-j__qAoq8STjoKo/edit?usp=drive_link&ouid=107389695956544702385&rtpof=true&sd=true)  Data-driven identification of post-acute SARS-CoV-2 infection subphenotypes (2022), [*Nature Medicine*](https://www.nature.com/articles/s41591-022-02116-3)  Acute and postacute sequelae associated with SARS-CoV-2 reinfection (2022), [*Nature Medicine*](https://www.nature.com/articles/s41591-022-02051-3) |

| **Subtype 7:**  Musculoskeletal | **Subtype 8:**  Dermatologic symptoms |
| --- | --- |
| **ICD-11:**  **15** Diseases of the musculoskeletal system or connective tissue | **ICD-11:**  **14** Diseases of the skin |
| **Summarization:**  Some of the most common Long COVID symptoms are joint pain and muscle aches.  Musculoskeletal subtype includes problems with bones, muscles, and joints (addition inclusions are cartilage, tendons, and ligaments issues).  **Publications:**  Development of a Definition of Postacute Sequelae of SARS-CoV-2 Infection (2023), [*JAMA Network*](https://jamanetwork.com/journals/jama/fullarticle/2805540), [Supplementary Materials](https://cdn.jamanetwork.com/ama/content_public/journal/jama/939166/joi230062supp3_prod_1712695240.88184.pdf?Expires=1719182313&Signature=1DWb1eL9fHoDGO~Lajw5Y1vRNcYVX66275UgIbyhUjNHBvihGpYvk3BwSZ1zT45umhUYazDXV5H2wOvdPNyeVfPwJHvIzedWmin63WyDW9p~mtx3JaRUc5oJ3807SSIjOtN5sFtde0fboaLxMjjJCOn~Exkf2fOpGhZpGcnBnaXdEzimHjAB9TlbwteUnJJsHiZwUuADuW4YVXlSYG9sASDUMzPo2A-u1EFCM3bnDwUDc-Go48i1d9G0NDpcX8poU5WW5xBH8zOSyB1R228hA2ZL-AA7Nv-sXZ46ZGmDyC7PFXwzOGuAV59TjlAdHMy-Jz6QuwZpZUgyswmDcalmJQ__&Key-Pair-Id=APKAIE5G5CRDK6RD3PGA)  Data-driven identification of post-acute SARS-CoV-2 infection subphenotypes (2022), [*Nature Medicine*](https://www.nature.com/articles/s41591-022-02116-3)  Acute and postacute sequelae associated with SARS-CoV-2 reinfection (2022), [*Nature Medicine*](https://www.nature.com/articles/s41591-022-02051-3) | **Summarization:**  Those diagnosed with COVID-19 may have ongoing hair and skin problems, which constitute a dermatological subtype for Long COVID.  **Publications:**  Development of a Definition of Postacute Sequelae of SARS-CoV-2 Infection (2023), [*JAMA Network*](https://jamanetwork.com/journals/jama/fullarticle/2805540), [Supplementary Materials](https://cdn.jamanetwork.com/ama/content_public/journal/jama/939166/joi230062supp3_prod_1712695240.88184.pdf?Expires=1719182313&Signature=1DWb1eL9fHoDGO~Lajw5Y1vRNcYVX66275UgIbyhUjNHBvihGpYvk3BwSZ1zT45umhUYazDXV5H2wOvdPNyeVfPwJHvIzedWmin63WyDW9p~mtx3JaRUc5oJ3807SSIjOtN5sFtde0fboaLxMjjJCOn~Exkf2fOpGhZpGcnBnaXdEzimHjAB9TlbwteUnJJsHiZwUuADuW4YVXlSYG9sASDUMzPo2A-u1EFCM3bnDwUDc-Go48i1d9G0NDpcX8poU5WW5xBH8zOSyB1R228hA2ZL-AA7Nv-sXZ46ZGmDyC7PFXwzOGuAV59TjlAdHMy-Jz6QuwZpZUgyswmDcalmJQ__&Key-Pair-Id=APKAIE5G5CRDK6RD3PGA)  Defining the Subtypes of Long COVID and Risk Factors for Prolonged Disease: Population-Based Case-Crossover Study (2024), [*JMIR Publications*](https://publichealth.jmir.org/2024/1/e49841), [Suppl. Materials](https://docs.google.com/document/d/1Wte1k_dqrU9_YGrrS-j__qAoq8STjoKo/edit?usp=drive_link&ouid=107389695956544702385&rtpof=true&sd=true) |

| **Subtype 9:**  Endocrine or metabolic | **Subtype 10:**  General symptoms/conditions |
| --- | --- |
| **ICD-11:**  **05** Endocrine, nutritional or metabolic diseases | **ICD-11:**  **21** Systems, signs or clinical findings, not elsewhere classified (Symptoms of the immune system; symptoms, signs or clinical findings of the nervous system; general symptoms, signs or clinical findings) |
| **Summarization:**  There can be persisting hormonal and metabolic changes in patients with Long COVID. The endocrine system is responsible for hormone production and regulation, metabolism, and reproduction. Endocrine and metabolic disorders constitute a subtype for Long COVID.  **Publications:**  Long COVID: major findings, mechanisms and recommendations (2023), [*Nature Reviews Microbiology*](https://www.nature.com/articles/s41579-022-00846-2)  Development of a Definition of Postacute Sequelae of SARS-CoV-2 Infection (2023), [*JAMA Network*](https://jamanetwork.com/journals/jama/fullarticle/2805540), [Supplementary Materials](https://cdn.jamanetwork.com/ama/content_public/journal/jama/939166/joi230062supp3_prod_1712695240.88184.pdf?Expires=1719182313&Signature=1DWb1eL9fHoDGO~Lajw5Y1vRNcYVX66275UgIbyhUjNHBvihGpYvk3BwSZ1zT45umhUYazDXV5H2wOvdPNyeVfPwJHvIzedWmin63WyDW9p~mtx3JaRUc5oJ3807SSIjOtN5sFtde0fboaLxMjjJCOn~Exkf2fOpGhZpGcnBnaXdEzimHjAB9TlbwteUnJJsHiZwUuADuW4YVXlSYG9sASDUMzPo2A-u1EFCM3bnDwUDc-Go48i1d9G0NDpcX8poU5WW5xBH8zOSyB1R228hA2ZL-AA7Nv-sXZ46ZGmDyC7PFXwzOGuAV59TjlAdHMy-Jz6QuwZpZUgyswmDcalmJQ__&Key-Pair-Id=APKAIE5G5CRDK6RD3PGA)  Defining the Subtypes of Long COVID and Risk Factors for Prolonged Disease: Population-Based Case-Crossover Study (2024), [*JMIR Publications*](https://publichealth.jmir.org/2024/1/e49841), [Suppl. Materials](https://docs.google.com/document/d/1Wte1k_dqrU9_YGrrS-j__qAoq8STjoKo/edit?usp=drive_link&ouid=107389695956544702385&rtpof=true&sd=true)  Solving the puzzle of long Covid (2024), [*Science*](https://www.science.org/doi/10.1126/science.adl0867)  High-dimensional characterization of post-acute sequelae of COVID-19 (2021), [*Nature*](https://www.nature.com/articles/s41586-021-03553-9) | **Summarization:**  General subtype includes symptoms that are not specific for other subtypes (appetite, fatigue, fever, etc.) or are not fitting for other subtypes (autoimmunity, immunosuppression, viral persistence, etc.)  **Publications:**  Long COVID: major findings, mechanisms and recommendations (2023), [*Nature Reviews Microbiology*](https://www.nature.com/articles/s41579-022-00846-2)  Development of a Definition of Postacute Sequelae of SARS-CoV-2 Infection (2023), [*JAMA Network*](https://jamanetwork.com/journals/jama/fullarticle/2805540), [Supplementary Materials](https://cdn.jamanetwork.com/ama/content_public/journal/jama/939166/joi230062supp3_prod_1712695240.88184.pdf?Expires=1719182313&Signature=1DWb1eL9fHoDGO~Lajw5Y1vRNcYVX66275UgIbyhUjNHBvihGpYvk3BwSZ1zT45umhUYazDXV5H2wOvdPNyeVfPwJHvIzedWmin63WyDW9p~mtx3JaRUc5oJ3807SSIjOtN5sFtde0fboaLxMjjJCOn~Exkf2fOpGhZpGcnBnaXdEzimHjAB9TlbwteUnJJsHiZwUuADuW4YVXlSYG9sASDUMzPo2A-u1EFCM3bnDwUDc-Go48i1d9G0NDpcX8poU5WW5xBH8zOSyB1R228hA2ZL-AA7Nv-sXZ46ZGmDyC7PFXwzOGuAV59TjlAdHMy-Jz6QuwZpZUgyswmDcalmJQ__&Key-Pair-Id=APKAIE5G5CRDK6RD3PGA)  Solving the puzzle of long Covid (2024), [*Science*](https://www.science.org/doi/10.1126/science.adl0867)  High-dimensional characterization of post-acute sequelae of COVID-19 (2021), [*Nature*](https://www.nature.com/articles/s41586-021-03553-9) |
