## Supplementary material for "Global Prevalence of Long COVID, its Subtypes and Risk factors: An Updated Systematic Review and Meta-Analysis": eMethods 1d. Supplementary search strategy

***eMethods 1d*.** Systematic review extended search strategies

**Supplementary Search:**

Scanning tables of contents of key journals

- Journals: (publication period 01/01/2024 - 07/23/2024)

The New England Journal of Medicine

The Lancet

JAMA and JAMA Network

Nature Medicine

PloS

BMJ

Annals of Internal Medicine

Gray literature searches

- Latin American and Caribbean Health Sciences Literature, [LILACS](https://pesquisa.bvsalud.org/portal/?u_filter%5B%5D=fulltext&u_filter%5B%5D=db&u_filter%5B%5D=mj_cluster&u_filter%5B%5D=type_of_study&u_filter%5B%5D=la&fb=&output=site&lang=en&from=1&sort=&format=summary&count=100&page=1&range_year_start=2021&range_year_end=2024&skfp=&index=&q=%28%22Post+COVID-19+Condition%22+OR+%22Long+COVID%22+OR+%22Post-acute+sequelae+of+COVID-19%22%29+AND+%28%22Prevalence%22+OR+%22Risk+factors%22%29&where=&filter%5Bla%5D%5B%5D=en&range_year_start=2024&range_year_end=2024)

**Date searched:** 07/23/2024

**Number of results:** 497

**Date filter:** 2024 - 2024

Title, abstract, subject:

("Post COVID-19 Condition" OR "Long COVID" OR "Post-acute sequelae of COVID-19") AND ("Prevalence" OR "Risk factors")

Language = English

- Google Scholar, [Google scholar](https://scholar.google.com/scholar?start=0&q=(Post+COVID-19+Condition+OR+Long+COVID+or+Post-acute+sequelae+of+COVID-19)+AND+(Prevalence+OR+Risk+factors)&hl=en&scisbd=1&as_sdt=0,23)

**Date searched:** 07/23/2024

**Number of results:** 70

**Date filter:** 2024 - 2024 (sort by date)

(Post COVID-19 Condition OR Long COVID or Post-acute sequelae of COVID-19) AND (Prevalence OR Risk factors)

Second updated search from first search in May 29, 2024:

1 AND 2 AND (3 OR 4 OR 5 OR 6 OR 7 OR 8 OR 9 OR 10 OR 11 OR 12)

Web of Science Core Collection

**Date searched:** 07/23/2024

**Number of results:** 142

**Date filter:** 05/29/2024 - present

**Other filters applied:** Search in TOPIC field
